# GRAd-COV2 vaccine provides potent and durable immunity in randomised placebo-controlled phase 2 trial (COVITAR)

**DOI:** 10.1101/2022.10.08.22280836

**Authors:** Stefania Capone, Francesco M. Fusco, Stefano Milleri, Silvio Borrè, Sergio Carbonara, Sergio Lo Caputo, Sebastiano Leone, Giovanni Gori, Paolo Maggi, Antonio Cascio, Miriam Lichtner, Roberto Cauda, Sarah Dal Zoppo, Maria V. Cossu, Andrea Gori, Silvia Roda, Paola Confalonieri, Stefano Bonora, Gabriele Missale, Mauro Codeluppi, Ivano Mezzaroma, Serena Capici, Emanuele Pontali, Marco Libanore, Augusta Diani, Simone Lanini, Simone Battella, Alessandra M. Contino, Eva Piano Mortari, Francesco Genova, Gessica Parente, Rosella Dragonetti, Stefano Colloca, Luigi Visani, Claudio Iannacone, Rita Carsetti, Antonella Folgori, Roberto Camerini, COVITAR study group

**Author notes:** Trial group members are listed in the supplemental material. Correspondence to: Stefania Capone, Department of Preclinical and Clinical Immunology, ReiThera srl, Rome, 00128, Italy.

## Abstract

**Background:** SARS-CoV-2 ongoing pandemic and heterologous immunization approaches implemented worldwide for booster doses call for diversified vaccines portfolio. We report safety and immunogenicity of GRAd-COV2, a novel gorilla adenovirus-based COVID-19 vaccine, in a phase 2 trial aimed at identifying the appropriate dose and schedule.

**Method:** 917 eligible adults aged 18 years or older, including participants with co-morbidities, were randomised to receive, 21 days apart, a single vaccine administration at 2×10^11^ viral particles (vp) followed by placebo, or repeated vaccine administration at 1×10^11^ vp, or two doses of placebo. Primary endpoints were the incidence of local and systemic solicited AEs for 7 days post each dose and the post-treatment (35 days after the first dose), geometric mean titers (GMTs) and geometric mean fold rise (GMFRs) of ELISA antibody responses to Spike protein. Additional humoral and cellular immune response parameters were monitored for up to six months.

**Results:** The safety profile of GRAd-COV2 was characterized by short-term, mild-to-moderate pain and tenderness at injection site, fatigue, headache, malaise, and myalgia. Neither related SAEs nor deaths were reported. Humoral (binding and neutralizing) Ab responses peaked at day 35 after a single administration, were boosted by a second vaccination, were sustained until day 57 to then decline at day 180. Potent, VOC cross-reactive T cell responses peaked already after first dose with high frequencies of long-lived CD8 T cells.

**Conclusion:** GRAd-COV2 was safe, and induced robust immune responses after a single immunization; the second administration increased humoral but not cellular immune responses.

**Trial Registration:** ClinicalTrials.gov NCT04791423.

**Funding:** ReiThera Srl

## Introduction

Since mid 2022 COVID-19 vaccine supply is no longer a limiting factor in efforts to control the global pandemic (1). Over 350 COVID-19 vaccine candidates have been developed or are in development using different technology platforms (2). The need for a range of vaccines is due to the fact that multiple factors influence policy decisions and each vaccine has distinctive features, advantages and disadvantages to be considered in different health care settings, economies, subpopulations and age groups.

Moreover, the continued emergence of new SARS-CoV-2 variants of concern (VOC) is adding complexity for vaccine developers and for policy decision makers. Omicron and its sublineages, with an unprecedented mutation burden focused in the Spike protein, have rapidly displaced previous circulating variants since late 2021. Antigenic changes leading to significant evasion from humoral immunity induced either by infection with other SARS-CoV-2 variants or by vaccination, together with functional and structural modifications affecting transmissibility and pathogenicity (3, 4) call for considering omicron a distinct SARS-CoV-2 serotype that needs vaccine adaptation (5, 6). However, such an approach may be practically unfeasible given the speed at which novel variants have emerged and then disappeared and if the future variants will not linearly evolve from the latest circulating variants. Preclinical and real-world data suggest that, following repeated prototype-based vaccine booster doses, the cross-reactivity of neutralizing response is widely improved while variant-specific adapted vaccines seem to generate a more restricted immunity (7-10). While initial data on bivalent vaccines are encouraging (11), the safety and immunogenicity profile associated to novel candidate vaccines based on the prototype Spike are still of interest and enable crucial direct comparisons across a range of traditional and innovative vaccine platforms, an unprecedented circumstance before the COVID-19 era.

Adenoviral vaccine platform has successfully been exploited in at least 4 effective and widely approved COVID-19 vaccines: Vaxzevria, Jcovden, Sputnik V and Convidecia. GRAd-COV2 is a candidate vaccine based on a novel non-replicating gorilla group C adenovirus encoding for a prefusion stabilized full length Spike. The vaccine induced potent and durable humoral and Th1-skewed cellular immune response upon a single intramuscular administration in animal models (12) and healthy adult volunteers within a phase 1, dose escalation trial (13). The vaccine was well tolerated (14) and the safety profile was similar in terms of quality and severity to that of other COVID-19 genetic vaccines.

Here, we expanded GRAd-COV2 safety and immunogenicity evaluation in a phase 2 trial, where we also compared a two-dose versus a single dose regimen, with the aim to select the best vaccination schedule to be further progressed in efficacy studies.

These relevant clinical data in humans are of more general interest for deepening our understanding of the immunological features embedded in innovative genetic vaccine platforms that have finally demonstrated all their potential in the COVID-19 pandemic but that have much broader applications for future emerging pathogens or for the immunotherapy of cancer.

## Results

### Trial population

Between March 18, 2021 and April 9, 2021, a total of 923 volunteers older than 18 years were screened and 917 randomised. A total of 917 participants were dosed: 305 were assigned to receive a single dose of GRAd-COV2 plus placebo, 308 a repeated dose of GRAd-COV2, and 304 placebo (Figure 1). 652 participants (71.1%) belonged to the stratum of < 65 years of age without risk factors, 90 participants (9.8%) to the stratum of <65 years of age with risk factors, and 175 (19.1%) participants to the stratum of ≥65 years of age. Baseline characteristics of overall population are reported in table 1. On 21 June 2021, following the DSMB recommendation and EC approval, the randomisation code was broken to allow participants assigned to placebo group to access to the public vaccination campaign that was implemented in Italy during the first quarter of 2021, hence the blinding was maintained up to day 57. Participants in placebo group who received marketed COVID-19 vaccines withdrew from the study while the participants assigned to vaccine groups continued to be followed-up for 1 year. This report considers all participants dosed with GRAd-COV2 vaccine who completed 180 days follow-up for safety and assessment of immunological parameters.

**Figure 1.**
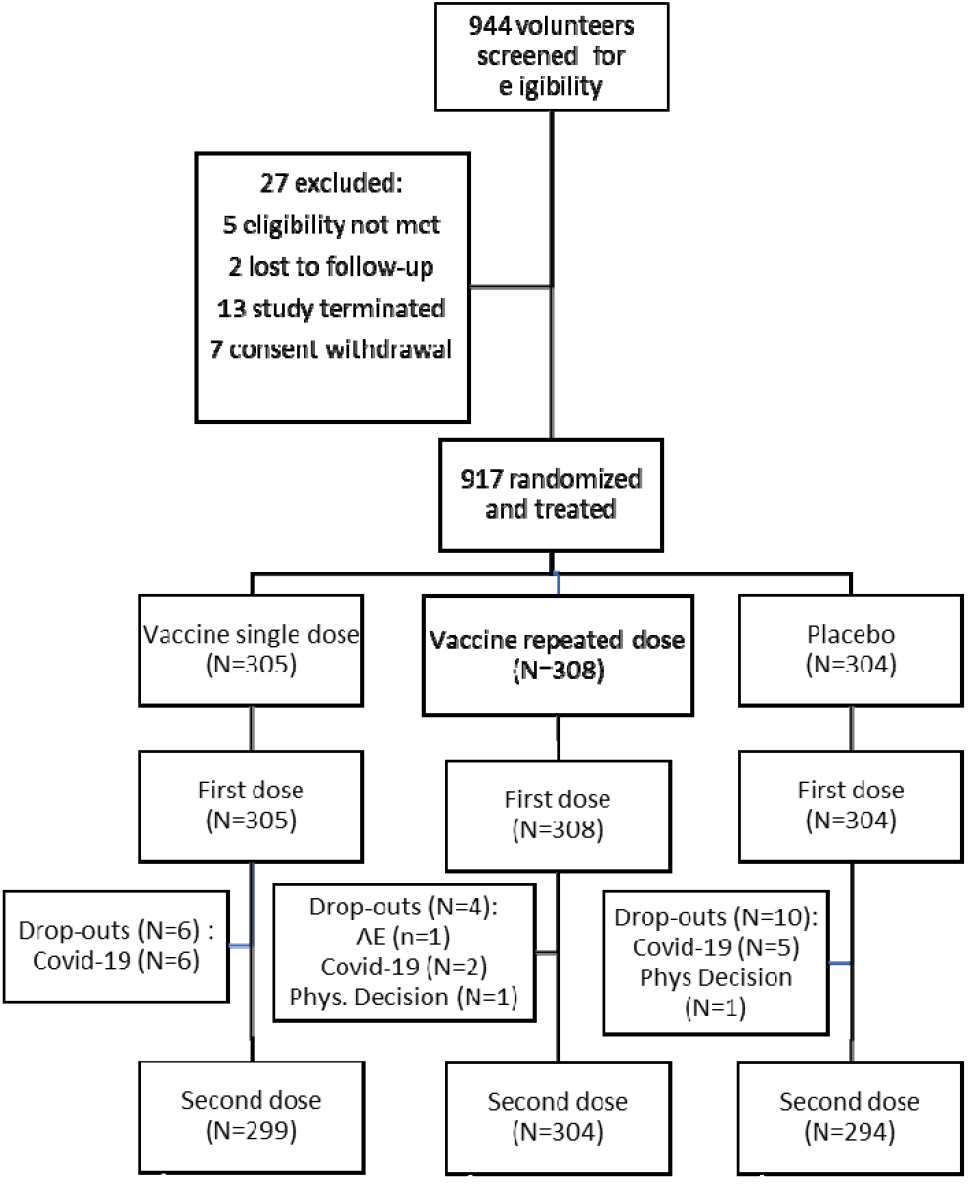
Trial Profile. All participants were followed-up in blind condition up to day 57 after the first dose, afterword the randomisation code was opened to allow participants assigned to placebo group to have access to the vaccination campaign.

**Table 1.** Participants Baseline characteristics.

|  | **Vaccine single dose**<br>**at 2 x 10<sup>11</sup> vp (n=305)** | **Vaccine repeated dose**<br>**at 1 x 10<sup>11</sup> vp (n=308)** | **Placebo**<br>**(n=304)** |
| --- | --- | --- | --- |
| **Protocol Strata** |  |  |  |
| Age 18-<65 years &<br>not at risk n (%) | 218 (71.5) | 219 (71.1) | 215 (70.7) |
| Age 18-<65 years &<br>at risk n (%) | 29 (9.5) | 30 (9.7) | 31 (10.2) |
| Age ≥65 years<br>n (%) | 58 (19.0) | 59 (19.2) | 58 (19.1) |
| **Age, years** |  |  |  |
| Mean (SD) | 46.0 (16.63) | 47.2 (16.07) | 47.3 (15.54) |
| **Sex n (%)** |  |  |  |
| Male | 197 (64.6) | 200 (64.9) | 184 (60.5) |
| Female | 108 (35.4) | 108 (35.1) | 120 (39.5) |
| **Body mass index, kg/m<sup>2</sup>** |  |  |  |
| Mean (SD) | 25.3 (4.4) | 25.0 (4.4) | 25.6 (4.6) |
| **Underlying diseases n (%)** |  |  |  |
| None | 267 (87.5) | 270 (87.7) | 261 (85.9) |
| Significant cardiac<br>diseases | 12 (3.9) | 14 (4.5) | 17 (5.6) |
| Chronic lung diseases | 9 (3.0) | 8 (2.6) | 6 (2.0) |
| Severe obesity | 3 (1.0) | 3 (1.0) | 6 (2.0) |
| Diabetes<br>(type 1, type 2) | 7 (2.3) | 6 (1.9) | 6 (2.0) |
| Liver diseases | 2 (0.7) | 3 (1.0) | 3 (1.0) |
| HIV infection | 9 (3.0) | 7 (2.3) | 6 (2.0) |

### Reactogenicity and safety

Overall, GRAd-COV2 recipients, both in single and repeated dose group, reported more local reactions than placebo recipients (80.7%, 75.3%, and 28 %, respectively after the first dose). Among GRAd-COV2 recipients, mild-to-moderate pain and tenderness at the injection site was the most commonly reported local reaction (Figure 2A and 2B). Among GRAd-COV2 recipients receiving the second dose, local reactions, after the second injection were reported less frequently (68.5%, Figure 2B). Participants ≥ 65 years of age reported less frequent local reactions (69%, 50.8%, and 17.2%, respectively in the three groups after the first dose) than younger participants (83.8%; 81.5%, and 30.5%, respectively, Figure 2C). A noticeably lower percentage of participants reported injection-site erythema or swelling. No participants reported a grade 4 local reaction (Figure 3). In general, local reactions were mostly mild-to-moderate in severity and resolved within 2 days.

**Figure 2.**
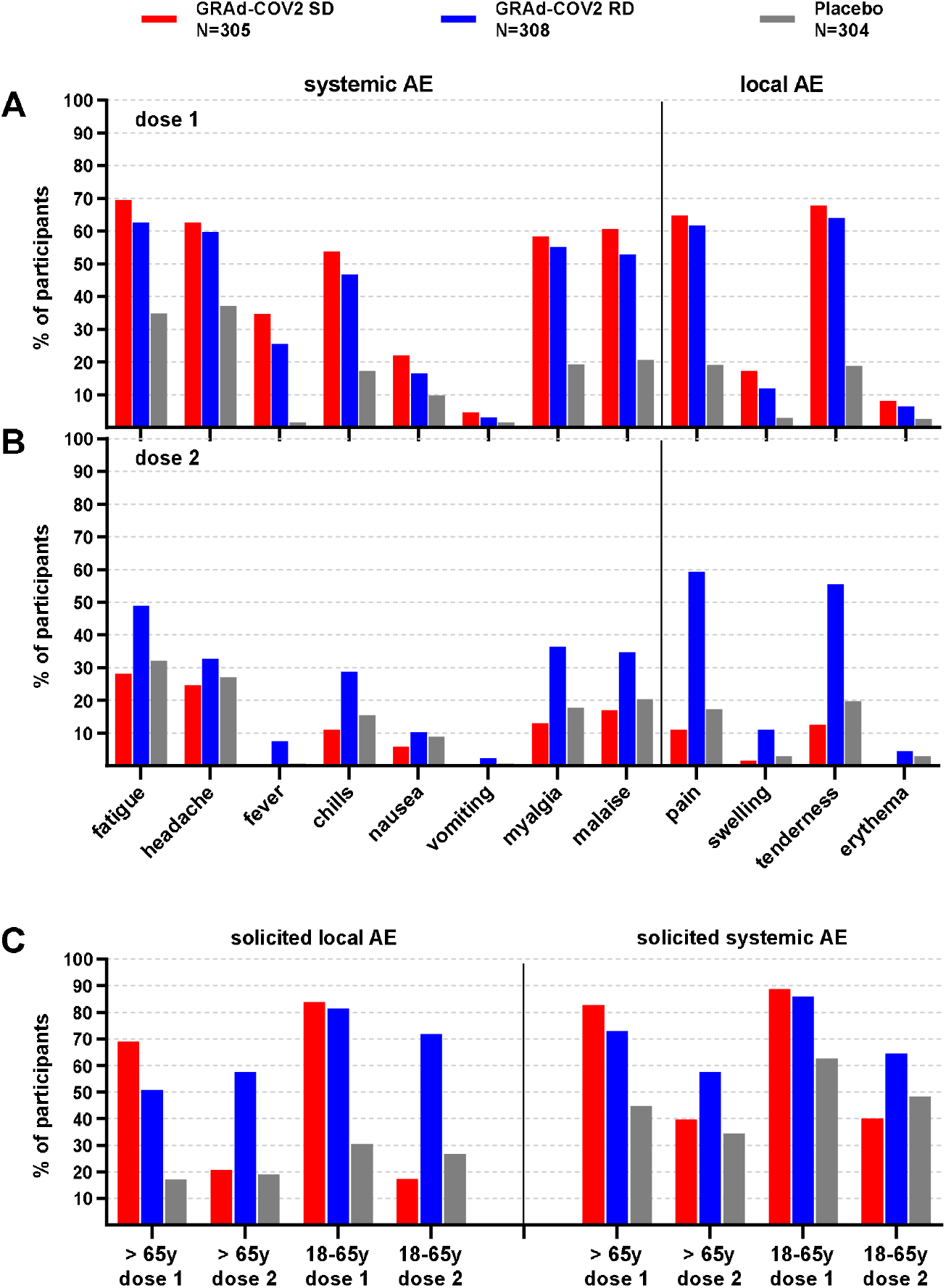
Frequency of participants with solicited systemic and local adverse events. Data are percentage of participants. (**A**): frequency of participants with solicited systemic and local AEs within 7 days after the first dose. (**B**): frequency of participants with solicited systemic and local AEs within 7 days after the second dose. Only events with a frequency ≥ 1% are reported. (**C**): frequency of participants with solicited local and systemic AEs (any) within 7 days after the first and second dose according to the age category (18-65 years; > 65 years).

**Figure 3.**
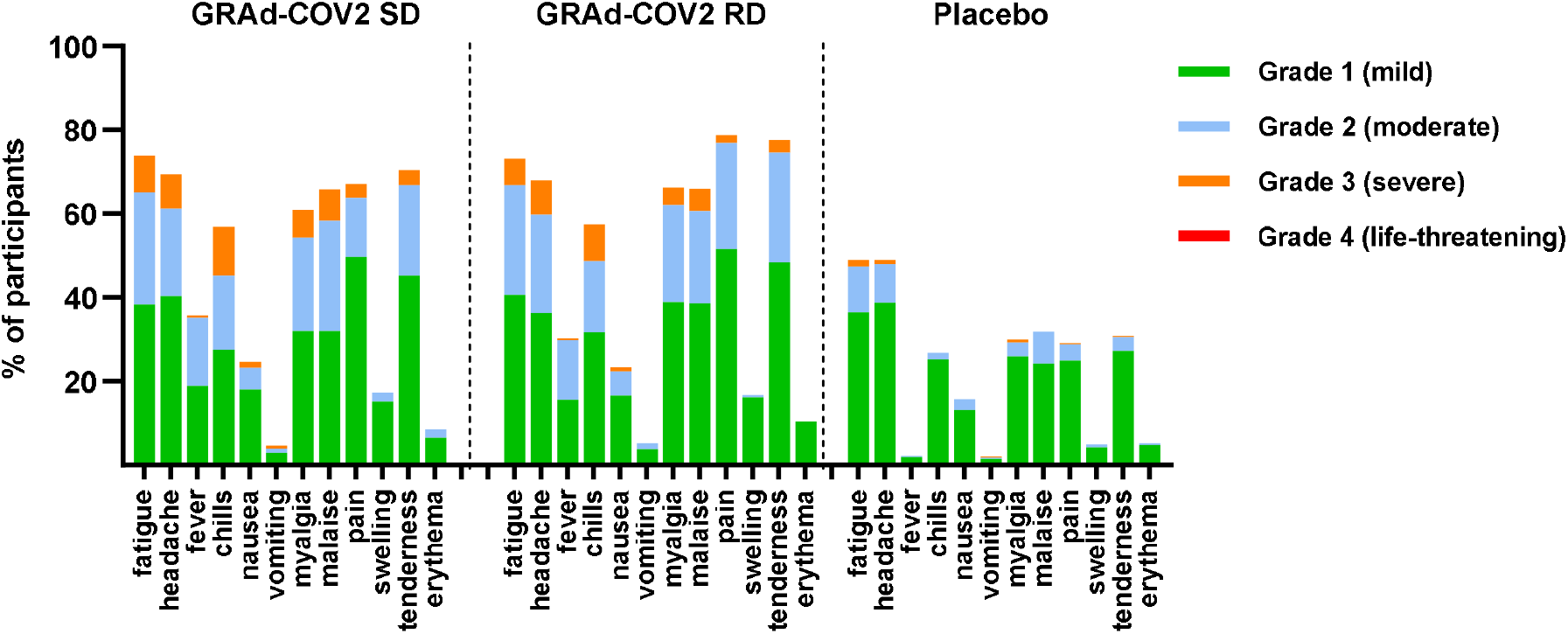
Severity of adverse events. Data are percentages of participants. Most frequent (>1%) AEs (solicited and unsolicited) within 28 days after any dose of vaccination. Severity was assessed for AEs according to toxicity grading scales modified and abridged from the US FDA guidance. No grade 4 AEs were observed.

Both groups of GRAd-COV2 recipients reported more solicited systemic reactions than placebo group (87.5%, 82.1%, and 59.2%, respectively after the first dose Figure 2A). Systemic solicited events were reported more often by younger than by older vaccine recipients and less often after dose 2 than dose 1 (Figure 2C). The most commonly reported solicited systemic AEs were fatigue, headache, malaise, myalgia, and chills, although fatigue and headache were also reported by many placebo recipients. Fever, nausea, and vomiting were less reported. Among GRAd-COV2 recipients the frequency of severe solicited systemic events was slightly higher in the group receiving single 2×10^11^ vp then in the group receiving 1×10^11^ vp repeated dose (Figure 3). Systemic AEs were generally mild to moderate and observed within the first 2 days after vaccination and resolved shortly thereafter. No grade 4 severity was reported.

Unsolicited AEs analyses are provided for all enrolled 917 participants, with a follow-up time of 28 days after dose 2. Comparable rate of GRAd-COV2 single dose, repeated dose, and placebo recipients reported any unsolicited AE (16.7%, 13.6% and 14.1%, respectively) or a related unsolicited AE (3.6%, 3.9%, and 3%). Very few participants in all groups had severe unsolicited AEs (0, 0.3%, and 0.3%, respectively). Neither AEs leading to withdrawal nor related SAEs or deaths were reported. Detailed description of solicited and unsolicited AEs is reported in Supplemental Table 1.

### Serology

By 21 days after the first dose, Spike binding IgG were induced in the majority of participants (Figure 4A and Table 2); similar GMT (40.54 and 41.13 AU/ml) and seroconversion rates (93.5% and 92.5%) were observed between single dose (SD) and repeated dose (RD) arms, despite the 2-fold difference in vaccine dosage (2×10^11^ and 1×10^11^ vp respectively for SD and RD arms). Spike IgG titers in the SD arm peaked at d36 (GMT 45.15 AU/ml) and remained quite stable until d57, to then contract around 3-fold by 6 months (GMT 13.40 AU/ml at d180). The effect of a second GRAd-COV2 administration was evident at d36 (14d post dose 2 in RD arm), with a significant increase of Spike binding IgG (GMT 77.94 AU/ml, P=0.0001), and seroconversion rate up to 99.3% in RD arm. Titers were diminished at d57 (GMT 55.74 AU/ml) but still significantly higher than in SD arm (P=0.001), to then decline to similar levels at d180 (GMT 15.50 AU/ml), a 5-fold contraction with respect to d36 peak.

**Figure 4:**
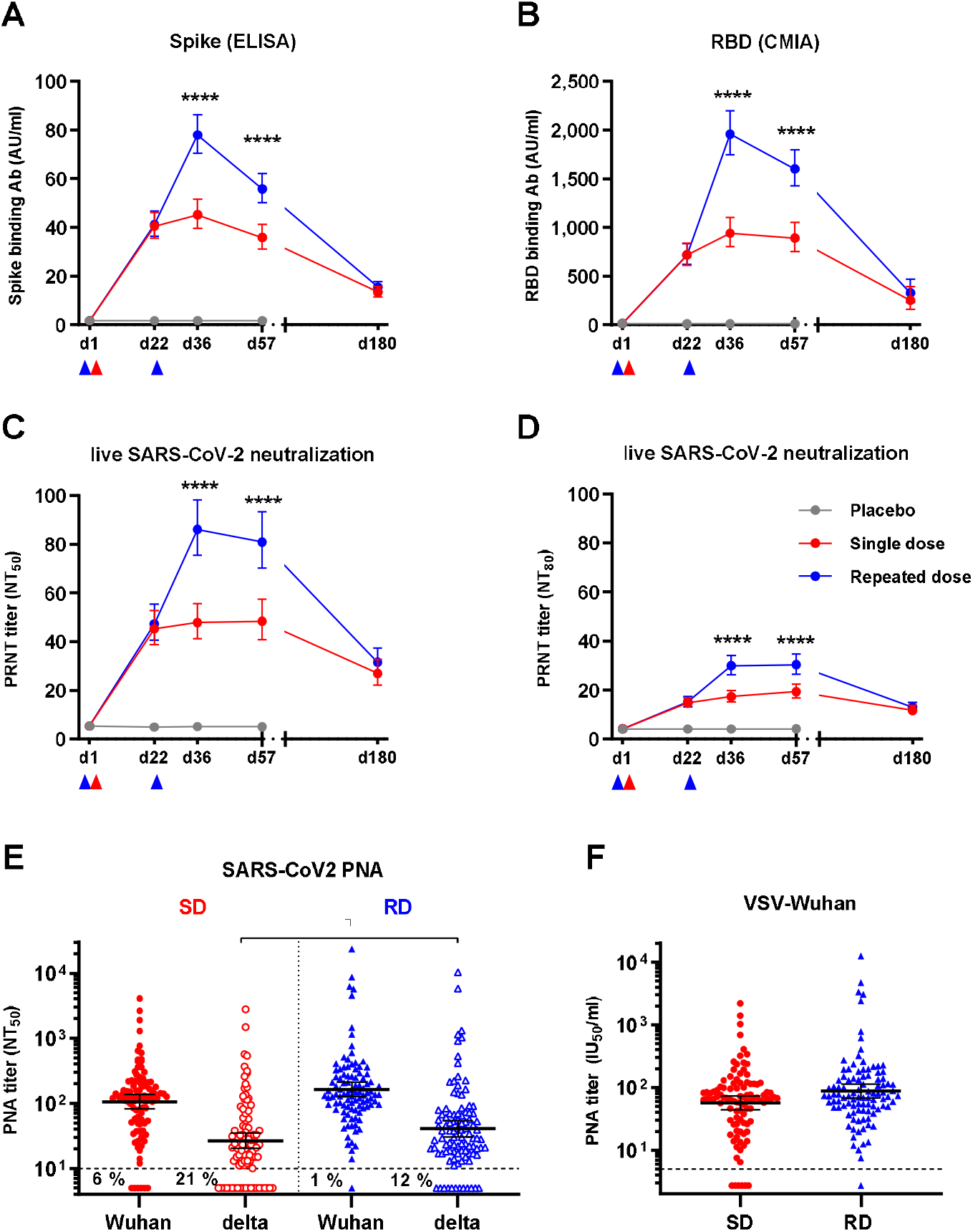
Spike/RBD binding and SARS-CoV2 neutralizing antibody kinetics in GRAd-COV2 vaccinated and placebo arms. The magnitude and kinetics of antibodies binding to full length trimeric Spike (**A**), RBD (B) and of live SARS-CoV-2 neutralizing antibodies, expressed as 50% (**C**) or 80% neutralizing titer (**D**), following GRAd-COV2 or placebo administration are reported over 6 months follow up. Datapoints are the geometric mean (GM) and 95% confidence interval (CI) at each study visit for each study arm. For binding antibodies (panels **A** and **B**), data are expressed as arbitrary units (AU)/ml, as per assay manufacturer datasheet. Arrowheads below x axes indicate vaccination. Statistical analysis of variance, as described in Methods, is displayed only for comparison between SD and RD vaccine arms; difference between placebo and both vaccine arms was highly significant (P=<0.0001) at all post vaccination visits. (**E**-**F**) Neutralizing titers at d36 visit in a subset of 100 subjects in SD or RD vaccine study arms, measured by SARS-CoV-2 pseudoparticle neutralization assay (PNA) based on VSV pseudotyped with Spike from Wuhan (filled symbols) or delta (open symbols) strains. Each symbol corresponds to one serum sample, horizontal line and error bars represent GM and 95% CI. Two-tailed Mann-Whitney test was used, and the only significant difference is shown in panel E. In (**E**) data are expressed as 50% neutralization titer, while in (**F**) and for Wuhan strain only as appropriate, data are converted in international units (IU)/ml. Dashed lines indicate assay LOD. In all panels, gray symbols/lines indicate placebo arm, while red and blue symbols/lines indicate single dose (SD) and repeated dose (RD) GRAd-COV2 arms, respectively.

**Table 2.** Antibody response to Spike and neutralizing antibodies to live SARS-CoV-2 at baseline and post-vaccination.

| Visit | Statistic | Anti-S Elisa (AU/ml) |  |  | SARS-CoV-2 neutralization (NT<sub>50</sub>) |  |  | SARS-CoV-2 neutralization (NT<sub>80</sub>) |  |  |
| --- | --- | --- | --- | --- | --- | --- | --- | --- | --- | --- |
|  |  | SD (N=298) | RD (N=300) | PL (N=301) | SD (N=298) | RD (N=300) | PL (N=301) | SD (N=298) | RD (N=300) | PL (N=301) |
| **Day 1** | n | 298 | 300 | 301 | 298 | 297 | 300 | 298 | 299 | 301 |
|  | GM | **1.73** | **1.77** | **1.71** | **5.35** | **5.35** | **5.31** | **4.15** | **4.16** | **4.11** |
|  | 95% CI for GM | 1.66,1.80 | 1.68,1.87 | 1.64,1.78 | 4.99,5.74 | 5.01,5.72 | 4.98,5.67 | 4.02,4.28 | 4.05,4.27 | 4.01,4.21 |
| **Day 22** | n | 291 | 295 | 288 | 291 | 294 | 288 | 291 | 295 | 288 |
|  | GM | **40.54** | **41.13** | **1.71** | **45.29** | **47.34** | **4.91** | **14.70** | **15.09** | **4.09** |
|  | 95% CI for GM | 35.62,46.14 | 36.25,46.67 | 1.64,1.79 | 38.83,52.82 | 40.50,55.35 | 4.63,5.20 | 12.84,16.82 | 13.07,17.41 | 3.99,4.20 |
|  | GMFR | 23.63 | 23.17 | 1.01 | 8.64 | 8.84 | 0.95 | 3.58 | 3.63 | 1.00 |
|  | Seroresponse (%) | 93.5 | 92.5 | 0 | 73.2 | 74.8 | 0 | 46.7 | 43.1 | 0 |
| **Day 36** | n | 287 | 293 | 279 | 286 | 292 | 278 | 287 | 292 | 278 |
|  | GM | **45.15** | **77.94** | **1.73** | **47.88** | **86.11** | **5.09** | **17.31** | **29.91** | **4.12** |
|  | 95% CI for GM | 39.54,51.56 | 70.36,86.33 | 1.65,1.81 | 41.22,55.60 | 75.43,98.30 | 4.77,5.43 | 15.18,19.73 | 26.31,34.01 | 4.02,4.23 |
|  | GMFR | 26.29 | 43.87 | 1.02 | 9.09 | 16.08 | 0.98 | 4.21 | 7.21 | 1.01 |
|  | Seroresponse (%) | 92 | 99.3 | 0 | 76.2 | 90.1 | 0.4 | 53 | 76 | 0 |
| **Day 57** | n | 279 | 288 | 232 | 279 | 288 | 232 | 279 | 288 | 232 |
|  | GM | **35.88** | **55.74** | **1.80** | **48.36** | **80.91** | **5.08** | **19.31** | **30.37** | **4.19** |
|  | 95% CI for GM | 31.23,41.22 | 50.11,62.01 | 1.68,1.94 | 40.75,57.38 | 70.19,93.26 | 4.71,5.48 | 16.66,22.39 | 26.48,34.84 | 4.02,4.37 |
|  | GMFR | 20.85 | 31.32 | 1.07 | 9.17 | 15.13 | 0.99 | 4.69 | 7.28 | 1.02 |
|  | Seroresponse (%) | 90.3 | 97.9 | 1.3 | 72 | 87.5 | 1.7 | 54.8 | 75.7 | 0.9 |
| **Day 180** | n | 193 | 238 |  | 194 | 235 |  | 194 | 235 |  |
|  | GM | **13.40** | **15.50** |  | **26.94** | **31.51** |  | **11.67** | **12.95** |  |
|  | 95% CI for GM | 11.45,15.68 | 13.56,17.73 |  | 22.16,32.75 | 26.50,37.48 |  | 9.87,13.80 | 11.17,15.02 |  |
|  | GMFR | 7.75 | 8.70 |  | 5.02 | 5.85 |  | 2.83 | 3.10 |  |
|  | Seroresponse (%) | 74.6 | 82.4 |  | 59.8 | 62.6 |  | 35.6 | 39.6 |  |

**Notes:**
N = Number of volunteers in the IAS population. n = Number of volunteers in the IAS population with available data at each timepoint. GM = Geometric Mean.
GMFR = GM fold rise. SD=single dose. RD=repeated dose. PL=placebo. The 95% CIs were calculated based on the t-distribution of the natural log transformed values then back transformed to the original scale.
Data from LABCORP and VIROCLINICS-DDL Central Laboratory. Seroresponse is defined as $\geq 4$ -fold rise in titres post-baseline.

Assessment of RBD-binding IgG (Figure 4B) provided substantially similar results in terms of IgG kinetics for the SD and RD vaccine regimens, with clear effect of a GRAd-COV2 second dose (GMT 942 and 1959 AU/ml at d36, 892 and 1602 AU/ml at d57 in SD and RD respectively, P=0.0001 at both time points). Decline of RBD IgG titers to similar levels at 6 months (d180) was also confirmed (253 and 330 AU/ml in SD and RD, respectively).

Upon conversion into WHO Binding antibody units (BAU)/ml (Supplemental Figure 1A-B), a single administration of GRAd-COV2 provided peak (d36) GM titers of 212 and 134 BAU/ml on Spike and RBD respectively in SD arm, and of 366 and 278 BAU/ml on Spike and RBD respectively after a second GRAd-COV2 dose in RD arm.

As measured in a live SARS-CoV-2 neutralization assay (Figure 4C and Table 2), neutralizing antibodies were induced (GM NT50 of 45.29 and 47.88 at d22 and d36 respectively) and remained stable until d57 in SD arm (GM NT50 of 48.36 at d57), with peak seroconversion rate of 76.2% at d36. A second GRAd-COV2 administration raised neutralizing titers from GM 47.34 (d22) to 86.11 NT50 at d36 in RD arm, with seroconversion rate reaching 90.1%. Titers remained stable up to d57, to then decline at 6 months reaching similar levels in both arms (GMT around 30 NT50 at d180), a 1.8- and 2.7-fold contraction from peak in SD and RD arms respectively. Expressed as NT80, peak neutralization GMT of 19.31 (SD) and 30.37 (RD) were reached at d57 in both study arms, with maximal seroconversion rates of 54.8% (d57) for SD and 76% (d36) in RD, again confirming the positive effect of a second GRAd-COV2 administration (Figure 4D and Table 2).

A representative sample of d36 sera from 100 subjects per vaccine arm tested with Spike pseudotyped VSV (Figure 4E) returned 50% neutralization titer (NT50) GM of 105.8 and 163.1 for SD and RD arms respectively on Wuhan strain. A 4-fold loss in neutralization potency was observed on delta variant (GM NT50 of 26.52 and 40.58), with the RD regimen resulting in a lower frequency of subjects with undetectable delta neutralizing titer (12% vs 21% in RD and SD). When expressed in WHO international units (Figure 4F), neutralizing titers on Wuhan strain were at 56.58 and 87.15 IU50/ml in SD and RD arms. A subset of d36 sera from 10 subjects per study arm was finally tested with a third confirmatory lentivirus-based pseudoneutralization assay, providing comparable NT50 titers to those seen with the other two neutralization assays. Reassuringly, all serology assays used to assess binding and neutralizing antibodies yielded highly correlated datasets (Supplemental Figure 2A-C).

Only minor differences in vaccine immunogenicity were attributable to age or co-morbidities (Supplemental Figure 3A, 3B, 3D, 3E). Older age volunteers in RD arm receiving the lower vaccine dosage (1×10^11^ vp) responded less vigorously than younger healthy volunteers throughout all time points, but GMT increased adequately after second dose, highlighting the benefit of a second GRAd-COV2 administration in this age cohort. Conversely, immune responses were comparable in the two age cohorts at the higher vaccine dose (2×10^11^ vp, SD arm). A trend for stronger immune response to both SD and RD vaccination regimens was also noted in females (Supplemental Figure 3C and 3F).

In a subset of volunteers with negative anti-N but detectable Spike binding antibodies at baseline, Spike binding and neutralizing antibodies reached 10-fold higher levels at d22 post GRAd-COV2 administration compared to seronegative volunteers, with no appreciable boosting effect of the second vaccine dose (Supplemental Figure 4A-D). The study also enrolled a small set of HIV-infected volunteers who responded with similar antibody titers magnitude and kinetics to both GRAd-COV2 SD and RD vaccination regimens compared to uninfected subjects (Supplemental Figure 4E-H). A 10 to 20-fold higher level of Spike binding and neutralizing antibodies were found at d180 visit in study participants that received approved (mostly mRNA) COVID-19 vaccines after unblinding at d57. Such titers were clearly higher than at peak with either SD or RD primary series GRAd-COV2 vaccination (Supplemental Figure 4I-L). Similarly, intercurrent SARS-CoV-2 infection or exposure enhanced or boosted both binding and neutralizing antibodies on top of GRAd-COV2 immunogenicity at the time when intercurrent exposure/infection was detected (Supplemental Figure 5A-F).

### T cell response

Potent Spike-specific T cell response were detected by IFNγ ELISpot 3 weeks after a single GRAd-COV2 vaccination (Figure 5A, d22), with GM of 1438 and 920 spot forming cells (SFC) per million PBMC in the SD and RD arm respectively. Administration of a second GRAd-COV2 dose did not result in increased T cell response, that remained otherwise stable at d36 in both SD and RD arms (GM 1515 and 1020 SFC/million PBMC respectively). Low to moderate levels of IFNγ secretion in response to Spike stimulation was also detectable in a subset of subjects in the placebo arm. Despite the trend for slightly higher responses in the SD arm, possibly due to the 2-fold higher vaccine dose, there was not statistically significant difference between SD and RD at both d22 and d36 visits, while the difference between each vaccine arm and Placebo arm was strongly significant (P=<0.0001 by Mann-Whitney at both visits). The T cell response was broad and targeted to epitopes sparse onto the whole Spike protein, with some preferential recognition of peptide pool S1b that included the RBD region (Figure 5B). Importantly, the vaccine-induced T cell response to Spike from the delta and omicron variants was mostly conserved (Figure 5C). As seen in phase 1 trial ^11^, the T cell response was Th1 skewed with virtually absent IL5 secretion in response to Spike antigen stimulation (Figure 5D).

**Figure 5.**
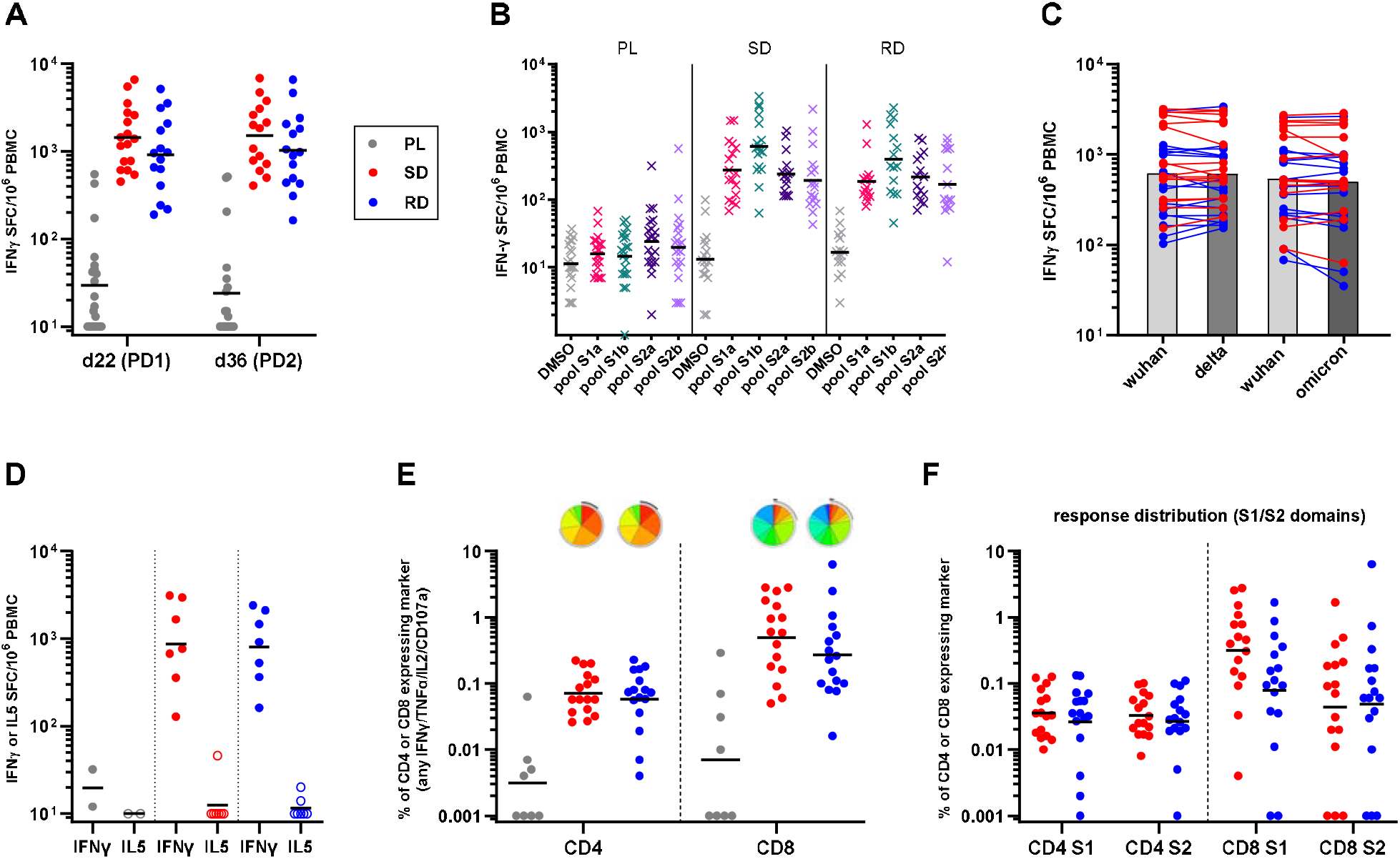
Spike specific T cell response after GRAd-COV2 vaccination. PBMC were isolated and cryopreserved for the analysis of T cell responses from a subset of 54 volunteers, 21 enrolled in placebo (PL), 17 in SD and 16 in RD arms. (**A**) Total T cell response to SARS-CoV-2 Spike at day 22 (post dose 1-PD1) and day 36 (post dose 2-PD2), evaluated by IFNγ ELISpot and expressed as IFNγ spot-forming cells per million PBMC. (**B**) Breadth of response to Spike: response to DMSO (negative control-grey symbols) and peptide pools covering the S1a (pink symbols), S1b (green symbols), S2a (purple symbols) and S2b (violet symbols) portion of Spike, evaluated in ELISpot at day 22. (**C**) Cross-reactivity of the T cell response to variants of concern: total Spike response to Wuhan and Delta or Wuhan and Omicron variants, evaluated in distinct ELISpot assays using day 22 PBMC from all GRAd-COV2 vaccinated subjects. The response on each variant in an individual volunteer is connected by a line, and bars are set at geometric mean. (**D**) IFNγ (Th1) and IL-5 (Th2) production upon Spike peptide pools stimulation, evaluated at day 36 visit in eight subjects per vaccine arm and two placebo recipients by two-color ELISpot. Multiparametric flow cytometry analysis of CD4 and CD8 T cells responses at day 36 in all GRAd-COV2 and eight placebo recipients: (**E**) Total Spike response and (**F**) breadth of response on S1 and S2 spike domains. Data are expressed as the percentage CD4 and CD8 expressing any combination of the analyzed functions (IFNγ, TNFα, IL-2 or CD107a) within CD69+ fraction in response to Spike antigen stimulation. Pie charts (base: median) representing the functional profile of Spike-specific CD4 and CD8 are shown in panel **E** and are better described in Supplemental Figure 7. In all panels, red circles represent SD arm subjects, blue circles represent RD arm subjects, grey circles represent placebo (PL) arm subjects, and black horizontal lines indicate geometric mean.

Multiparameter flow cytometry analysis conducted at d36 (Figure 5E and Supplemental Figure 6A-B) showed that GRAd-COV2 vaccination induced Spike-specific T cell response composed of CD4 (GM 0.071% and 0.058% in SD and RD arms), and even higher frequency of CD8 (GM 0.5% and 0.27% in SD and RD respectively). Polyfunctionality analysis of CD4 compartment showed that subsets of Spike-specific cells expressing all three cytokines (IFNγ/TNFα/IL2) as well as any combinations of two cytokines or IFNγ only were present at similar levels (Figure 5E and Supplemental Figure 7A), while in the Spike-specific CD8 compartment the most represented subsets were dual IFNγ/CD107a, dual IFNγ/TNFα or IFNγ only, with fair presence of triple IFNγ/TNFα/CD107a (Figure 5E and Supplemental Figure 7B). Overall, around one half of the Spike-specific CD8 showed degranulation/cytotoxic potential (CD107a+). The distribution of the different (poly)functional cell subsets was virtually identical in the SD and RD arm. The level of Spike-specific CD154+/CD69+ CD4 was superimposable to Th1 CD4 response as detected by any combination of IFNγ/TNFα/IL2 secretion, or to CD4 cells secreting IFNγ only; this observation confirms that contribution of non-Th1 (i.e. Th2, Th17) to Spike specific CD4 response to GRAd-COV2 is negligible (Supplemental Figure 8A-B). Spike specific CD4 and CD8 recognized both S1 and S2 Spike domains, highlighting the breadth of responses in both T cell subsets (Figure 5F).

### Spike-specific T and B cell memory 6 months after vaccination

High frequency of circulating Spike-specific T cells readily secreting IFNγ upon antigen stimulation were found in all volunteers in RD and SD arms 6 months (d180) after the first GRAd-COV2 vaccination (Figure 6A), at levels only minimally reduced compared to those measured at peak (GM ratio d36/d180 of 1.9 in RD and 2.3 in SD subjects with response at both time points evaluable). Receipt of approved COVID-19 vaccines after d57 stabilized T cell responses (GM ratio d36/180 of 1.1 in SD+vax subjects), but did not further amplify them above peak levels achieved with GRAd-COV2 vaccination, at variance with the potent boosting of binding and neutralizing antibodies in the same subjects (Supplemental Figure 9A-B).

**Figure 6.**
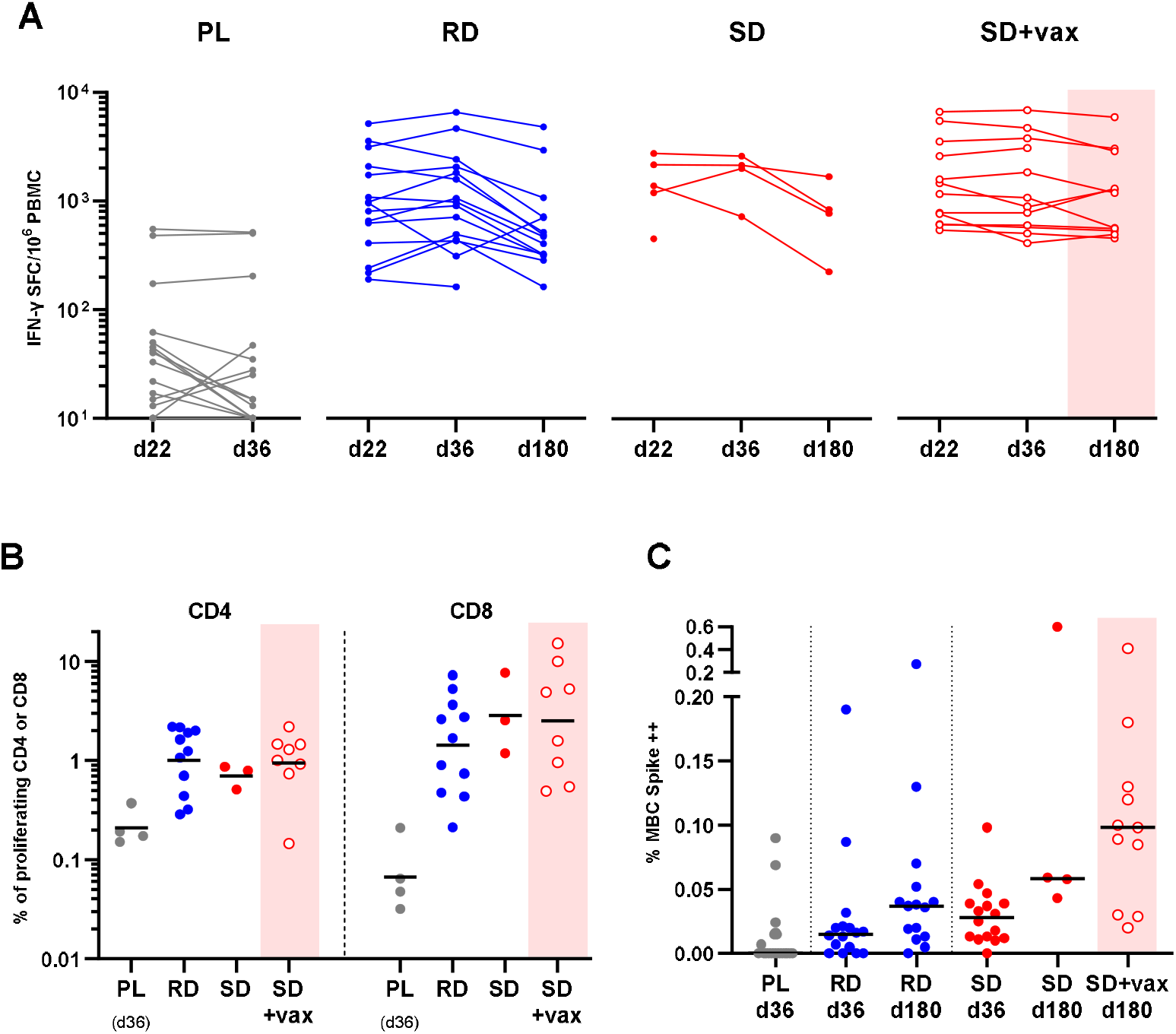
Long term T and B memory response in the PBMC sub-study. Kinetics of T cell response to SARS-CoV-2 Spike at day 22 (post dose 1-PD1), day 36 (post dose 2-PD2) and d180, evaluated by IFNγ ELISpot and expressed as IFNγ spot-forming cells per million PBMC. proliferative CD4 and CD8 T cell responses (% CD4 or CD8 CellTrace low) following 5 days incubation with Spike peptide pools, assessed in PBMC collected at d180 (d36 for the four Placebo arm subjects). (**C**) quantification of Spike-specific memory B cells (MBC) percentages in all 54 PBMC sub-study subjects at d36 and d180 visits. In all panels, gray symbols/lines indicate placebo arm, while red and blue symbols/lines indicate single dose (SD) and repeated dose (RD) GRAd-COV2 arms, respectively. Open red symbols and pink shaded areas indicate volunteers in the SD cohort that received an approved COVID-19 vaccine between d57 and d180 visit. Horizontal black lines are set at geometric mean.

GRAd-COV2-induced Spike-specific T cells had strong proliferative capacity (Figure 6B and Supplemental Figure 10) 6 months post-vaccination, with frequencies as high as GM 1% for CD4 and 2-3% for CD8. A booster dose of approved COVID-19 vaccines did not enhance further the proliferating memory T cells pool, in agreement with the ELISpot data. Proliferative CD4 and CD8 response were detectable on both S1 and S2 Spike domains, again highlighting the breadth of the GRAd-COV2 induced memory T cell response (Supplemental Figure 11).

Long-lived Spike specific memory B cells were generated by GRAd-COV2 vaccination (Figure 6C and Supplemental Figure 12) with no major differences between SD or RD regimes. If any, the higher levels seen at d36 in SD regimen may relate to the higher vaccine dose received by the volunteers at priming. The memory B cell frequency increased from d36 to d180 (P=0.035 for RD by two tailed, paired Wilcoxon test). Coherently with the efficient boosting of antibody responses by approved COVID-19 vaccines, the pool of memory B cells was also clearly amplified in most volunteers receiving heterologous vaccination regimes (P=0.002 for SD+vax by two tailed, paired Wilcoxon test).

## Discussion

The first randomised controlled trial of GRAd-COV2, given in a single dose or a two-dose regimen has shown that the novel gorilla adenovirus-based vaccine is well tolerated and immunogenic in healthy adults and in at-risk individuals, with no related SAE. The administration of a second dose was better tolerated as previously reported for two-doses adenoviral vectored vaccines regimens (15). Humoral and cellular immune responses were induced in the majority of vaccine recipients after a single immunization, with anti-S antibodies doubling two weeks after the second shot. The vaccine was better tolerated and only slightly less immunogenic in older adults, the population that benefits most from a higher vaccine dose (14) and second dose administration. We also report adequate immune response in a small number of persons leaving with HIV, similarly to what was reported for Vaxzevria (16).

Importantly, no thrombotic event was recorded in COVITAR participants. A recent FDA release estimates the risk of vaccine induced thrombosis with thrombocytopenia (VITT) at 3.23 events per million of administered doses for Jcovden, (17) while estimates are higher for Vaxzevria, between 1 case per 26,500 to 1 case per 127,300 first doses (18). Given the low frequencies reported for this rare but serious event with other adenoviral platform-based vaccines, only implementation in clinical practice may reveal if and to what extent GRAd vector is prone to induce the syndrome.

The introduction in January 2021 of the first WHO International Standard for anti-SARS-CoV-2 immunoglobulin has been instrumental for calibration of different SARS-CoV-2 serological assays and for the expression of data in a common unitage, allowing the comparison of immunogenicity dataset associated to different COVID-19 vaccines. Reported peak IgG concentrations against the prototype Wuhan Spike were in the range of 60-100 BAU/ml after a single administration of Jcovden or Vaxzevria, increasing to around 200-500 depending on assay, antigen (Spike or RBD) and dosing interval for homologous double dose regimens (19-23). Here we show that a single administration of GRAd-COV2 induces peak binding antibody levels in the range of 150-250 BAU/ml, reaching 250-400 on RBD and Spike respectively with a second vaccine dose; similarly, in a subset of GRAd-COV2 recipients, peak neutralizing antibody titers as detected by PNA was around 60 (SD) and 80 (RD) IU_50_/ml, well in agreement with levels reported for Jcovden or Vaxzevria; in turn this would predict similar vaccine efficacy (19, 24). However, this remains to be formally proven in a phase 3 study.

It is highly improbable that substantial cross-neutralization on omicron would be detected in serum of GRAd-COV2 vaccinated subjects, as suggested by the moderate nAb titers on ancestral, and the clear reduction on delta variant as detected by PNA assay. However, this is the case for most vaccines after primary series, and multiple boosting strategy or bivalent vaccines is indeed the current standard for vaccination campaigns. Of note, extending the interval between first and second GRAd-COV2 administration beyond the 3 weeks explored in this study is expected to further increase immunogenicity, as now clearly established for both adenoviral vectors and mRNA-based vaccines (25-30).

Although not formally assessed as correlate of protection for SARS-CoV-2 infection, vaccine induced T cell responses may ensure long-term protection from severe disease also thanks to their broad cross-reactivity against all circulating VOC (14, 31, 32). Here we show that GRAd-COV2 vaccine induces potent and broad Spike-specific Th1 skewed cellular response following the first dose. Administration of a second GRAd-COV2 dose do not increase the magnitude or alter the polyfunctionality profile of the T cell response, at least with a short 3 weeks interval, similarly to other adeno-based vaccines (15). Polyfunctional CD4 responses induced by GRAd-COV2 are well in range to those reported for other COVID-19 vaccines, while frequencies of CD8 T cells are remarkable, placing GRAd-COV2 as potential best in class for CD8 responses (33). This finding may be attributed to the slightly higher vaccine dosage than other adenoviral-based vaccines, but is also well aligned with previous experience indicating group C adenoviral vectors as the most potent inducers of T cell responses (34). As expected, T cell response cross-recognized Spike from both delta and omicron suggesting high potential for cross-recognition of even highly divergent variants. Importantly, vaccine induced T cells retained immediate effector functions as well as proliferative capacity up to 6 months after immunization demonstrating the establishment of long-lasting and proficient antigen-specific memory T cells that is a hallmark of successful vaccination. Moreover, GRAd-COV2 induced Spike-specific memory B cells that increase over time, as reported for other COVID-19 vaccines (33, 35, 36).

The study has several limitations. First, the epidemiological context and the national vaccine campaign leading to study unblinding at day 57 has made it impossible to transition to phase 3 part of the study and to assess the vaccine efficacy, as originally planned in the study protocol. Nevertheless, the blinded phase of the study has allowed to rigorously evaluate the safety of single and repeated dose regimens, and has enabled the study Steering Committee jointly with DSMB to recommend the two-dose regimen for further clinical development. Second, the study was conducted in Italian clinical centers only, with population ethnicity limited to white-Caucasian subjects. Further studies are needed to ascertain the vaccine safety and immunogenicity in more diverse populations. Third, the durability of the immune response can only be assessed up to the presently reported 6 months follow up; in fact, within d180 and d360 visits, almost all subjects enrolled in SD and RD arms have received COVID approved vaccine as booster doses. The dataset at d360 will anyway provide further interesting immunogenicity data on the combination of GRAd-COV2 with existing approved (mostly mRNA-based) COVID-19 vaccines. Fourth, since omicron was not yet circulating at the time of our study, it was not included in the pre-planned serology panel.

Although the implementation of this vaccine for primary course is of limited or no value in light of omicron prevalence, GRAd-COV2 could be deployed as a component of highly immunogenic heterologous prime-boost regimens as it has been shown with other mRNA/Ad-based vaccines (19, 21, 22, 37) and incidentally demonstrated in the GRAd-COV2 phase 1 trial (38) and in the current report. While it is now well established that an mRNA booster dose provides best peak immunogenicity in the short term, recent evidence suggests that adenoviral vectored vaccines may be a sound option as booster doses in mRNA primary cycle recipients, in light of slower antibody decay rates and superior T cell boost, especially of (VOC crossreactive) CD8 (39, 40). Indeed, GRAd-COV2 is based on a different non cross-reacting adenoviral serotype from all approved adenovirus-based vaccines already deployed in worldwide vaccination campaigns; this may represent an advantage for future booster doses thanks to absent or low preexisting anti-GRAd immunity in the human population. Together with the favorable “tractability” typical of the platform, i.e. low cost of goods, easier manufacturing process tech-transfer allowing local production and reasonable thermostability, GRAd-COV2 is an attractive vaccine option especially in lower income countries.

In conclusion, we propose that the novel GRAd adenoviral vector is an attractive novel vaccine platform; simply by changing the antigenic load, the platform is suitable for developing effective 2nd generation COVID-19 or pan-(sarbe)coronavirus vaccines or for any new emerging pathogens requiring the generation of well-coordinated antibody and T cell response. Specifically, the induction of a potent and durable CD8 T cell response by the GRAd might be instrumental to fight SARS-CoV-2 variants, future emerging pathogens as well as diseases where this type of immune response is defective or suppressed. In addition, the adenoviral platform is suitable for mucosal administration, as shown in many preclinical and clinical settings even for COVID-19 (41, 42), which may be a highly desirable feature for the development of a vaccine with transmission-blocking potential (43).

## Methods

### Trial design and oversight

This phase 2 randomised, observer-blind, placebo controlled, multicenter trial (COVITAR study, ClinicalTrials.gov NCT04791423) is the first part of a phase 2/3 protocol study and was conducted at 24 centers in Italy in accordance with the Declaration of Helsinki and Good Clinical Practices. An independent Data Safety Monitoring Board (DSMB) was established before the start of the trial and reviewed unblinded safety data twice during the study. Around 900 adult female and male, ≥ 18 years of age were planned to be included. The main exclusion criteria included: allergy to any vaccine component, Guillain-Barré syndrome, laboratory-confirmed SARS-CoV-2 infection, immunodeficiency state, any vaccination (licensed or investigational) other than for influenza within 30 days before/after administration of study intervention, and pregnancy. Full inclusion and exclusion criteria are in the protocol. Mild/moderate well controlled comorbidities were allowed, including HIV infection. Randomisation was stratified by age (< or ≥ 65 years); for participants < 65 years, by comorbidities representing risk factors for COVID-19 severe illness (per CDC recommendation, May 2020). At least 25% of enrolled participants had to be either ≥ 65 years or < 65 years and “at risk”.

By the use of an interactive Web-based system, participants were randomly assigned at a ratio of 1:1:1 to receive, 21 days apart, a single administration of GRAd-COV2 at a dose of 2×10^11^ viral particles (vp) followed by placebo, or a repeated GRAd-vaccine dose of 1×10^11^ vp, or two doses of placebo. Neither participants, nor investigators or Sponsor’s staff involved in clinical management or study monitoring were aware of the study intervention administered. Since GRAd-COV2 and placebo were visually distinct prior to dose preparation, in order to maintain blindness preparation of the syringes was done by an unblinded pharmacist, and then handed over to the investigator for administration to the participant.

### Study approval

The study protocol was approved by the Italian Regulatory Agency (AIFA), the COVID national Ethics Committee (Lazzaro Spallanzani Institute), and the local Ethics Committees of the other 23 clinical centers. Study protocol is available in Supplemental material. All participants received and signed a written informed consent prior to enrollment.

### Vaccine product and placebo

GRAd-COV2 (ΔE1, ΔE3, ΔE4) was manufactured by ReiThera srl under good manufacturing practice conditions in the proprietary cell line ReiCell35S, a suspension adapted packaging cell line derivative of HEK293. The vaccine was purified by an extensive downstream process including host cell DNA precipitation, depth filtration, two chromatographic purification steps followed by nuclease digestion and ultrafiltration. The clinical material was finally formulated in formulation buffer (10 mM Tris, 75 mM NaCl, 1 mM MgCl2, 0.02% PS80, 5% sucrose, 0.1 mM EDTA, 10 mM Histidine, 0.5% ethanol, pH 7.4) at a concentration of 2 × 10^11^ viral particles/ml. Commercially available sterile 0.9% (w/v) saline solution was administered as Placebo, and used to dilute the investigational vaccine for the 1×10^11^ vp dose.

### Trial procedures

Participants received the two injections (1 mL volume) in the deltoid muscle, and remained in observation for 30 minutes after vaccination for acute reactions.

The primary end points of this trial were: solicited local or systemic adverse events (AEs) within 7 days after each dose of vaccine or placebo, recorded through an electronic diary; unsolicited AEs reported by participants through 1 month after the second dose; serious adverse events (SAEs) and adverse events of special interest (AESIs) through 1 year after the second injection. AEs data up to approximately 24 weeks after the second dose are included in this report and safety results are reported for all participants who provided informed consent and received at least one dose of vaccine or placebo.

All participants had humoral immunogenicity assessment at 1 (pre-dose), 22, 36, 57, 180 and 360 days after the first dose. A subset of participants in a single clinical site (N=54, at CRC Verona, Italy) had cellular (T and B) immune response assessment in peripheral blood mononuclear cells (PBMC) at study day 22, 36 and 180. The detailed methods for immunogenicity assays are reported in Supplemental material. Main serology assays were centralized.

Primary assessment of humoral response was conducted by a semi-quantitative ELISA on full length Spike, using COVID-SeroIndex, Kantaro Quantitative SARS-CoV-2 IgG Antibody IVD Kit (R&D Systems). SARS-CoV-2 infections were monitored by testing sera for seroconversion to Nucleocapsid (N) antigen using SARS-CoV-2 IgG kit, a chemiluminescence microparticle assay (CMIA) on ARCHITECT platform (Abbott Diagnostics, Chicago, IL, USA). Both Spike ELISA and N CMIA were run at centralized lab (LabCorp, Geneva). SARS-CoV-2 neutralizing antibodies were measured by a plaque reduction neutralization test (Viroclinics-DDL, the Netherlands) using SARS-CoV-2 Bav/Pat1/2020 strain and determining 50% and 80% neutralization titers (NT_50_ and NT_80_). Additional exploratory serology on selected samples or study visits, as detailed in the results section, were also conducted: IgG to receptor binding domain (RBD) were measured with SARS-CoV-2 IgG II Quant, a CMIA assay on ARCHITECT platform (Abbott Diagnostics, Chicago, IL, USA); neutralization activity was assessed with two pseudovirus neutralization assays (PNA) either based on vescicular stomatitis virus (VSV) pseudotyped with SARS-CoV-2 Spike glycoproteins from Wuhan (D614) or delta strains (Nexelis Laval, QC, Canada), or by PhenoSense Anti-SARS CoV-2 Neutralizing Antibody Assay, based on D614 Spike pseudotyped lentivirus (Monogram Biosciences Inc., CA).

Cellular immune responses to Spike from prototype Wuhan, delta and omicron variants were measured primarily by IFNγ enzyme-linked Immunospot (ELISpot). Additional investigation was performed by multi-parametric flow cytometry analysis for the characterization of CD4 and CD8 T cell functional profile and proliferation capacity, and for the enumeration of Spike-specific memory B cells (MBC). All cellular assays were conducted at ReiThera or Ospedale Pediatrico Bambino Gesù (Rome).

### Statistics

All statistical analyses and data processing were performed using the SAS® System software (release 9.4). The safety analyses included all participants who received at least one dose of GRAd-COV2 or placebo. The findings are descriptive in nature and not based on formal statistical hypothesis testing. Safety analyses are presented as counts, percentages, and associated Clopper–Pearson 95% confidence intervals for local reactions, systemic events, and any adverse events after vaccination, according to preferred terms in the *Medical Dictionary for Regulatory Activities* (MedDRA), version 24.0, for each group. The immunogenicity analysis set (IAS) included all participants in the safety analysis set who had immune response assessments and no protocol deviations judged to have a potential interference with the generation or interpretation of an immune response (SARS-CoV-2 infection or commercial COVID-19 vaccination). Number of cases and geometric mean (GM) with its 95% confidence interval were calculated for anti-S ELISA, SARS-CoV-2 neutralization NT_50_ and NT_80_. The 95% CIs for geometric means were calculated based on the t-distribution of the natural log-transformed values than back transformed to the original scales for presentation. Comparisons among treatment groups were performed by means of the analysis of variance for repeated measures on natural log-transformed values of immunogenicity outcomes where treatment group, study day, treatment group by study day interaction as fixed effects, with adjustment for protocol stratification factor.

## Data Availability

All data produced in the present study are available upon reasonable request to the authors

## Author contributions

RCam, SCapo, AF, and SLa conceived and designed the trial and developed the study protocol; FMF, SM, SBor, SCar, SLC, SLe, GG, PM, AC, MLic, RCau, SDZ, MVC, AG, SR, PC, SBon, GM, MC, IM, SCapi, EP, MLib, AD, SLa were study site principal investigators; SCapo, RCar, EPM, SBa, AMC were responsible for laboratory testing and assay development; CI performed the statistical analyses; RD, FG, GP, SCo, LV contributed to the implementation of the study; RCam; SCapo, RCar, LV, SLa had full access to and verified all the data in the study and take responsibility for the integrity and accuracy of the data analysis.; RCam, SCapo, AF wrote the manuscript; all authors critically reviewed and approved the final version.

## Acknowledgments

ReiThera srl manufactured the vaccine, funded and sponsored the trial, coordinated interactions with contract Clinical Research Organization (CRO) Exom staff and regulatory authorities. The CRO took charge of trial operation to meet the required standards of the International Council for Harmonization of Technical Requirements for Pharmaceuticals for Human Use and GCP guidelines. We are grateful to all the participants who volunteered for this study, and the members of the COVITAR data and safety monitoring board and steering committee for their dedication and their diligent review of the data. We also acknowledge the contributions of the COVITAR Clinical Trial Group (see the Supplemental material); Antonella Bacchieri (statistician consultant) for data interpretation; the ReiThera GMP, QC and QA staff for manufacturing and release of the GRAd-COV2 clinical lot, and all the ReiThera colleagues not named here who contributed to the success of this trial. We also thank Abbott for supplying the SARS-CoV-2 IgG II Quant assay kits for the study. Pseudovirus neutralization assays performed by Nexelis was supported by the Global Health Discovery Collaboratory Platforms of the Bill & Melinda Gates Foundation.

