## Supplementary material for "GRAd-COV2 vaccine provides potent and durable immunity in randomised placebo-controlled phase 2 trial (COVITAR)"

#### Index

#### Supplemental tables and figures

##### Supplemental Table 1 Overview of adverse events in the safety population

| | Vaccine single dose (n=305) | Vaccine repeated dose (n=308) | Placebo (n= 304) |
|---|---|---|---|
| Subject with at least one AE | 293 (96.1%) | 299 (97.1%) | 239 (78.6%) |
| Subjects with at least one unsolicited TEAE | 51 (16.7%) | 42 (13.6%) | 43 (14.1%) |
| Subjects with at least one related unsolicited TEAE | 11 (3.6%) | 12 (3.9%) | 9 (3.0%) |
| Subjects who discontinued the study due to unsolicited TEAE | 0 | 1 (0.3%) | 0 |
| Subjects with at least one SAE | 2 (0.7%) | 3 (1.0%) | 1 (0.3%) |
| Subjects with at least one related SAE | 0 | 0 | 0 |
| Subjects with AESI | 0 | 0 | 0 |
| Subjects with at least one local solicited AE | 253 (83.0%) | 272 (88.3%) | 128 (42.1%) |
| Subjects with at least one related local solicited TEAE | 250 (82.0%) | 271 (88.0%) | 126 (41.4%) |
| Subjects who discontinued the study due to local solicited TEAE | 0 | 0 | 0 |
| Subjects with at least one systemic solicited TEAE | 271 (88.9%) | 280 (90.9%) | 214 (70.4%) |
| Subjects with at least one related systemic solicited TEAE | 268 (87.9%) | 279 (90.6%) | 210 (69.1%) |
| Subjects who discontinued the study due to systemic solicited TEAE | 0 | 0 | 0 |

Notes: Treatment Emergent Adverse Events (TEAEs) are defined as AEs that started after the first dose administration and until 28 days post each dose of study intervention. Serious Adverse Events (SAEs) were recorded from the date of signature of informed consent form through the last participant contact. Solicited TEAEs were recorded for 7 days post each dose of study intervention. Solicited TEAEs recorded after 7 days post each dose are included in the summary table. All TEAEs are considered unsolicited unless categorized as solicited TEAEs. Adverse Events of Special Interest (AESIs) were recorded from day 1, post treatment, through the last participant contact. N= number of subjects, % = percentage of subjects. Percentages are calculated on the number of subjects (n) by treatment group.

**Supplemental Figure 1**

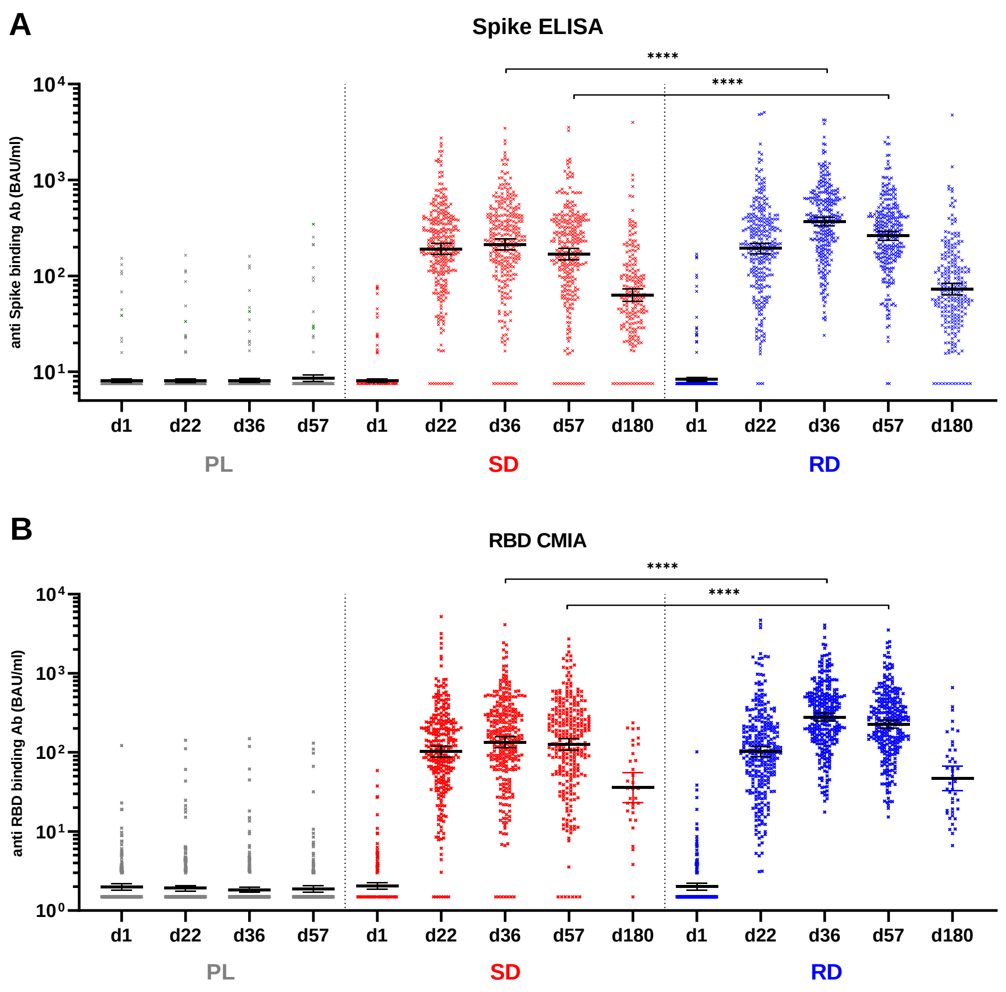

**Supplemental Figure 1. Spike and RBD binding IgG concentrations expressed in BAU/ml**

(A) trimeric Spike binding antibody concentrations, detected by ELISA (COVID-SeroKlir, Kantaro Semi-Quantitative SARS-CoV-2 IgG Antibody Kit, R&D Systems) (B) RBD binding antibody concentrations, detected by chemiluminescent microparticle immunoassay (CMIA-Abbott SARS-CoV-2 IgG II Quant assay). For both (A) and (B), the original dataset expressed in arbitrary units/ml were converted to binding international units (BAU)/ml following manufacturers indications. Symbols for each study volunteer at each study visits are shown in grey for placebo arm (PL), in red for GRAd-COV2 single dose (SD) arm and in blue for repeated dose (RD) arm. Line and error bars indicate geometric mean (GM) and 95% confidence interval (CI). Statistical analysis is displayed only for comparison between SD and RD vaccine arms; difference between placebo and both vaccine arms was highly significant (P=<0.0001) at all post vaccination visits.

**Supplemental Figure 2.**

A

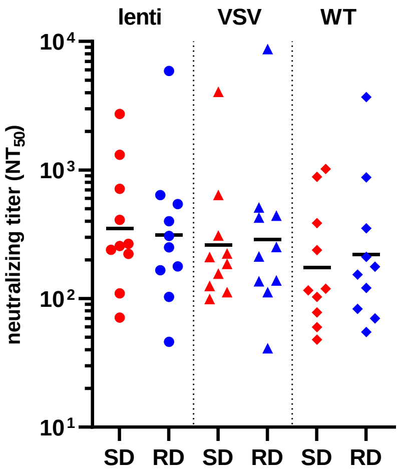

B

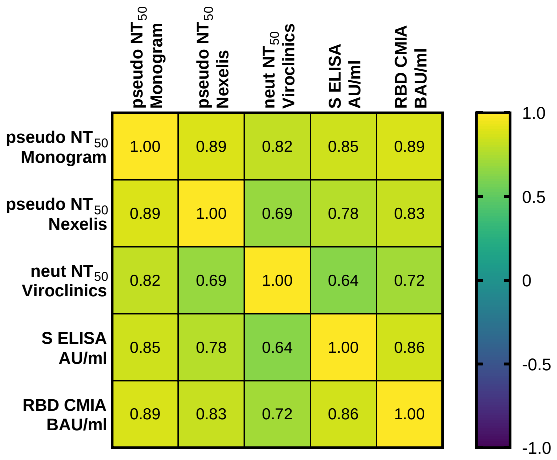

C

| | pseudo $NT_{50}$ Monogram | pseudo $NT_{50}$ Nexelis | neut $NT_{50}$ Viroclinics | S ELISA AU/ml | RBD CMIA BAU/ml |
|---|---|---|---|---|---|
| pseudo $NT_{50}$ Monogram | | 1.3466768e-007 | 0.00001 | 0.000002 | 4.5051107e-007 |
| pseudo $NT_{50}$ Nexelis | 1.3466768e-007 | | 0.00078 | 0.000046 | 0.0000100 |
| neut $NT_{50}$ Viroclinics | 0.0000105 | 0.0007802 | | 0.002199 | 0.0004720 |
| S ELISA AU/ml | 0.0000018 | 0.0000460 | 0.00220 | | 0.0000022 |
| RBD CMIA BAU/ml | 4.5051107e-007 | 0.0000100 | 0.00047 | 0.000002 | |

**Supplemental Figure 2. correlation amongst all serology assays on d36 sera from 10 participants per vaccine arm**

(A) SARS-CoV-2 50% neutralizing antibody titer ($NT_{50}$) measured in d36 serum from 20 volunteers (10 enrolled in SD and 10 in RD arms) by three distinct assays: PhenoSense Anti-SARS CoV-2 Neutralizing Antibody Assay based on D614G Spike pseudotyped lentivirus (Monogram Biosciences); SARS-CoV-2 pseudoparticle neutralization assay (PNA) based on VSV pseudotyped with D614 Spike (Nexelis); live Wuhan (Berlin strain) SARS-CoV-2 neutralization assay (Viroclinics). Horizontal black lines indicate geometric mean. (B) and (C) A correlation (non-parametric Spearman, two-tailed) matrix was computed for the 20 vaccinated volunteers, comparing the measured anti-SARS-CoV-2 binding and neutralizing antibody response to evaluate how the different immune parameters correlate each other. Spearman r values are shown in the heatmap (A), and the table (B) reports P values for each pair of variables.

#### Supplemental Figure 3

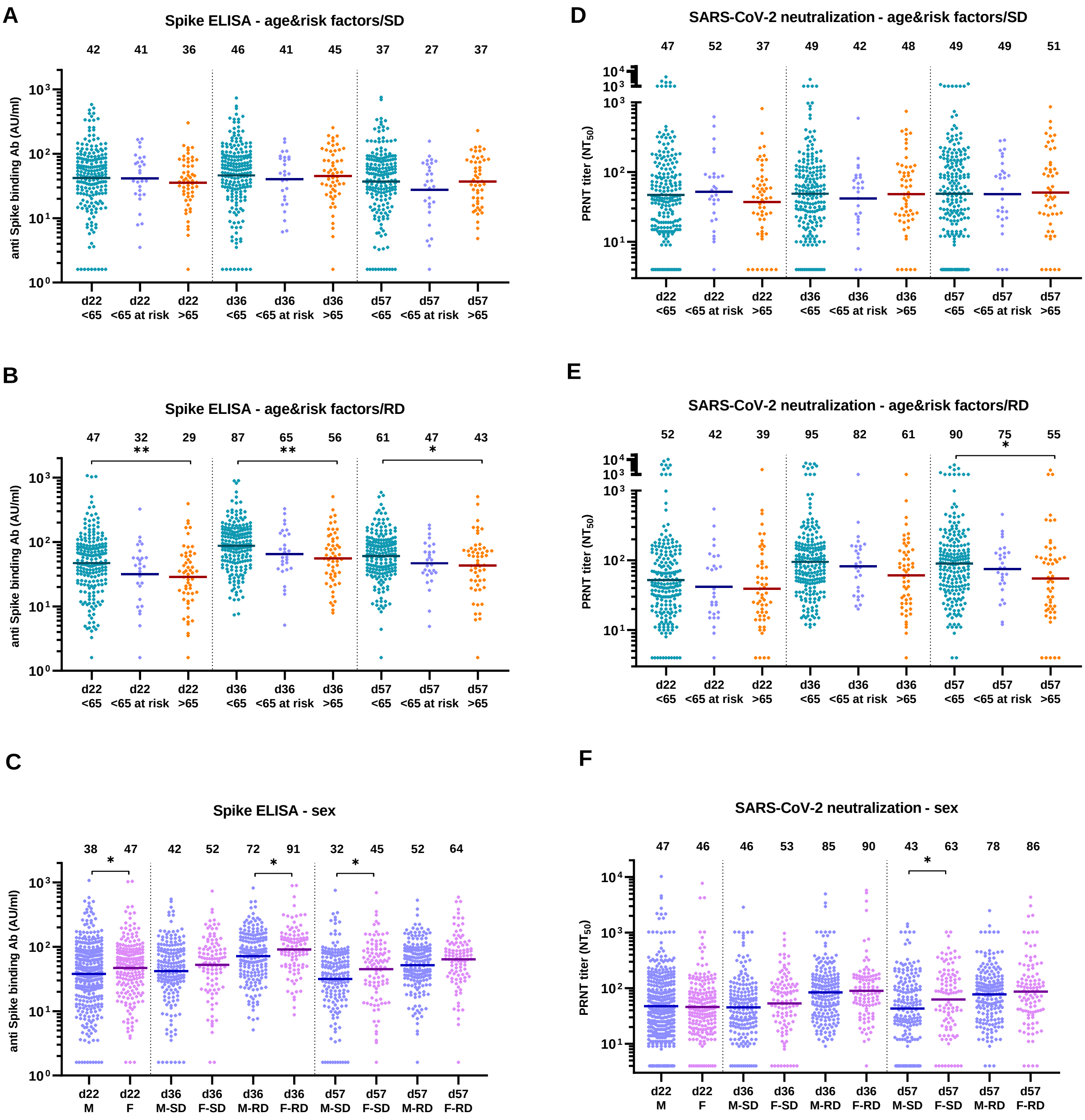

**Supplemental Figure 3. Serology analysis in study subpopulations**

Spike ELISA (panels A to C, expressed as AU/ml) and live SARS-CoV-2 neutralization (panels D to F, expressed as $NT_{50}$) dataset at visits d22, d36 and d57 are shown split by main study subpopulations. Effect of age and comorbidities on vaccine immunogenicity is shown for GRAd-COV2 SD arm (panels A and D) and for GRAd-COV2 RD arm (panels B and E), with volunteers distributed according to main per-Protocol subpopulations: age <65 years, age <65 years with comorbidities, age >65 years. Two-tailed Kruskall-Wallis test with Dunn's test correction for multiple comparisons was used to compare datasets within each study visit. Effect of sex is shown in panels C and F, with dataset split into male (M) and female (F) subjects. At visit d22, volunteers from both SD and RD arms were combined, since main statistical analysis did not detect any statistically significant difference in vaccine immunogenicity post dose 1. For d36 and d57 visits, data are shown according to study arm (SD or RD). Two tailed Mann-Whitney test was used to compare datasets within each study visit and study arm. Across all panels, horizontal lines and numbers reported above graphs indicate geometric mean. Only significant differences are displayed on the graphs.

### Supplemental Figure 4

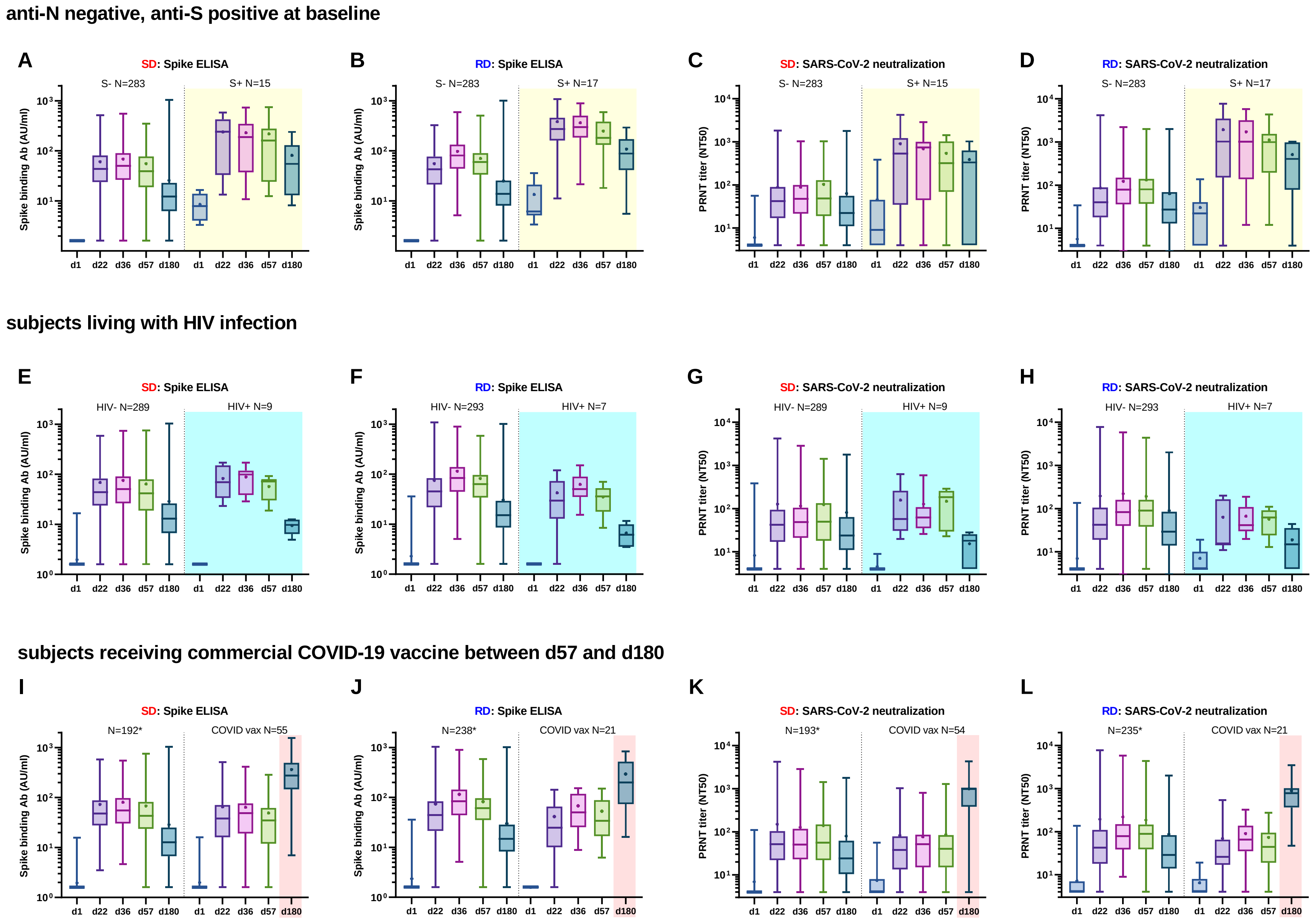

**Supplemental Figure 4. binding and neutralizing antibodies in baseline Spike seropositive, HIV infected and COVID-19 commercial vaccines recipients**

Spike ELISA (panels A-B-E-F-I-J, expressed as AU/ml) and live SARS-CoV-2 neutralization (panels C-D-G-H-K-L, expressed as $NT_{50}$) dataset at all study visits for the two GRAd-COV2 vaccine study arms (SD and RD) are shown split by population category of interest, for a descriptive analysis of vaccine immunogenicity in: subjects seronegative to SARS-CoV-2 N but seropositive (by ELISA) to S at study entry (panels A to D, yellow shaded area); subjects living with HIV infection (panels E to H, light blue shaded area); subjects that received COVID-19 approved vaccines between d57 and d180 (panels I to L, pink shaded area). Data are shown as box and whiskers plots, with whiskers extending from minimum to maximum value and mean indicated by a dot. Numerosity (N) of each subgroup at baseline is indicated for reference. For panels I to L, the asterisk on numerosity indicates that only subjects with available data at d180 visit were included.

#### Supplemental Figure 5

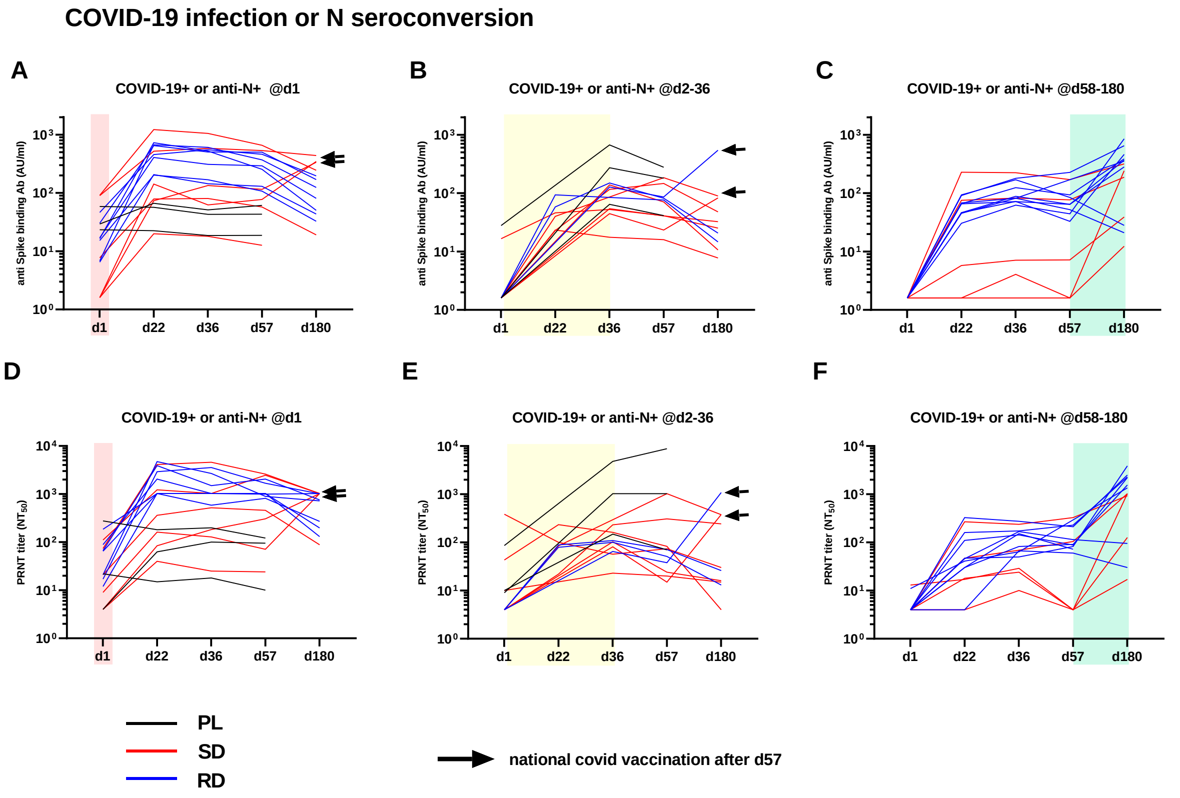

**Supplemental Figure 5. binding and neutralizing antibodies in subjects with intercurrent SARS-CoV-2 exposure or infection**

Kinetics of Spike binding antibodies (ELISA, panels A to C, expressed as AU/ml) and SARS-CoV-2 neutralizing antibodies (panels D to F, expressed as $NT_{50}$) in individual volunteers at all study visits are shown for three distinct categories: subjects found seropositive to nucleocapsid (N) antigen at study entry (panels A and D pink shaded area); subjects seroconverting to N without symptoms or with documented SARS-CoV-2 infection either during vaccination phase (between d2 and d36, panels B and E, yellow shaded area) or between d57 and d180 (panels C and F, green shaded area). The line colors indicate study arm in which the subject is enrolled: black=placebo; red=SD; blue= RD. black arrow indicates volunteers that received COVID-19 approved vaccines between d57 and d180.

**Supplemental Figure 6**

**A: Intracellular staining gating strategy**

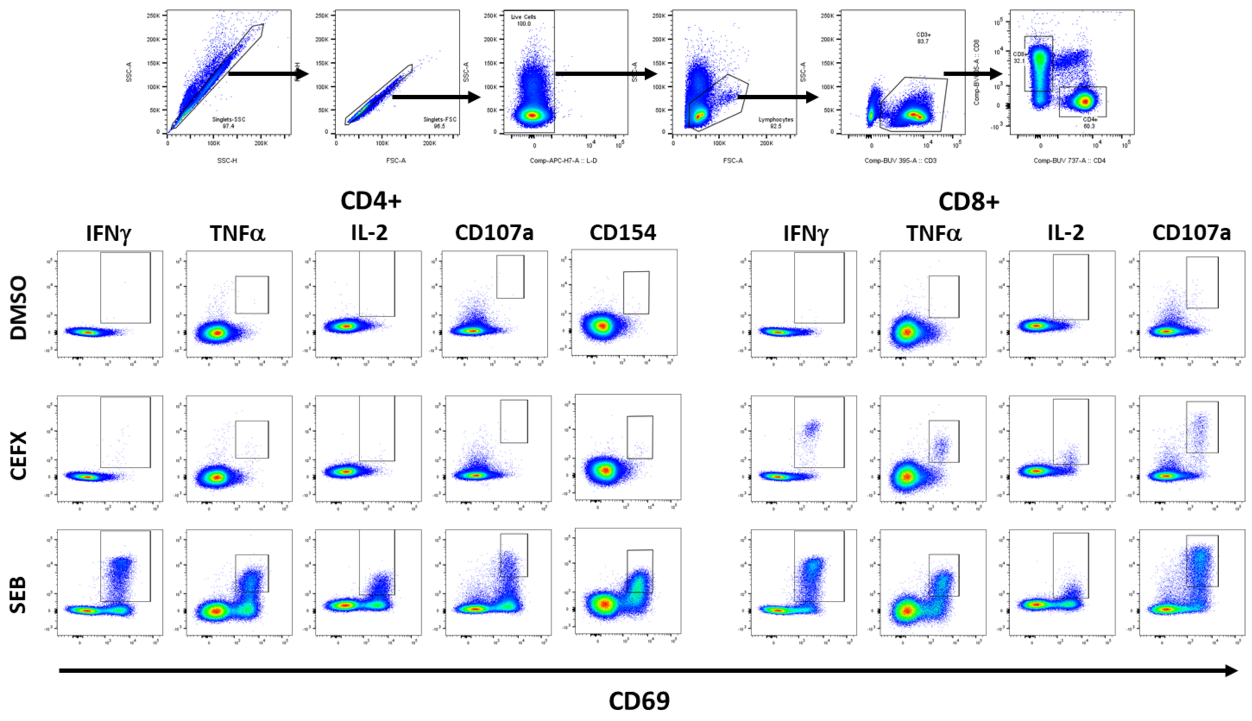

**B: Intracellular staining representative CD4 and CD8 responses to spike peptide pools in one volunteer**

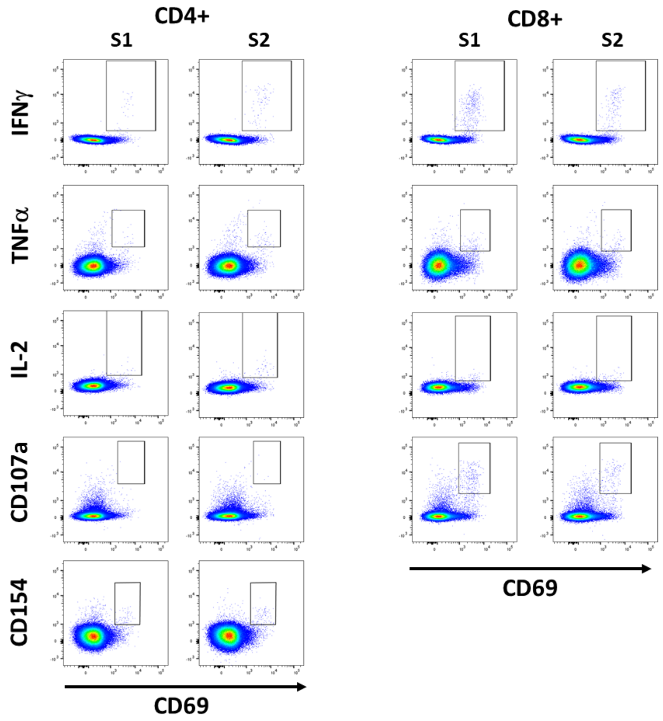

**Supplemental Figure 6. ICS gating strategy and example of Spike specific responses in a representative subject**

Representative plots for A) gating strategy of CD4+ and CD8+ T cells populations (upper plots) and analysed functions in negative control (DMSO, top lane) and positive controls (CEFX, central lane, SEB, bottom lane). B) representative plots for Spike-specific CD4+ (left) and CD8+ (right) T cell responses against S1 (first and third columns) and S2 (second and fourth columns) Spike peptide pools.

**Supplemental Figure 7**

**A**: **CD4**

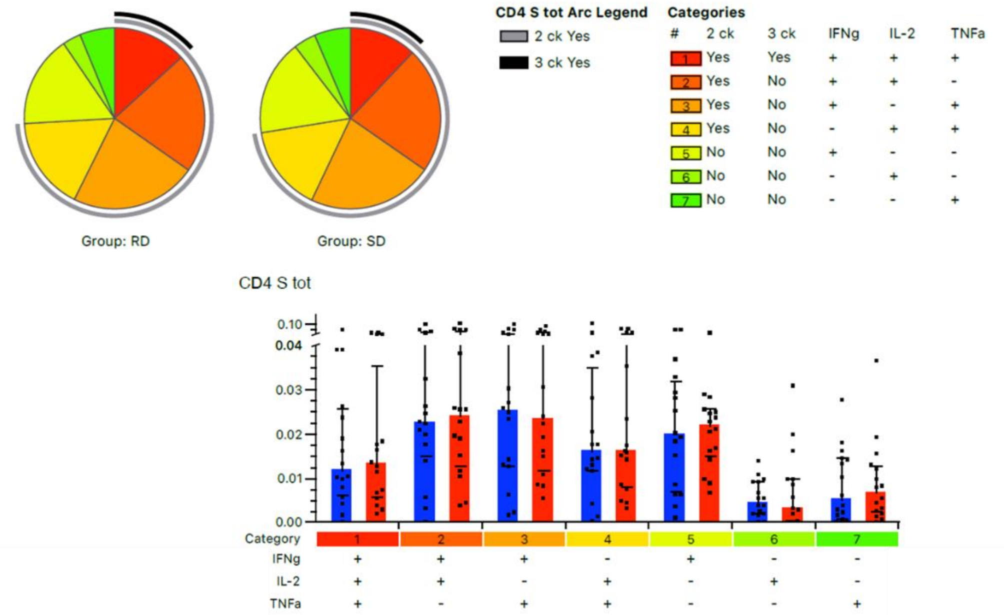

**B**: **CD8**

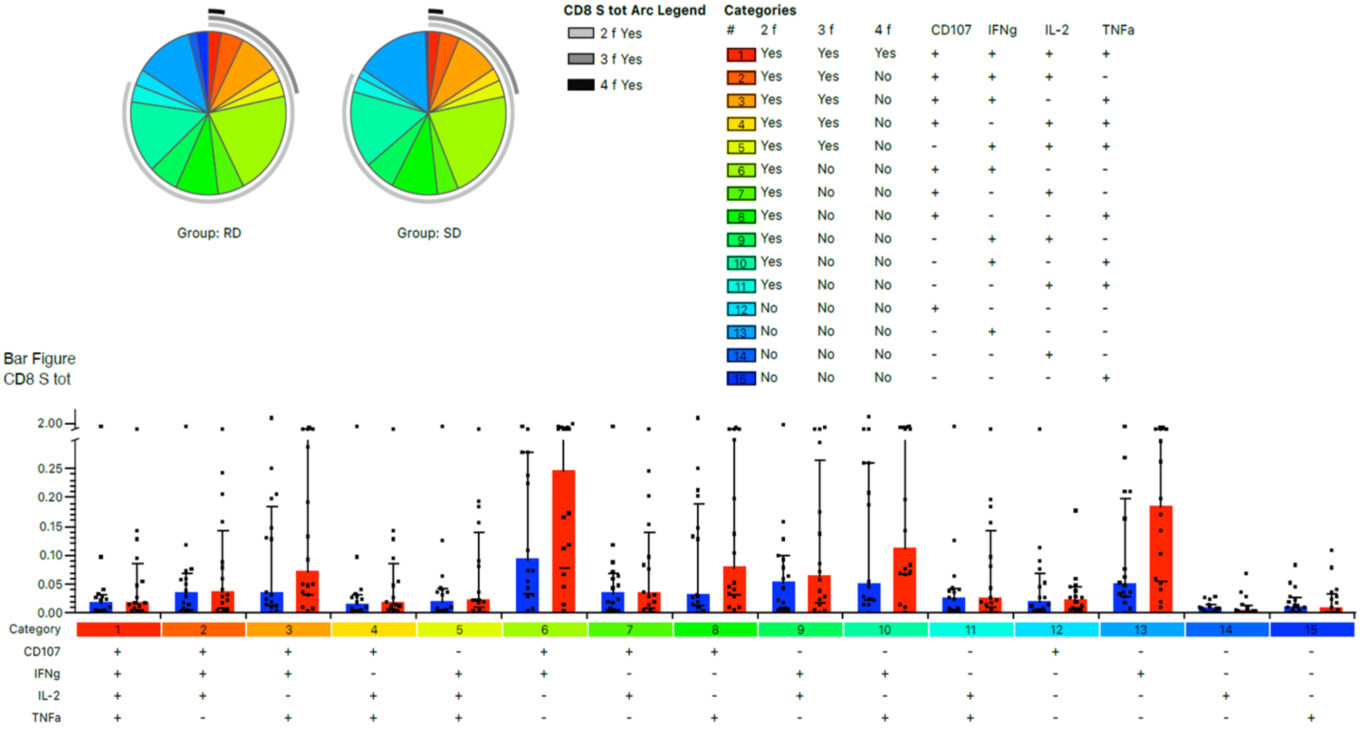

**Supplemental Figure 7: Polyfunctionality analysis**

Co-expression of 1 to 3 (IL-2, TNFα and IFNγ) and 1 to 4 functions (CD107a, IL-2, TNFα and IFNγ) was analyzed for CD4 (A) and CD8 (B) Spike-specific T cells, respectively. Pie charts (base: median) indicates relative abundance of each population (pie slices) and co-expression (arcs) while bar graphs (set at median with whiskers representing IQR) indicate their absolute percentages and distribution in RD group (blue bars) and SD group (red bars).

**Supplemental Figure 8**

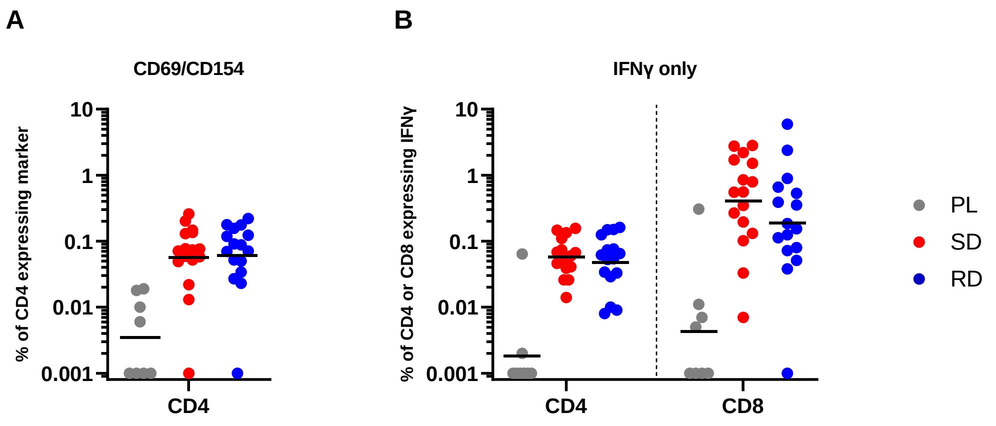

**Supplemental Figure 8. additional ICS data**

Dot plots represents the percentages of Spike-specific CD4 T cells co-expressing CD69 and CD154 activation markers (A) and the percentages of Spike-specific CD4 and CD8 expressing IFNγ (B) in the Placebo (grey dots), RD (blue dots) and SD (red dots) groups.

#### Supplemental Figure 9

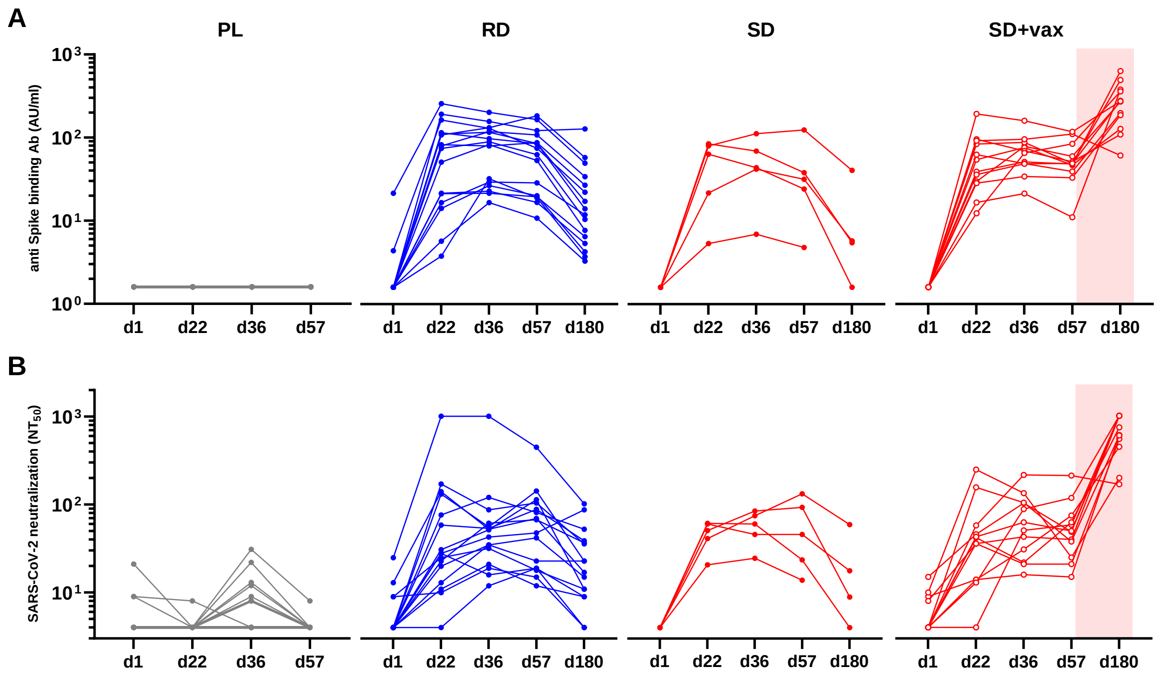

**Supplemental Figure 9. serology kinetics in PBMC sub-study volunteers**

Kinetic of Spike-specific binding (A) and neutralizing (B) antibodies in each single donor belonging to the PBMC sub-study, divided by Placebo (grey dots and lines), RD (blue dots and lines), SD (red dots and lines) and SD+vax (empty red dots and red lines, with pink shaded area indicating time frame of COVID-19 approved vaccine receipt) groups.

**Supplemental Figure 10**

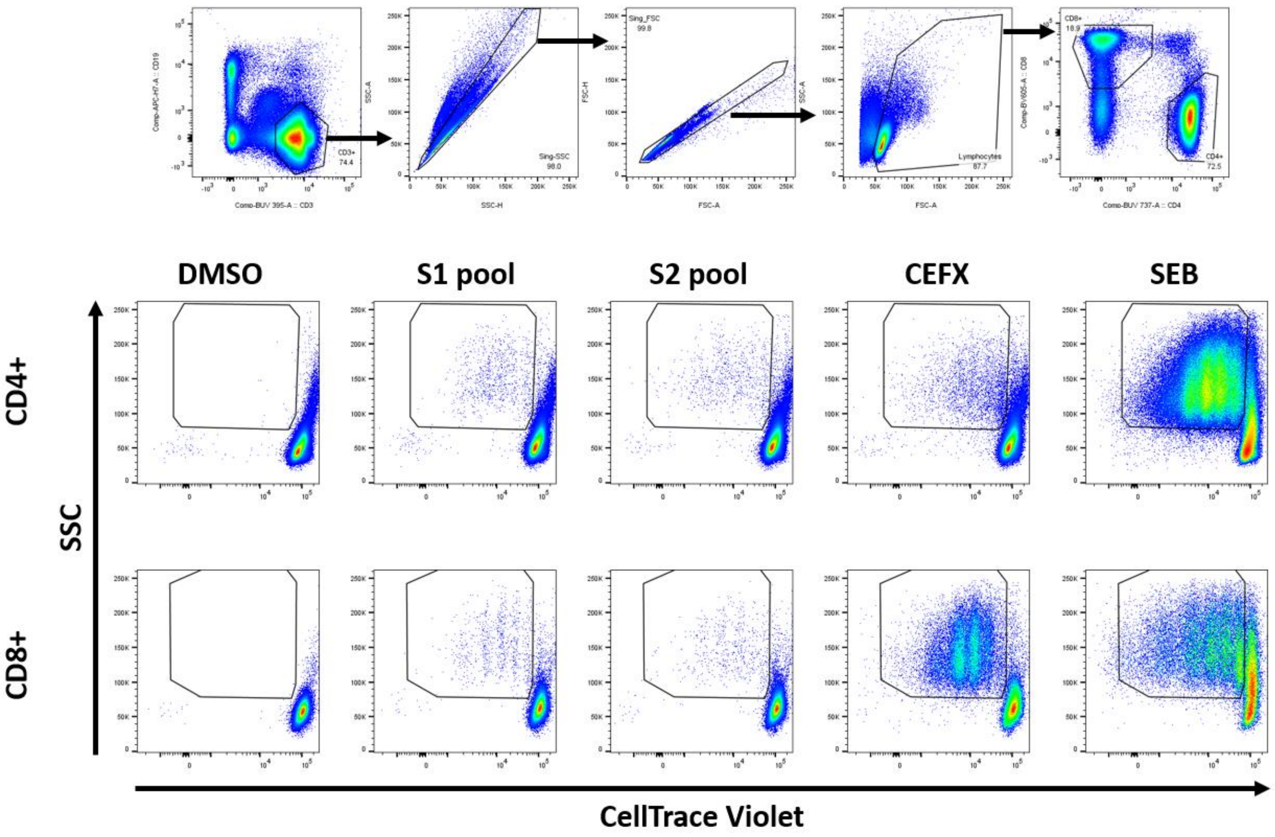

**Supplemental Figure 10. T cell proliferation assay gating strategy**
Representative plots for gating strategy (upper lane) of T cell populations and proliferative response (i.e. dilution of CellTrace dye) of CD4 (middle lane) and CD8 (lower lane) after 5 days stimulation with DMSO (first column-negative control), S1 spike sub-pool (second column) S2 Spike sub-pool (third column), CEFX (fourth column-peptide pool positive control) and SEB (fifth column-superantigen positive control).

**Supplemental Figure 11**

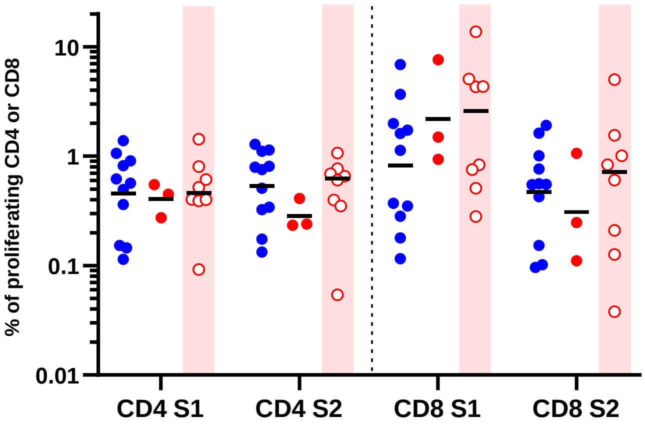

**Supplemental Figure 11. breadth of proliferative response at d180**

Dot plot represent the percentage of proliferating CD4 (left part) and CD8 (right part) T cells stimulated with either S1 spike sub-pool (first and third groups) or S2 Spike sub-pool (second and fourth groups) in subjects representing the RD (blue dots), SD (red dots) and SD+vax (empty red dots and pink shaded area, highlighting that before d180 visit the volunteers have received a COVID-19 approved vaccine) groups.

**Supplemental Figure 12**

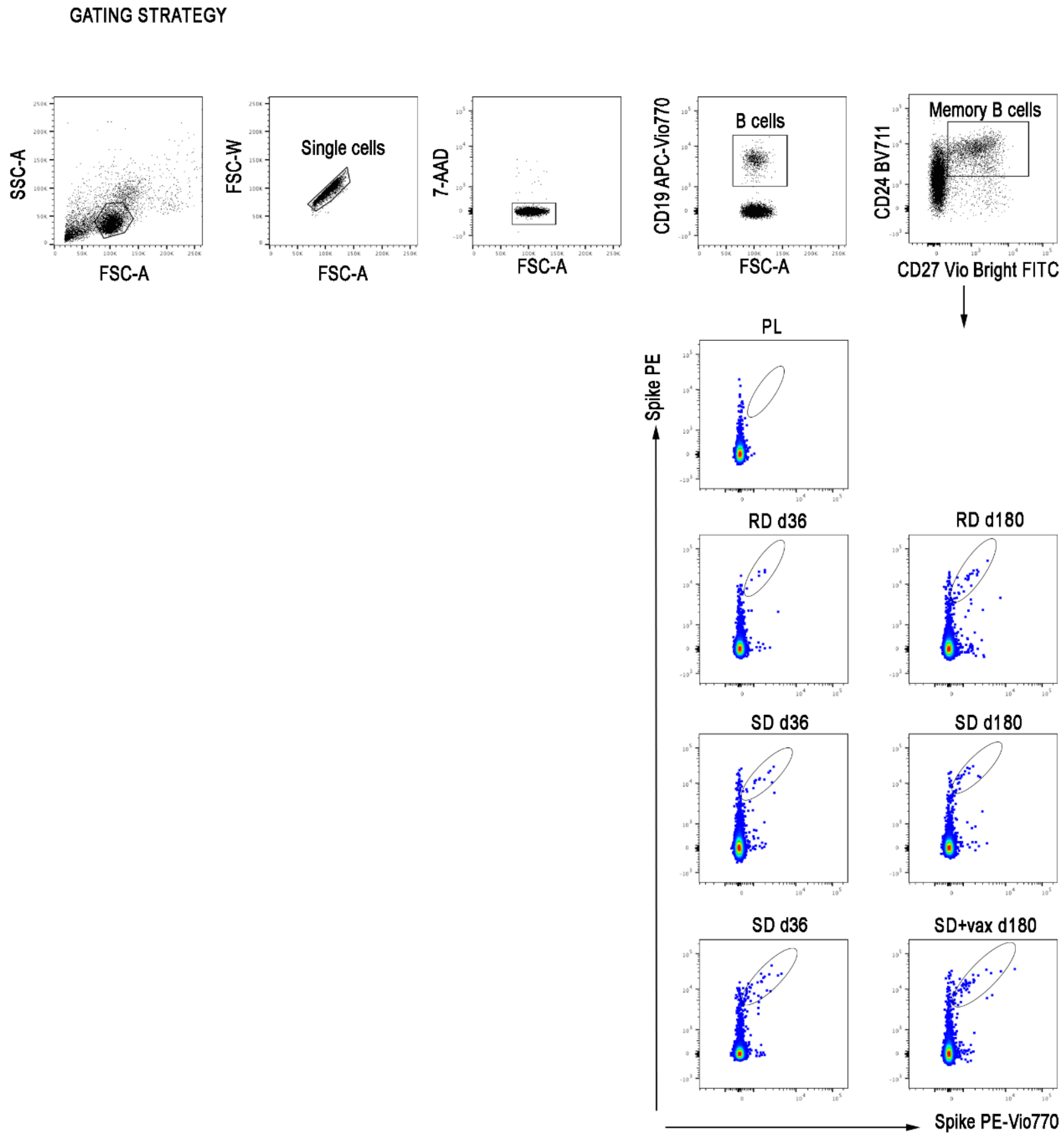

**Supplemental Figure 12. Spike-specific B cell staining gating strategy**
Representative plots for gating strategy (upper lane) of B cell populations and Spike-specific memory B cells in Placebo (upper plot), RD (second lane plots), SD (third lane plots) and SD+vax (fourth lane plots) at day 36 (left column) and day 180 (right column).

**Supplemental Methods**

**Spike, RBD and Nucleocapsid Serology**

Determination of Spike binding antibodies in serum from all participants at all visits was performed using COVID-SeroIndex, Kantaro Quantitative SARS-CoV-2 IgG Antibody Kit (R&D Systems) following manufacturer's instruction. Data are expressed as arbitrary units (AU)/ml; LLOQ is 3,2 AU/mL and ULOQ is 160 AU/mL. Conversion into IgG concentrations expressed as binding antibody international units (BAU)/mL was calculated by multiplying AU/mL values by the conversion factor of 0,0235 and by the final serum dilution (i.e. 200), according to manufacturer's indication. For antibodies to Nucleocapsid and RBD, the semiquantitative SARS-CoV-2 IgG and the SARS-CoV-2 IgG II Quant chemiluminescence microparticle assays (CMIA, Abbott Diagnostics, Chicago, IL, USA) were respectively used, all run on the ARCHITECT platform and according to manufacturer's instructions. For N, data are expressed as index, whereby an index value > 1.4 is considered positive; For RBD assay, data are expressed as arbitrary units (AU)/ml, and the positivity cutoff is 50 AU/ml. For quantitative analysis, the LOD is 6.8 AU/ml, the LOQ is 21 AU/ml, and the analytical Measuring Interval is 21-40.000 (extendable to 80.000). For conversion purpose, the mathematical relationship of the Abbott AU/mL unit to the WHO BAU/mL unit follow the equation: BAU/mL = 0.142 × AU/mL. For plotting and calculations, all values below LOD/LOQ were reported as ½ LOD/LOQ.

**SARS-CoV-2 plaque reduction neutralization test (PRNT)**

The SARS-CoV-2 PRNT50 titers of vaccinated volunteers' serum samples were determined by means of a plaque reduction neutralization test (PRNT) developed and run at Viroclinics Biosciences. Briefly, a standard number of SARS-CoV-2 (Bav/Pat1/2020 strain) infectious units were incubated with eight two-fold serial dilutions of heat inactivated serum, starting from 1:8 and up to 1:1024. After a 1 hour pre-incubation, the virus/serum mixtures were inoculated on VeroE6 cells (ATCC) for 1 hour, than washed, replaced with infection medium and the cells were left overnight. After 16 to 24 hours, the cells were formalin-fixed, permeabilized with ethanol, and incubated with primary anti-SARS-CoV-2 nucleocapsid monoclonal antibody (Sino Biological 40143-MM05 clone #05) followed by a secondary anti-mouse IgG horseradish peroxidase conjugate (Life Technologies A16072) and TrueBlue substrate (KPL 50-78-02), which forms a blue precipitate on virus-positive cells. Images of all wells were acquired by an ImmunoSpot analyzer (Cellular Technology Limited, CTL), equipped with software capable to accurately count the virus positive cells. The 50 and 80% neutralization titers were calculated according to a method described earlier.[1] During assay validation, PRNT80 met the criteria for specificity, accuracy, precision, linearity, dilutional linearity and end of run analysis. The LLOQ, ULOQ and the assay range were set at PRNT80 21-3972. For plotting and calculations, all values below LLOQ were reported as ½ LLOQ.

**Pseudotyped VSV neutralization assay**

A subset of 200 d36 serum samples was tested by a validated pseudotyped virus neutralisation assay (PNA) that assessed particle entry-inhibition, developed and run at Nexelis Laval (Canada). The assay is described in detail in an earlier publication.[2] Briefly, pseudotyped virus particles containing a luciferase reporter were made from a modified vesicular stomatitis virus (VSVDG) backbone expressing the full-length S protein of SARS-CoV-2 from which the last 19 amino acids of the cytoplasmic tail were removed. For the purpose of this study, pseudotyped Spike were from either Wuhan-Hu-1 (D614-validated) or from delta variant (B.1.617.2). Seven two-fold serial dilutions of heat-inactivated serum samples were prepared and incubated at 37 °C with 5% $CO_2$ for 1h with pseudotyped virus at a predefined target working dilution.

Serum-virus complexes were then transferred onto 96 well white flat-bottom plates (Corning), previously seeded overnight with Vero E6 cells and incubated at 37 °C and 5% CO2 for 20 h. Following this incubation, luciferase substrate from ONE GloTM Ex luciferase assay system (Promega) was added to the cells. Plates were then read to quantify relative luminescence units (RLU), inversely proportional to the level of neutralising antibodies present in the serum. The neutralising titre of a serum sample was calculated as the reciprocal serum dilution corresponding to the 50% neutralisation antibody titre (NT50) for that sample; for Wuhan dataset, the NT50 titres were transformed to international units per mL (IU/mL), based on the WHO international standard for anti-SARS-CoV-2 immunoglobulin, using a conversion factor determined during assay validation (1/1872). The assay's cut-off and LLOQ were 5.3 IU/mL (10 as NT50) and 5.9 IU/mL, respectively. For plotting and calculations, all values below LLOQ were reported as ½ LLOQ.

**PhenoSense SARS CoV-2 nAb Assay**

A subset of 20 d36 serum samples were tested for measurement of nAb activity using the PhenoSense SARS CoV-2 nAb Assay (Monogram Biosciences, South San Francisco, CA), based on HIV-1 pseudovirions that express the SARS CoV-2 spike protein. The pseudovirus is prepared by co-transfecting HEK293 producer cells with an HIV-1 genomic vector that contains a firefly luciferase reporter gene together with a SARS CoV-2 spike protein expression vector. For the purpose of this study, the pseudotyped Spike was from SARS-CoV-2 Wuhan strain (D614). Neutralizing antibody activity is measured by assessing the inhibition of luciferase activity in HEK293 target cells co-trasfected to express the ACE2 receptor and TMPRSS2 protease following pre-incubation of the pseudovirions with serial dilutions of the (previously heat-inactivated) serum specimen for one hour at 37 °C. Cell suspension and virus-serum mix are incubated 3 days, then Steady Glo (Promega) is added and RLUs are measured at a luminometer. Neutralizing antibody titers are reported as the reciprocal of the serum dilution of serum samples at which RLUs were reduced by 50% (ID50) compared to pseudovirus control wells. To ensure that the measured nAb activity is SARS CoV-2 nAb specific, each test specimen is also assessed using a non-specific pseudovirus (specificity control) that expresses a nonreactive envelope protein of an unrelated virus (avian influenza virus H10N3). The LLOQ, corresponding to the reciprocal of the serum starting dilution, is set at 40, while the ULOQ is established by the final serum dilution, corresponding to 787320.

**IFNγ ELISpot assay**

The frequency of IFNγ-producing T cells was assessed by enzyme-linked immunosorbent assay (Human IFN-γ ELISpot plus kit; Mabtech) after specific stimulation. PBMCs were thawed quickly in 37°C water bath and collected in thawing medium [CTL Wash supplemented medium in RPMI-1640 (Sigma-Aldrich) and supplementing with L-glutamine (Gibco) and Benzonase (Merck) 50U/ml]. After one wash, cells were suspended into 50 ml polypropylene vented cap tubes with prewarmed medium [RPMI-1640 (Sigma-Aldrich) supplemented with 10% heat-inactivated highly defined FBS (Cytiva HyClone), 2 mmol/L L-glutamine, 10 mmol/L HEPES buffer (N-2-hydroxyethylpiperazine-N-2-ethane sulfonic acid, Sigma-Aldrich), 100 U/ml penicillin, and 100 µg/ml streptomycin (Gibco)], hereafter termed R10. Cells were rested at $2x10^6$ cells/ml density in incubator at 37°C and 5% of CO2 for at least 16 hours before plating.
PBMCs were then counted using Muse instrument (Luminex), plated in ELISpot assays at $2 \times 10^5$ cells/ and stimulated with specific peptides pools. For main IFNγ ELISpot analysis at all three available study visits (d22, d36 and d180), cells were stimulated with 4 peptide pools (S1a, S1b-including RBD domain, S2a and S2b) covering the full-length Spike Wuhan protein. [3] Furthermore, PBMCs at day 22 were tested using commercially available S1 and S2 peptide pools (JPT, Pepmix) of the Delta (Spike B.1.617.2 ) and Omicron (Spike B.1.1.52) variants, in comparison with peptide pools covering Wuhan reference spike from the same commercial source. Plates with cells and stimuli were incubated for 18-20 hours at 37°C and 5% of

CO2. At the end of incubation, the colorimetric assay was developed according to manufacturer's instructions. Single color spots were counted at S6 ImmunoSpot Ultimate UV image analyzer (Cellular Technology Limited, CTL). Spontaneous cytokine production (background) was assessed by incubating PBMC with DMSO, the peptides diluent (Sigma). Results are expressed as spot forming cells (SFC)/$10^6$ PBMCs in stimulating cultures after subtracting spontaneous background, and then summing the counts for the four or two peptide pools to express total Spike response. Data from four volunteers (PL: 1020036 d36; 1020049 d22 and d36; RD: 1020025 d22 and d36; SD: 1020057 d36) were excluded from IFNγ ELISpot main analyses since their spontaneous IFNγ secretion in DMSO wells was above the mean+1standard deviation of the study population (mean=32 SFC, SD=76.37, mean+1SD=109 SFC/million PBMC), leading to unreliable quantitative analysis.

**Human IFNγ-IL5 Fluorospot**

Th1/Th2 response on day 36 frozen PBMCs was assessed using human IFNγ-IL5 Fluorospot (Mabtech). Cells were thawed and plated at 2,5x$10^5$ cells/well and stimulated with 4 peptide pools covering the full-length Spike Wuhan protein (S1a, S1b, S2a and S2b). Precoated plates were activated and blocked with R10 medium, and, after 18-20 hours of cells and stimuli incubation at 37°C with 5% of CO2, the ELISpot assay was developed according to manufacturer's instructions. Spot analysis was performed with an automated C.T.L. ELISpot reader equipped with filters for excitation 480 nm/emission 520 nm (FITC) for IFNγ detection and excitation 570 nm/emission 600 nm (Cy3) for IL5 detection. Results are expressed as spot forming cells (SFC)/$10^6$ PBMCs in stimulating cultures after subtracting spontaneous background.

**Intracellular Cytokines Staining**

PBMC were thawed and treated as described before for ELISpot assays. After ON resting and counting, cells were aliquoted into 5ml polypropylene conical snapcap tubes (Grenier Bio-one, AU), 1x$10^6$ cells/tubes (200µl final volume) and stimulated with either DMSO as negative control, CEFX (20 µg/ml, JPT, GE) or SEB (0,1 mg/ml Sigma-Aldrich, USA, Mo) as positive controls, S1a+b pool or S2a+b pool (0,3 mg/ml) for 5 hours at 37°C 5% $CO_2$. 1µl of GolgiPlug (BD, USA, NJ) was added after 1h of incubation for intracellular cytokine accumulation.

After stimulation, cells were transferred in clean FACS tube, washed with cold PBS to stop the stimulation, and stained with anti-CD107a PE-CF594 clone (H4A3) and FVS780 (both BD) for 20' at RT. Cells were washed with PBS 2% FBS 2mM EDTA, then fixed and permeabilized with BD Fixation/Permeabilization Kit (BD) following manufacturer instructions and stained intracellularly with CD3 BUV395 clone UCHT1, CD4 BUV737 clone SK3, CD8 BV605 clone SK1, CD154 BB700 clone TRAP1, IFNγ APC clone B27, IL-2 PE clone mQ1-17H12, TNFα BV650 clone Mab11, CD69 APC-R700 clone FN50 in the presence of Brilliant Stain Buffer (all from BD) for 30' at 4°C. Cells were finally acquired at Fortessa LSR (BD).

Sample analysis was performed with FlowJo software (version 10.8.1, BD) following gating strategy depicted in Supplementary Figure F. Multi-functional cells were identified by combining each single-function-positive gate with Boolean tools available in FlowJo. For each donor, the percentage of cells responding to Spike peptides-pools were subtracted of their negative control. Polyfunctional analysis of CD4 and CD8 T cells were run using SPICE software (version 6.1, NIH, USA, MD).

**Proliferation Assay**

PBMC were thawed as described before, counted after centrifugation and stained with 1 µl/sample of CellTrace Violet (Thermofisher, USA, MA). After staining, cells were washed extensively, resuspended in R10 medium and aliquoted into a 24-well plate at 2x$10^6$ cells/well (1ml final volume). Cells were then stimulated with either DMSO as negative control, SEB (0,1 mg/ml Sigma-Aldrich, USA, Mo) as positive control, S1a+b pool or S2a+b pool (0,3 mg/ml) or CEFX (1µg/ml) for 5 days at 37°C 5% $CO_2$. After stimulation, cells were harvested and transferred into new FACS tubes to be stained with CD3 BUV395 clone UCHT1, CD4 BUV737 clone SK3, CD8 BV605 clone SK1 (all from BD) and CD19 APC-Vio770 clone LT19 (Miltenyi) in the presence of Brilliant Stain Buffer (BD) for 30' at 4°C. Sample analysis was performed with FlowJo software (version 10.8.1, BD) following gating strategy depicted in Supplementary Figure J. For each donor, the percentage of cells proliferating to Spike peptides-pools were subtracted of their negative control and summed to provide total Spike proliferative response.

**Detection of Spike-specific memory B cells**

Detection of antigen-specific memory B cells was performed with the SARS-CoV-2 Spike B cell analysis kit (Miltenyi Biotec #130-128-022). Briefly, the kit allows the formation of tetramers made of a recombinant SARS-CoV-2 Spike-Protein (expressed in HEK)-Biotin with either Streptavidin-PE or Streptavidin-PE-Vio770. ~$5x10^6$ previously frozen PBMC samples were prepared and stained with antibody staining mix containing the Spike tetramers, the 7-AAD and the fluorochrome-conjugated antibodies (CD19 APC-Vio770 clone LT19, CD27 Vio Bright FITC clone M-T271, CD24 BV711 clone ML5, IgA VioGreen clone IS11-8E10, IgM APC clone Pj2-22H3, IgG VioBlue clone IS11-3B2.2.3-all from Miltenyi; CD24 BV711 clone ML5 from BD). Memory B cells were identified as CD19+CD24+CD27+ and Spike-specific memory B cells were double positive for PE and PE-Vio770. Samples were acquired on Fortessa LSR (BD) (BD) and analysed using FlowJo (version 10.7.1, BD). Limit of detection (LOD) and limit of quantification (LOQ) were calculated as previously reported. [4-7] Briefly, LOD was calculated as 20x100/total no. of events and LOQ was computed as 30x100/total no. of events. Gating strategy is illustrated in supplementary figure L.

**Supplemental authors list (COVITAR CLINICAL TRIAL GROUP)**

| INSTITUTION/AFFILIATION | COVITAR GROUP |
|---|---|
| Centro Ricerche Cliniche di Verona- CRC, Verona, Italy | Luigi Ziviani<br>Feliciana Malescio<br>Irene Turrini |
| ARC-Net Research Center, University of Verona, Verona, Italy | Rita Lawlor |
| San Giuseppe Moscati Hospital, Avellino, Italy | Annamaria Romano<br>Mariagrazia Nunziata<br>Salvatore Armato<br>Nicole Mazzeo |
| "D. Cotugno" Hospital, Azienda Specialistica dei Colli, Naples, Italy | Maria Aurora Carleo<br>Chiara Dell'Isola<br>Raffaella Pisapia<br>Agostina Pontarelli |
| Unit of Infectious Diseases and Hepatology, Azienda Ospedaliero–Universitaria of Parma, Parma, Italy | Andrea Olivani |
| Department of Medicine and Surgery, University of Parma, Parma, Italy | Sara Grasselli<br>Diletta Laccabue |
| UOC di Malattie Infettive<br>Ospedale Guglielmo da Saliceto<br>AUSL Piacenza, Italy | Maria Cristina Leoni<br>Franco Paolillo<br>Annalisa Mancini |
| Struttura Complessa Pneumologia - Azienda Sanitaria Universitaria Giuliano-Isontina – Trieste, Italy | Barbara Ruaro<br>Marco Confalonieri<br>Francesco Salton |
| Dept Public Health and Infectious Disease Sapienza University of Rome, Infectious Disease Unit, SM Goretti Hospital Latina, Italy | Giulia Mancarella |
| Infectious Disease Unit, SM Goretti Hospital Latina, Italy | Raffaella Marocco<br>Margherita De Masi<br>Valeria Belvisi |
| Fondazione Policlinico Universitario A. Gemelli IRCCS – Roma, Italy | Silvia Lamonica<br>Antonella Cingolani<br>Cristina Seguiti |
| UO Malattie Infettive, ASST Cremona, Italy | Paola Brambilla<br>Alice Ferraresi<br>Matteo Lupi |
| Infectious Diseases Unit, Foundation IRCCS Ca' Granda Ospedale Maggiore Policlinico, Milan, Italy | Serena Ludovisi<br>Giulia Renisi<br>Roberta Massafra |
| I divisione Malattie Infettive - ASST FBF SACCO, Milan, Italy | Martina Pellicciotta<br>Luciana Armiento |
| UOC Farmacia - ASST FBF SACCO, Milan, Italy | Stefania Vimercati<br>Mariagrazia Piacenza |

| | |
|---|---|
| Infectious diseases Unit, ASST Monza and University of Milano-Bicocca, Italy | Paolo Bonfanti |
| Infectious diseases Unit, ASST Monza, Italy | Paola Columpsi |
| Phase 1 Research Unit, ASST Monza and University of Milano-Bicocca, Italy | Marina Elena Cazzaniga |
| Clinica di Malattie Infettive e Tropicali ASST-Sette Laghi, Varese, Italy | Cristina Rovelli |
| Malattie Infettive e Tropicali presso l'Università di Pavia, Italy | Mariaelena Ceresini<br>Letizia Previtali |
| Unit of Infectious Diseases, Department of Medical Sciences, University of Torino, Italy | Laura Trentini<br>Chiara Alcantarini<br>Walter Rugge<br>Stefano Biffi |
| ASL Vercelli Malattie Infettive, Vercelli, Italy | Federica Poletti<br>Roberto Rostagno<br>Roberta Moglia |
| Centro di Farmacologia Clinica per la Sperimentazione dei Farmaci - Azienda Ospedaliero-Universitaria Pisana - via Roma, 67 - 56126 Pisa, Italy | Ferdinando De Negri<br>Elisabetta Fini<br>Alice Cangialosi |
| Malattie infettive, Dipartimento di Scienze Mediche e Chirurgiche - AOU Policlinico Foggia, Italy | Serena Rita Bruno<br>Marianna Rizzo<br>Mariangela Niglio |
| UOC Farmacia - AORN S.Anna e S. Sebastiano, Caserta, Italy | Anna Dello Stritto |
| UOC Organizzazione e Programmazione Servizi Ospedalieri e Sanitari - AORN S.Anna e S. Sebastiano, Caserta, Italy | Alfredo Matano |
| UOC Patologia Clinica - AORN S.Anna e S. Sebastiano, Caserta, Italy | Arnolfo Petruzziello |
| UOC Malattie Infettive - Fondazione IRCCS Policlinico San Matteo di Pavia, Italy | Pietro Valsecchi<br>Teresa Pieri |
| UOC Malattie Infettive - PO V. Emanuele II, Bisceglie (BT), Italy | Mauro Altamura<br>Angela Calamo |
| UO Farmacia Ospedale Mons. Dimiccoli, Barletta, Italy | Anna Giannelli |
| Dipartimento di Prevenzione ASL, BT, Andria, Italy | Stefania Menolascina |
| UOC Malattie Infettive, AOU Policlinico Umberto 1; Sapienza Università di Roma, Italy | Silvia Di Bari<br>Vera Mauro<br>Raissa Aronica |
| Department Infectious Diseases St Anna Hospital and University Ferrara, Italy | Daniela Segala<br>Rosario Cultrera<br>Laura Sighinolfi |

| | |
|---|---|
| Department of Health Promotion Sciences, Maternal and Infant Care, Internal Medicine and Medical Specialties, University of Palermo, 90127 Palermo, Italy | Michelle Abbott<br>Andrea Gizzi<br>Federica Guida Marascia<br>Giacomo Valenti |
| Department of Infectious Diseases - EO Ospedali Galliera – Genova, Italy | Marcello Feasi<br>Nicoletta Bobbio<br>Filippo Del Puente |
| Exxom group, Milan, Italy | Martina Frascà<br>Miriam Mazzoleni<br>Nadia Garofalo |
| ReiThera Srl, Rome, Italy | Virginia Ammendola<br>Fabiana Grazioli<br>Federico Napolitano |
| B Cell Unit, Immunology Research Area, Bambino Gesù Children’s Hospital, IRCCS, Rome, Italy | Valentina Marcellini |

**Study protocol**

### 1 CLINICAL STUDY PROTOCOL

Protocol Title: A Phase II/III, Randomized, Stratified, Observer-Blind, Placebo-Controlled Study to Evaluate the Efficacy, Safety, and Immunogenicity of GRAd-COV2 Vaccine in Adults Aged 18 Years and Older

Protocol Number: RT-CoV-2_01

Study Acronym: COVITAR (GRAd-**COV**2 **ITA**lian Vaccine **R**eithera Study)

Short Title: Phase II/III Observer-blinded, Placebo-controlled Study of GRAd-COV2 for the Prevention of COVID-19 in Adults

| | |
|---|---|
| **EudraCT Number:** | 2020-005915-39 |
| **Phase of Development:** | II/III |
| **Sponsor:** | ReiThera SrL<br>Via di Castel Romano 100, 00128 Castel Romano, RM (Italy)<br>Tel: +39 06 99775399 |
| **Medical Monitor:** | Roberto Camerini, MD, Medical Director<br>ReiThera Srl,<br> |
| **Protocol Version:** | 2.0 |
| **Protocol Date:** | Final Version: 11 June 2021 |

-CONFIDENTIAL-

All financial and nonfinancial support for this study will be provided by ReiThera Srl. The study will be conducted according to the International Council for Harmonisation (ICH) Technical Requirements for Registration of Pharmaceuticals for Human Use, E6(R2) Good Clinical Practice (GCP) Guidance.

#### PROTOCOL APPROVAL SIGNATURES

| | |
|---|---|
| **Protocol Title:** | A Phase II/III, Randomized, Stratified, Observer-Blind, Placebo-Controlled Study to Evaluate the Efficacy, Safety, and Immunogenicity of GRAd-COV2 Vaccine in Adults Aged 18 Years and Older |
| **Protocol Number:** | RT-CoV-2_01 |

The signature below provides the necessary assurance that this trial will be conducted according to all stipulations of this protocol, including all statements regarding confidentiality, and according to local legal and regulatory requirements and applicable ICH E6 GCP guidelines.

I agree to conduct this trial in compliance with the International Conference on Harmonization Tripartite Guideline on Good Clinical Practice (E6R2) and applicable regulatory requirements.

I agree to conduct this trial in accordance with the current protocol and will not make changes to this protocol without obtaining the sponsor's approval and National Ethical committee for COVID approval, except when necessary to protect the safety, rights, or welfare of subjects.

**Sponsor Signatory**

| | |
|---|---|
| Antonella Folgori<br>Chief Executive Officer<br>ReiThera Srl<br>Via di Castel Romano 100<br>00128 Rome, Italy | ____________________<br>Signature<br><br>____________________<br>Date (DD-Mmm-YYYY) |

This document is confidential.

**Coordinating Investigator**

Simone Lanini
Dirigente Medico INMI Lazzaro
Spallanzani IRCCS
Via Portuense 292, Roma – Italy


______________________________
Signature

______________________________
Date (DD-Mmm-YYYY)

**Medical Monitor**

Roberto Camerini
Medical Director
ReiThera Srl
Via di Castel Romano 100
00128 Rome, Italy


______________________________
Signature

______________________________
Date (DD-Mmm-YYYY)

#### INVESTIGATOR SIGNATURE PAGE

**Protocol Title:** A Phase II/III, Randomized, Stratified, Observer-Blind, Placebo-Controlled Study to Evaluate the Efficacy, Safety, and Immunogenicity of GRAd-COV2 Vaccine in Adults Aged 18 Years and Older

**Protocol Number:** RT-CoV-2_01

I have read and understood all sections of the protocol entitled “A Phase II/III, Randomized, Stratified, Observer-Blind, Placebo-Controlled Study to Evaluate the Efficacy, Safety, and Immunogenicity of GRAd-COV2 Vaccine in Adults Aged 18 Years and Older” and the most recent version of the Investigator’s Brochure.

I agree to supervise all aspects of the protocol and to conduct the clinical investigation in accordance with the current Protocol, the International Council for Harmonisation (ICH) Technical Requirements for Registration of Pharmaceuticals for Human Use, E6(R2) Good Clinical Practice (GCP) Guidance, and all applicable government regulations. I will not make changes to the protocol before consulting with ReiThera Srl. or implement protocol changes without IRB/IEC approval except to eliminate an immediate risk to participants.

I agree to administer study treatment only to participants under my personal supervision or the supervision of a sub-investigator. I will not supply study treatment to any person not authorized to receive it. I also agree that persons debarred from conducting or working on clinical studies by any court or regulatory agency will not be allowed to conduct or work on studies for the Sponsor or a partnership in which the Sponsor is involved. I will immediately disclose it in writing to the Sponsor if any person who is involved in the study is debarred, or if any proceeding for debarment is pending, or, to the best of my knowledge, threatened.

I will not disclose confidential information contained in this document including participant information, to anyone other than the recipient study staffs and members of the IRB/IEC. I agree to ensure that this information will not be used for any purpose other than the evaluation or conduct of the clinical investigation without the prior written consent from ReiThera Srl. I will not disclose information regarding this clinical investigation or publish results of the investigation without authorization from ReiThera Srl.

The signature below provides the necessary assurance that this study will be conducted according to all stipulations of the protocol, including statements regarding confidentiality, and according to local legal and regulatory requirements, and ICH E6(R2) GCP guidelines.

Title, Name of the Investigator: ____________________________________________

City, Institution: _________________________________________________________

Signature ______________________________________________ Date _____________

This document is confidential.

### 2 SYNOPSIS

| | |
|---|---|
| **Title of Study:** | A Phase II/III, Randomized, Stratified, Observer-Blind, Placebo-Controlled Study to Evaluate the Efficacy, Safety, and Immunogenicity of GRAd-COV2 Vaccine in Adults Aged 18 Years and Older |
| **Protocol Number:** | RT-CoV-2_01 |
| **Investigators/Study Sites:** | Multicenter International (approx. 24 sites for phase II and 80 sites for phase III) |
| **Phase of Development:** | Phase II/III |

**Objectives and Endpoints**

| **Objective** [a] | **Estimand** [b] **Description/Endpoint** |
|---|---|
| **Primary for Phase III part of the Study** | |
| 1) To estimate the efficacy of a single or repeated (21 days apart) IM dose of GRAd-COV2 compared to placebo for the prevention of COVID-19 in adults<br><br>≥ 18 years of age | **Population:** Full analysis set, excluding participants who are seropositive at baseline. |
| | **Endpoint:** A binary response, whereby a participant is defined as a COVID-19 case if their first case of SARS-CoV-2 RT-PCR-positive symptomatic illness occurs ≥ 28 days post first dose or ≥ 7 days after the second dose of study intervention (depending on the selected regimen). Otherwise, a participant is not defined as a COVID-19 case. The endpoint will be treated as time to event |
| | **Intercurrent events**: The intercurrent events (ie, withdrawals before having met the primary efficacy endpoint, deaths for reasons that are unrelated to COVID-19 and COVID-19 infections that occur before 28 days post single dose or before 7 days post second dose) will be handled using a "while on treatment" strategy, where the time to COVID-19 is censored at the date of withdrawal or death or early COVID-19 infection. In case the double-dose schedule is selected for phase III, subjects missing the second dose will be excluded from the primary analysis. |
| | **Summary measure:** VE, calculated as 1-HR, where HR (=Hazard Ratio) between GRAd-COV2 and placebo is estimated with a stratified Cox proportional hazard model. |
| 2) To assess the safety and tolerability of a single or repeated (21 days apart) IM dose of GRAd-COV2 compared to placebo in adults ≥ 18 years of age | a) Incidence of AEs for 28 days post each dose of study intervention. |
| | b) Incidence of SAEs, MAAEs, and AESIs from Day 1 post treatment through Day 360. |

This document is confidential.

| **Objective [a]** | **Estimand [b] Description/Endpoint** |
|---|---|
| **Primary for phase II part of the Study** | |
| 1) To assess the reactogenicity of a single or repeated (21 day apart) IM dose of GRAd-COV2 compared to placebo in adults ≥ 18 years of age | Incidence of local and systemic solicited AEs for 7 days post each dose of study intervention |
| 2) To identify the schedule to be further studied in phase III by assessing antibody responses to S antigen following a single or repeated (21 days apart) IM dose of GRAd-COV2 or placebo (first 450 participants) | a) Post-treatment GMTs and GMFRs from day of dosing baseline value to 35 days post first dose in SARS-CoV-2 S and/or RBD antibodies. |
| | b) Proportion of participants who have a post-treatment seroresponse (≥ 4-fold rise in titers from day of dosing baseline value to 35 days post first dose) to the S and/or RBD antigens of GRAd-COV2. |
| **Key Secondary (study phase indicated in parenthesis)** | |
| To estimate the efficacy of 1 single or repeated (21 days apart) IM dose of GRAd-COV2 compared to placebo for the prevention of severe or critical symptomatic COVID-19 (Phase III) | Time to first SARS-CoV-2 RT-PCR-positive severe or critical symptomatic illness occurring ≥ 28 days post first dose or ≥ 7 days after second dose of study intervention. |
| **Other Secondary (study phase indicated in parenthesis)** | |
| 1) To estimate the efficacy of a single or repeated (21 days apart) IM dose of GRAd-COV2 compared to placebo for the prevention of SARS-CoV-2 infection - Asymptomatic cases (Phase III) | Proportion of participants who have a post-treatment response (negative at baseline to positive post treatment with study intervention) for SARS-CoV-2 Nucleocapsid antibodies over time. |
| 2) To estimate the efficacy of a single or repeated (21 days apart) IM dose of GRAd-COV2 compared to placebo for the prevention of symptomatic COVID-19 using CDC criteria (Phase III) | Time to first case of SARS-CoV-2 RT-PCR- positive symptomatic illness occurring ≥ 28 days post first dose or ≥ 7 days after second dose of study intervention using CDC criteria. |
| 3) To estimate the efficacy of 1 single or repeated (21 days apart) IM dose of GRAd-COV2 compared to placebo for the prevention of COVID-19-related Emergency Department visits (Phase III) | Time to first COVID-19-related Emergency Department admission occurring ≥ 28 days post single dose or ≥ 7 days post second dose of study intervention. |
| 4) To evaluate VE to prevent death caused by COVID-19 (Phase III). | Time to COVID-19 related death occurring ≥ 28 days post single dose or ≥ 7 days post second dose of study intervention. |

This document is confidential.

| **Other Secondary (study phase indicated in parenthesis)** | |
|---|---|
| 5) To assess antibody responses to S antigen following a single or repeated (21 days apart) IM dose of GRAd-COV2 or placebo (Phase II and Illness Visits only) | a) Post-treatment GMTs and GMFRs from day of dosing baseline value to 35 days post first dose (14 days post second dose) in SARS-CoV-2 S and/or RBD antibodies |
| | b) The proportion of participants who have a post-treatment seroresponse (≥ 4-fold rise in titers from day of dosing baseline value to 35 days post first dose) to the S and/or RBD antigens of GRAd-COV2. |
| 6) To determine anti-SARS-CoV-2 neutralizing antibody levels in serum following 1 single or repeated (21 days apart) IM dose of GRAd-COV2 or placebo (Phase II and Illness Visits only) | a) Post-treatment GMTs and GMFRs from day of dosing baseline value to 35 days post first dose (14 days post second dose) in SARS-CoV-2 neutralizing antibodies. |
| | b) Proportion of participants who have a post-treatment seroresponse (≥ 4-fold rise in titers from day of dosing baseline value to 35 days post first dose) to GRAd-COV2 as measured by SARS-CoV-2 neutralizing antibodies. |

| | |
|---|---|
| **Study Design:** | Multicenter study assessing the safety, efficacy, and immunogenicity of GRAd-COV2 compared to placebo for the prevention of COVID-19. Up to approximately 80 sites in 12 countries will participate in this study. Study sites selection will be considered based on predicted transmission rates of SARS-CoV-2 in those locations. Participants will be adults ≥ 18 years of age who are healthy or have medically stable chronic diseases and are at increased risk for SARS-CoV-2 acquisition and COVID-19.<br><br>In the phase II part approximately 900 participants will be randomized in a 1:1:1 ratio to receive 1 single IM dose of GRAd-COV2 2 x $10^{11}$ vp plus 1 dose of saline placebo after 21 days (n= approximately 300 subject) or 2 repeated (21 days apart) IM dose of GRAd-COV2 1 x $10^{11}$ (n = approximately 300 subjects) or two doses of saline placebo (n = approximately 300 subjects) on day 1 and day 22. Randomization will be stratified based on age (below / equal or above 65 years) and, if participants are < 65 years of age, based on the presence or absence of risk factors for severe illness from COVID-19 based on CDC recommendation as of May 2020. Therefore, there will be 3 strata for randomization: ≥ 65 years, < 65 years and categorized to be at increased risk (“at risk”) for the complications of COVID-19, and < 65 years “not at risk”. Risk will be defined based on the study participants’ relevant past and current medical history. At least 25% of enrolled participants, will be either ≥ 65 years of age or < 65 years of age and “at risk” at Screening.<br><br>These 900 participants randomized in each age group, including at least 225 participants ≥ 65 years of age or < 65 of age “at risk”, will be assessed for safety, reactogenicity and immunogenicity of GRAd-COV2. An independent DSMB will provide oversight, to ensure safe and ethical conduct of the study and, together with a Steering Committee, based on safety (900 participants 1 week after dosing) and immunogenicity (450 participants at week 5 after the first dosing) data generated in phase II part, will recommend the expansion to phase III and the best regimen to be used. |

| | |
|---|---|
| | Once the phase III expansion is granted, according to the epidemic evolution and the availability on the market of alternative vaccine(s) and the characteristics of vaccination campaign, the phase III study design will be adapted following these 3 potential scenarios: 1) a superiority trial vs placebo on overall population; 2 ) a superiority trial vs placebo on an subset of population (low risk subjects for infection outcome) or 3) a non-inferiority trial vs the available alternative vaccine on a surrogate endpoint (correlates of protection), if available.<br><br>Same assumptions can be made for sample size computation in the first two scenarios, being the evaluation of risk referred to the infection outcome and not the likelihood of getting an infection. In case of scenarios 1 and 2, additional 9400 participants will enter the study for the evaluation of safety, efficacy and asymptomatic infection rate (SARS-CoV-2 Nucleocapsid antibodies seroconversion). At present, scenario 3 is not evaluated in terms of sample size, because the assumptions for these computations are not known. If scenario 3 is to be followed due to ethic issues about placebo use, a protocol amendment will be submitted to define the sample size and the other protocol details.<br><br>Participants will remain on study for 1 year following administration of the dose of study intervention (Day 360). If GRAd-COV2 is proven to be safe and efficacious based on the primary endpoint analysis, following discussion at that time with Regulators if appropriate, and the study DSMB recommendations, participants allocated to the placebo group may be offered GRAd-COV2, if doses are available. Participants in the placebo group of clinical trial who will be eligible for available vaccination outside the trial will discuss this opportunity with Investigator based on immunogenicity data collected at day 57 Placebo participants treated with GRAd-COV2 or alternative vaccines will continue to be followed in the study.<br><br>**Blinding:**<br><br>The phase II part of the study is a parallel-group preventive study with 3 arms that is participant and investigator blinded (observer blinded). The blinding of the phase III will depend on the scenario that will be implemented.<br><br>Since the COVID-19 vaccination campaign is rapidly progressing and around 50 % of the target population has received at least one dose of the available authorized vaccines, it is considered no longer feasible and even unethical to keep subjects participating in ongoing blinded clinical trials in the placebo arm. Therefore, considering that all randomized subjects have already reached day 57 visit, which is one of the identified key study time point for immune response assessment, upon approval by both the Competent Authority (CA) and Ethics Committee, the randomization code will be automatically broken for all subjects. The participants assigned to placebo arm will be discontinued from the study as soon as they receive alternative available vaccines in the frame of national vaccination campaign. On the contrary participants belonging the two study arms who received GRAd-COV2 vaccine will continue to be monitored for safety and efficacy according to the protocol even if they will decide to access to the vaccination campaign. |
| **Selection of Patients:** | Main Inclusion Criteria:<br><br>1. Adult female and male, ≥ 18 years of age at the time of consent<br><br>2. Medically stable such that, according to the judgment of the investigator, hospitalization within the study period is not anticipated and the participant |

This document is confidential.

| | |
|---|---|
| | appears likely to be able to remain on study through the end of protocol-specified follow-up. A stable medical condition is defined as disease not requiring significant change in therapy or hospitalization for worsening disease during the 3 months prior to enrollment<br>3. Able to understand and comply with study requirements/procedures based on the assessment of the investigator<br>4. Contraceptive use by women should be consistent with local regulations regarding the methods of contraception for those participating in clinical studies<br>5. Female participants, (a) Women of childbearing potential must: Have a negative pregnancy test on the day of screening and on Day 1; use one highly effective form of birth control for at least 28 days prior to Day 1 and agree to continue using one highly effective form of birth control through 60 days following administration of study intervention.<br>6. Capable of giving signed informed consent.<br>Main Exclusion Criteria:<br>1. History of allergy to any component of the vaccine<br>2. History of Guillain-Barré syndrome or any other demyelinating condition<br>3. Significant infection or other acute illness, including fever > 37.3 °C on the day prior to or day of randomization<br>4. History of laboratory-confirmed SARS-CoV-2 infection<br>5. Any confirmed or suspected immunosuppressive or immunodeficient state, including asplenia (only for phase II)<br>6. Recurrent severe infections and use of immunosuppressant medication within the past 6 months<br>7. History of primary malignancy except for: (a) Malignancy with low potential risk for recurrence after curative treatment (for example, history of childhood leukaemia) or metastasis (for example, indolent prostate cancer) in the opinion of the site investigator. (b) Adequately treated non-melanoma skin cancer or lentigo maligna without evidence of disease (c) Adequately treated uterine cervical carcinoma in situ without evidence of disease (d) Localized prostate cancer (only for phase II)<br>8. Clinically significant bleeding disorder (eg, factor deficiency, coagulopathy, or platelet disorder), or prior history of significant bleeding or bruising following IM injections or vene puncture<br>9. Severe and/or uncontrolled cardiovascular disease, respiratory disease, gastrointestinal disease, liver disease, renal disease, endocrine disorder, and neurological illness, as judged by the Investigator (mild/moderate well-controlled comorbidities are allowed) (only for phase II)<br>10. Any other significant disease, disorder, or finding that may significantly increase the risk to the participant because of participation in the study, affect the ability of the participant to participate in the study, or impair interpretation of the study data |

| | |
|---|---|
| | 11. Receipt of, or planned receipt of investigational or licensed products indicated for the treatment or prevention of SARS-CoV-2 or COVID-19<br><br>12. Receipt of any vaccine (licensed or investigational) other than licensed influenza vaccines within 30 days prior to and after administration of study intervention<br><br>13. Receipt of immunoglobulins and/or any blood products within 3 months prior to administration of study intervention or expected receipt during the period of study follow-up<br><br>14. Involvement in the planning and/or conduct of this study (applies to both Sponsor staff and/or staff at the study site)<br><br>15. For women only - currently pregnant (confirmed with positive pregnancy test) or breast-feeding<br><br>16. Has donated ≥ 450 mL of blood products within 30 days prior to randomization or expects to donate blood within 90 days of administration of study intervention. |
| **Planned Sample Size:** | The sample size calculations are based on the primary efficacy endpoint evaluated in the phase III part of the study. For the scenarios 1 and 2 (see above), approximately 10000 participants will be randomized in a 1:1 ratio to receive 1 single or repeated (21 days apart) IM dose of either GRAd-COV2 (the active group, n = approximately 5000) or saline placebo (the control group, n = approximately 5000) on Day 1 or Days 1 and 22. Since efficacy analysis is conducted on the phase II and phase III subjects combined, approximately 900 subjects will be enrolled in the phase II part and 9400 subjects in the phase III part of the study (considering that only one of the two vaccine arms evaluated in phase II will enter phase III).<br><br>With the assumptions of a true VE of 70%, a total of approximately 74 first confirmed COVID-19 cases will provide 90% power to conclude that true VE>30%. This would be achieved with approximately 10000 participants in a 1:1 allocation ratio. The calculations assume that the illness rate in the placebo group is 1.25% in six months, the 74 end-point cases required for the primary analysis can be obtained in six months and 10% of participants are non-evaluable or have serological evidence of prior infection with SARS-CoV-2. They also account for conduction of an interim and a primary analysis. The timing of these analyses will be driven by the number of events observed in the study. The interim analysis will be carried out when approximately 50% of the total amount of statistical information is available (ie, 37 cases have been collected). The O'Brien Fleming alpha-spending function has been used to control the overall study-wise type I error at 5% two-sided with 0.3% nominal alpha at the interim analysis and 4.9% at the primary analysis. Since this is an event driven study, the Sponsor may adjust the size of the study or the duration of the follow-up based on the blinded review of the total number of cases of COVID-19 accrued during the study, in addition to the estimated percentages of study participants with serologic evidence of SARS-CoV-2 infection at baseline. |
| **Investigational Therapy:** | GRAd-COV2, $2x10^{11}$ vp as single intramuscular injection or $1x10^{11}$ vp as a repeated intramuscular administration 21 days apart |
| **Reference Therapy** | Placebo: 0.9% (w/v) saline via intramuscular administration as a single intramuscular injection or as a repeated intramuscular administration 21 days apart |

| | |
|---|---|
| **Treatment Duration** | Study intervention will be administered on Day 1or on day 1 and 22. |
| **Statistical Methods and Planned Analyses:** | **Primary Efficacy Endpoint:**<br><br>The primary efficacy endpoint for phase III is a binary response, whereby a participant is defined as a COVID-19 case if their first case of SARS-CoV-2 RT-PCR-positive symptomatic illness occurs ≥ 28 days post single dose or ≥ 7 days post second dose of study intervention. Otherwise, a participant is not defined as a COVID-19 case. For the phase II participants, since all of them receive two doses, cases will be considered valid if they occur ≥ 7 days post second dose. For the phase III participants, the two described conditions will apply depending on the schedule selected in phase II, ie, if the single dose is selected, cases will be considered valid if they occur ≥28 days post dose; if the double dose is selected, cases will be considered valid if they occur ≥ 7 days post second dose.<br><br>**Primary Efficacy Analysis:**<br><br>A Cox proportional hazard model will be used to assess the magnitude of the treatment effect, ie, the hazard ratio (HR) between GRAd-COV2 and placebo will be determined and VE will be estimated by 1– HR. The model will include the treatment and the protocol defined stratification factor as the independent fixed effects. Model assumptions will be checked, and the robustness of the primary analysis will be assessed through various sensitivity analyses. At the interim analysis, VE will be presented with a 2-sided 99.7% CI, and statistical significance will be achieved if the lower limit of this CI is > 30%. At the primary analysis, VE will be presented with a 2-sided 95.10% CI, and statistical significance will be achieved if the lower limit of this CI is > 30%**.**<br><br>**Safety Analysis:**<br><br>**T**he primary safety endpoints will be as follows:<br><br>1. Incidence of AEs in the 28 days post first or second dose of study intervention.<br>2. Incidence of SAEs, MAAEs and AESIs from Day 1 post dose through Day 360.<br>3. Incidence of local and systemic solicited AEs for 7 days post first and second dose of study intervention.<br><br>The last endpoint will be assessed only in the phase II part of the study.<br><br>**Data Safety Monitoring Board:**<br><br>An independent DSMB will review the interim data to safeguard the interests of clinical study participants and to help ensure the integrity of the study. The DSMB and Steering Committee will jointly review unblinded statistical outputs and interim analysis results, provided by the independent unblinded statistician, and make recommendations to the Sponsor. |

[a].Illness Visits: Participants who present with qualifying symptoms will be virologically tested for SARS-CoV-2 and if positive, will complete illness visits.

[b].Estimand is the target of estimation to address the scientific question of interest posed by the primary objective. Attributes of an estimand include the population of interest, the variable (or endpoint) of interest, the specification of how intercurrent events are reflected in the scientific question of interest, and the population-level summary for the variable.

This document is confidential.

AE = adverse event; AESI = adverse event of special interest; CDC = Centers for Disease Control; COVID-19 = coronavirus disease 2019; GMFR = geometric mean fold rise; GMT = geometric mean titer; IM = intramuscular; MAAE = medically attended adverse event; RT-PCR = reverse transcriptase polymerase chain reaction; RBD = receptor binding domain; S = Spike; SAE = serious adverse event; SARS-CoV-2 = severe acute respiratory syndrome- coronavirus-2; VE = vaccine efficacy.

**NOTE: To be considered as a case of COVID-19 for the evaluation of the Primary Efficacy Endpoint, the following criteria must be met:**

- The participant must have experienced at least TWO of the following systemic symptoms: Fever (> 37.8 °C), new or worsening cough, myalgia/muscle pain, fatigue that interferes with activities of daily living, vomiting and/or diarrhea (only one finding to be counted toward endpoint definition), anosmia and / or ageusia (only one finding to be counted toward endpoint definition),

**OR**

- The participant must have experienced at least ONE of the following respiratory signs/symptoms: Pneumonia diagnosed by chest x-ray, or computed tomography scan; oxygen saturation of ≤ 94% on room air or requiring either new initiation or escalation in supplemental O2; new or worsening dyspnea/shortness of breath,

**AND**

- The participant must have at least one NP swab, nasal swab, or saliva sample (or respiratory sample, if hospitalized) positive for SARS-CoV-2 by RT-PCR.

This document is confidential.

### 3 SCHEDULE OF ACTIVITIES

The SoA tables include:

Table 1: Screening Period

Table 2: Treatment and Follow-Up Period - Phase II

Table 3: Treatment and Follow-Up Period – Main Study (Excluding Phase II Participants)

Table 4: Illness Visits (Participants with Qualifying Clinical Symptoms)

*Table 1 Schedule of Activities: Screening Period (All Subjects in Phase II/III)*

| **Procedure / Study Day** | **Day -14 to Day 1 [a]** |
|---|---|
| Informed consent | X |
| Assignment SID number | X |
| Medical history | X |
| Complete physical examination, including height and weight | X |
| Vital signs (including pulse oximetry) | X |
| Pregnancy test – urine or serum [b] | X |
| Assessment of AEs and SAEs | X |
| Concomitant medications | X |
| Verify eligibility criteria | X |

[a] If screening and dosing occur at the same time visit, only one evaluation is required.

[b] If urine tests positive or indeterminate, a quantitative serum β-hCG will be performed for confirmation.

β-hCG = beta-human chorionic gonadotropin; SAE = serious adverse event; SID = subject identification.

*Table 2 Treatment and Follow-up Period – Phase II*

This document is confidential.

| **Procedure** | **Treatment and Follow-up Period** | | | | | | | |
|---|---|---|---|---|---|---|---|---|
| **Day** | **1** | **8 [a]** | **22** | **29 [a]** | **36** | **57** | **180** | **360** |
| **Window (days)** | **NA** | **± 1** | **± 2** | **± 2** | **± 2** | **± 10** | **± 15** | **± 30** |
| Medical history | x | | | | | | | |
| Targeted physical examination | x | | | | | | | |
| Vital signs (including pulse oximetry) | x | | | | | | | |
| Pregnancy test – urine or serum[b] | X (predose) | | X (predose) | | | | | |
| Concomitant medications | x | x | x | x | X | As applicable, for treatment of SAEs, MAAEs, or AESIs | | |
| Verify eligibility criteria | x | | | | | | | |
| **Study intervention administration** | x | | x [d] | | | | | |
| **Efficacy Assessment** | | | | | | | | |
| Weekly telephone/email/text contacts – monitoring for COVID-19 qualifying symptoms [c] | Weekly throughout this period | | | | | | | |
| Nasal swab for SARS-CoV-2 RT-PCR (local laboratory) | x (predose) | | x (predose) | | | | | |
| Serum sample for SARS-CoV-2 serology testing for abs anti N antigen | x (predose) | | x (predose) | | X | x | x | x |
| **Immunogenicity Assessment** | | | | | | | | |
| Serum sample for SARS-CoV-2 nAbs assessment | x (predose) | | x (predose) | | X | x | x | x |
| PBMCs for assessment of T-cell responses (subset only in one site) | | | x (predose) | | x | | x | |
| Serum sample for SARS-CoV-2 anti-S antigen assessments | x (predose) | | x (predose) | | x | x | x | x |
| **Safety** | | | | | | | | |
| Local and systemic predefined solicited AEs (recorded daily by participant in Solicited AE e-Diary) | x (through Day 8) | | x (through Day 29) | | | | | |
| AEs recording | x | x | x (through Day 50) | | | | | |
| SAEs, MAAEs, and AESIs | x | x | x | x | x | x | x | x |
| Blood sample (hematology and [illegible] | x | | | | x | | | |
| Telephone contact for safety monitoring | | x | | x | | | | |

This document is confidential.

[a] Not a study site visit; participants will be contacted by telephone for safety monitoring.

[b] If urine tests positive or indeterminate, a quantitative serum β-HCG will be performed for confirmation.

[c] Weekly contact with participants to remind them to present to the study site for SARS-CoV-2 testing if they have qualifying symptoms.

[d] Single dose arms receive placebo at day 22

AE = adverse event; AESI = adverse events of special interest; CoV = coronavirus; COVID-19 = coronavirus disease 2019; MAAE = medically attended adverse event; NA = Not applicable; nAb = neutralizing antibody; PBMC = peripheral blood mononuclear cell; RT-PCR = reverse transcriptase polymerase chain reaction; SAE = serious adverse event; SARS-CoV-2 = severe acute respiratory syndrome-coronavirus-2; β-hCG = beta-human chorionic gonadotropin.

This document is confidential.

*Table 3 Treatment and Follow-up Period – Main Study (Excluding Phase II Participants)*

| Procedure | Treatment and follow-up | | | | | | | |
|---|---|---|---|---|---|---|---|---|
| **Day** | **1** | **8** [a] | **22**[c] | **29** [a] | **36** | **57** | **180** | **360** |
| **Window (days)** | **NA** | **± 1** | **± 2** | **± 2** | **±2** | **± 10** | **± 15** | **± 30** |
| Medical history | x | | | | | | | |
| Targeted physical examination | x | | | | | | | |
| Vital signs (including pulse oximetry) | x | | | | | | | |
| Pregnancy test – urine or serum [b] | x (predose) | | x (predose) | | | | | |
| Concomitant medications | x | x | x | x | x | As applicable, for treatment of SAEs, MAAEs, or AESIs | | |
| Verify eligibility criteria | x | | | | | | | |
| **Study intervention administration** | x | | x [c] | | | | | |
| **Efficacy assessments** | | | | | | | | |
| Weekly telephone/email/text contacts - monitoring for COVID-19 qualifying symptoms | Weekly throughout this period | | | | | | | |
| Serum sample for SARS-CoV-2 serology testing for abs anti-N antigen | x (predose) | | x | | x | x | x | x |
| Nasal swab for SARS-CoV-2 RT-PCR (local laboratory) (predose) | x (predose) | | x (predose) | | | | | |
| **Immunogenicity assessments** | | | | | | | | |
| Serum sample storage for exploratory assessments (left over aliquot after anti-N testing) | x (predose) | | x | | x | x | x | x |

This document is confidential.

| Safety | | | | | | | | |
|---|---|---|---|---|---|---|---|---|
| AEs | x | x | x (through Day 50) | | | | | |
| SAEs, MAAEs, and AESIs | x | x | x | x | x | x | x | x |
| Telephone contact for safety monitoring | | x | | x | | | | |

[a] Not a study site visit; participants will be contacted by telephone for safety monitoring.

[b] If urine tests positive or indeterminate, a quantitative serum β-HCG will be performed for confirmation.

[c] To be done only if double dose regimen is selected.

AE = adverse event; AESI = adverse events of special interest; NA= Not applicable; CoV = coronavirus; COVID-19 = coronavirus disease 2019; MAAE = medically attended adverse event; RT-PCR = reverse transcriptase polymerase chain reaction; SAE = serious adverse event; SARS-CoV-2 = severe acute respiratory syndrome-coronavirus-2; β-hCG = beta-human chorionic gonadotropin

This document is confidential.

*Table 4 Illness Visits (Participants with Qualifying Clinical Symptoms)*

| **Procedure** | **Site Visit** | **Home Collection by Participant** | | **Site/Remote Visit for SARS-CoV-2 Positive Participants Only** | | |
|---|---|---|---|---|---|---|
| **Day** | **1** | **3** | **8** | **14** | **21** | **28** |
| **Window (days)** | **NA** | **± 1** | **± 2** | **± 2** | **± 2** | **± 2** |
| Brief physical examination | x | | | | | |
| Vital signs (including pulse oximetry) | x | | | x | x | x |
| Concomitant medication | Throughout this period | | | | | |
| **Efficacy assessments** | | | | | | |
| Digital medical device | Daily throughout this period | | | | | |
| Symptoms associated with COVID-19 (recorded daily by participant in Illness e-Diary) | Daily throughout this period | | | | | |
| **Virology assessments** | | | | | | |
| Nasal swab for SARS-CoV-2 RT-PCR (local laboratory) | x | | | | | |
| SARS-CoV-2 RT-PCR (central laboratory) | x | | | x | x | x |
| **Immunogenicity assessments** | | | | | | |
| Serum sample for SARS-CoV-2 nAbs assessment (central laboratory) | x | | | x | | x |
| Serum sample for exploratory assessments (central laboratory) | x | | | x | | x |
| **Safety assessments** | | | | | | |
| SAEs, MAAEs, and AESIs | Daily throughout this period | | | | | |
| Telephone contact for safety monitoring | | x | x | | | |

This document is confidential.

Following availability of the SARS-CoV-2 RT-PCR results, only participants who test positive will continue with the illness visits, including any home collection requirements. Participants who test negative for SARS-CoV-2 will be instructed to stop all illness visit assessments and return the digital medical device (see Section 12.1.2).

AESI = adverse events of special interest; NA= Not applicable; CoV = coronavirus; COVID-19 = coronavirus disease 2019; MAAE = medically attended adverse event; nAb = neutralizing antibody; RT-PCR = reverse transcriptase polymerase chain reaction; SAE = serious adverse event; SARS-CoV-2 = severe acute respiratory syndrome-coronavirus-2.

This document is confidential.

13.5.3.1 Analysis for Deciding Phase III Expansion and Dose Selection in Phase II Part of the Study....79
13.5.3.2 Primary Efficacy Analysis ....104
13.5.3.3 Key Secondary Efficacy Endpoint ....81
13.5.3.4 Other Secondary Efficacy Endpoints ....106
13.7 Sensitivity Analyses ....88
13.8 Subgroup Analyses....111
13.9 Interim Analysis ....8111
13.10 Unblinding for Interim Analyses and Responsibility for Analyses....88
13.11 Data Monitoring Committee....113

This document is confidential.

#### 4.1 List of In-text Tables

Table 17 Appendix 3 – Table for Clinical Abnormalities: Systemic (General or Illness)..130

#### 4.2 List of In-text Figures

Figure 1 Study Flow Diagram ..........................................................................................62

### 5 LIST OF ABBREVIATIONS

| Abbreviation | Definition |
|---|---|
| AE | adverse event |
| AESI | adverse event of special interest |
| ALT | alanine aminotransferase |
| AST | aspartate aminotransferase |
| β-hCG | beta-human chorionic gonadotropin |
| CDC | Centers for Disease Control and Prevention |
| CI | confidence interval |
| CoV | coronavirus |
| COVID-19 | coronavirus disease 2019 |
| CRO | contract research organization |
| CSR | clinical study report |
| DBL | database lock |
| DSMB | Data Safety Monitoring Board |
| E | envelope |
| eCRF | electronic case report form |
| EDC | electronic data capture |
| e-Diary | electronic diary |
| ELISA | enzyme-linked immunosorbent assay |
| ELISpot | enzyme-linked immunospot |
| FIH | first-in-human |
| GCP | Good Clinical Practice |
| GMFR | geometric mean fold rise |
| GMT | geometric mean titer |
| IB | Investigator's Brochure |
| ICF | informed consent form |
| ICH | International Council for Harmonisation |
| IEC | Independent Ethics Committee |
| IgA | immunoglobulin A |
| IgG | immunoglobulin G |
| IgM | immunoglobulin M |
| IM | intramuscular |
| IMP | investigational medicinal product |
| IND | Investigational New Drug |
| IRB | Institutional Review Board |

This document is confidential.

| Abbreviation | Definition |
|---|---|
| IRT | Interactive Response Technology |
| IV | intravenous |
| M | membrane |
| MAAE | medically attended adverse event |
| MERS | Middle East respiratory syndrome |
| MERS-CoV | Middle East respiratory syndrome coronavirus |
| MSD | Meso Scale Discovery |
| NHP | non-human primate |
| PBMC | peripheral blood mononuclear cell |
| PSRT | Protocol Safety Review Team |
| RBD | receptor binding domain |
| RRR | relative risk reduction |
| RT-PCR | reverse transcriptase polymerase chain reaction |
| S | Spike |
| SAE | serious adverse event |
| SAP | Statistical Analysis Plan |
| SARS-CoV | severe acute respiratory syndrome-coronavirus |
| SARS-CoV-2 | severe acute respiratory syndrome-coronavirus 2 |
| SoA | Schedule of Activities |
| TBL | total bilirubin |
| ULN | upper limit of normal |
| US FDA | United States Food and Drug Administration |
| USA | United States of America |
| VE | vaccine efficacy |
| vp | viral particles |
| WHO | World Health Organization |
| w/v | weight/volume |

This document is confidential.

### 6 INTRODUCTION

GRAd-COV2 is being developed for the prevention of COVID-19. GRAd-COV2 is a replication-defective gorilla adenoviral vector (GRAd) encoding the SARS-CoV-2 surface glycoprotein (S, Spike) antigen under the control of CMV immediate early promoter. The encoded Spike antigen is stabilized in pre-fusion conformation by introducing 2 proline residues [1].

#### 6.1 Background

In December 2019, an outbreak of febrile syndrome associated to pneumonia of unknown origin occurred in Wuhan, Hubei district. Soon after in January 2020 the causative agent was isolated, sequenced and confirmed to be a novel betacoronavirus. The virus was named SARS-CoV-2 due to the high similarity with the previously emerged SARS-CoV that caused a global epidemic in 2002-2003 with more than 8000 cases and 774 confirmed deaths. The SARS-CoV-2 associated disease was named Coronavirus disease 2019, COVID-19. SARS-CoV-2 is the third novel betacoronavirus in the last 20 years to cause substantial human disease; however, unlike its predecessors SARS-CoV and MERS-CoV, SARS-CoV-2 transmits efficiently from person-to-person [2, 3]. Indeed, the infection spread rapidly worldwide and on 11 March 2020 the WHO officially declared it a pandemic. As of December 2020, the COVID-19 confirmed cases are well beyond 70 million worldwide, with >1.6 million deaths reported globally [4]. Europe has suffered more than 21 million cases, while USA alone are approaching 16 million cases.

Different countries and geographical areas have reacted differently to control viral transmission during the initial and second pandemic wave, going from strict lockdown to more relaxed forms of social distancing, but clearly the pandemic has caused major disruption of everyday life and seriously impacted the global economy.

Despite enormous efforts, no specific antiviral or therapeutic strategy has yet been approved to treat the COVID-19 disease, and the development of vaccines is regarded as a global priority and the only mean to bring the pandemic under control in a reasonable timeframe.

Like other coronaviruses, SARS-CoV-2 has four structural proteins, known as the S (Spike), E (envelope), M (membrane), and N (nucleocapsid) proteins; the N-protein holds the RNA genome, and the S, E, and M proteins together create the viral envelope.

The S-protein has a pivotal central role in viral infection and pathogenesis. Specifically, the S1 subunit holds a receptor-binding domain (RBD; residues 331–524), which mediates viral binding to functional ACE2 receptors on susceptible cells and is the main target for SARS-CoV-2 neutralizing antibodies [5, 6]. Consequently, Spike and its RBD domains were immediately identified as the main target antigen for inclusion in all candidate vaccines.

The effort for vaccine development has been exceptional and unprecedented, with more than 52 vaccines currently in clinical testing as of December 10 2020 [7], and 164 under preclinical evaluation. Alongside with more classical vaccine approaches, such as inactivated SARS-CoV-2 or recombinant Spike protein

This document is confidential.

trimers in adjuvant, the pandemic has provided the ideal validation arena for novel genetic vaccine approaches based either on viral vectors or directly on nucleic acids, namely RNA or DNA.

Genetic vaccine platform has many attracting features in facing outbreaks, such as flexibility and speed of construction, easily scalable manufacturing processes and intrinsic safety in immunocompromised. Amongst the most promising and clinically advanced genetic vaccine approaches undertaken in the global SARS-CoV-2 vaccine search are adenoviral vectors.

Replication-defective adenoviruses are among the most potent vectors for induction of antibody and T cell responses to encoded antigens [8]. To overcome the issue of pre-existing immunity in human population, simian adenoviral vectors were explored clinically as an alternative. Simian adenoviruses derived from Chimpanzee, Bonobo and Gorilla, which are not known to infect or cause pathological illness in humans, consequently, have low/no seroprevalence (0%-18%) in the human population. Prior studies in thousands of human volunteers using simian adenoviral vaccine vectors encoding different antigens (relevant to Ebola, malaria, Hepatitis-C, HIV, and RSV), have shown that this vaccine platform is safe and can generate potent, durable, and high-quality T-cell and antibody responses [8-18].

The Sponsor has isolated a novel, proprietary adenovirus strain from gorilla (GRAd32), and a replication-defective derived vector (GRAd) was engineered to encode pre-fusion stabilized SARS-CoV-2 Spike.

Immunogenicity studies in mice and macaques showed that a single intramuscular administration of the vaccine induced binding and neutralizing antibodies to SARS-CoV-2, along with potent, Th1 skewed CD4 and CD8 T cell responses. In a macaque model of SARS-CoV-2 challenge, GRAd-COV2 vaccination substantially reduced viral replication in the Broncho alveolar fluid of vaccinated animals compared to controls. Additional efficacy study is underway in the Syrian Hamster model.

The vaccine was administered in 91 healthy volunteers in a first in human phase 1 clinical trial (NCT04528641) as a single intramuscular injection. The study evaluated three escalating doses of GRAd-COV2 in two age cohorts: younger (18-55 years) and older (65-85 years) healthy adults. Interim analysis in younger adults up to 4 weeks of safety and immunogenicity demonstrated that the vaccine was well tolerated at all three doses ($5x10^{10}$, $1x10^{11}$ and $2x10^{11}$ viral particles). Vaccination induced Spike and RBD binding antibodies in all subjects, as measured with a validated diagnostic CLIA assay (DiaSorin), with robust ELISA titers in the range of those measured in COVID-19 convalescent patients. Neutralizing activity was detected in 42-66% of volunteers as measured by microneutralization assay (MNA90) and in 87-100% by plaque-reduction neutralization test (PRNT50). GRAd-COV2 vaccination at all three doses induced as well potent IFNγ secreting Spike specific T cell responses with similar frequencies of both Spike-specific CD4 and CD8.

Preliminary observation suggest that the safety profile and immunogenicity is similar in older subjects.

See the GRAd-COV2 IB, Sections 5 and 6 for additional information on nonclinical and clinical studies, respectively. Detail on the development and chemistry of GRAd-COV2 is provided in the IMPD v2.0 (03 December 2020). Overall, the preliminary data from the GRAd-COV2 clinical and nonclinical studies support further development of GRAd-COV2 for the prevention of COVID-19.

This document is confidential.

#### 6.2 Clinical Risks/Benefits of GTRAd-COV2

More detailed information about the known and expected benefits and potential risks of GRAd-COV2 can be found in the GRAd-COV2 IB.

Based on preliminary clinical data from Study RT-CoV-2 (NCT04528641), the most common local solicited AEs was injection site pain. Common systemic solicited AEs in the study included fever, fatigue, headache and malaise. The majority of events were mild or moderate in severity and resolved within 1 to 7 days (Section 2.2 and GRAd-COV2 IB).

There are no identified risks for GRAd-COV2. Important potential risks are hypersensitivity including anaphylaxis/anaphylactic reactions, vaccine-associated enhanced respiratory disease, and Guillain-Barré syndrome and other immune-mediated reactions.

Serious allergic reactions including anaphylaxis may occur, as with any vaccine. Acute allergic reactions may include cardio-respiratory, skin, and gastrointestinal signs and symptoms, such as hypotension, bronchospasm, angioedema, urticaria, and diarrhea.

Vaccine-associated enhanced respiratory disease is a potential risk for GRAd-COV2. The risks of inducing disease enhancement and lung immunopathology in the event of COVID-19 following GRAd-COV2 vaccination are unknown.

As with many vaccines, temporary ascending paralysis (Guillain-Barré syndrome) or other immune-mediated reactions that can lead to organ damage may occur, but this should be extremely rare.

Recipients of GRAd-COV2 do not have any guaranteed benefit, however, GRAd-COV2 may be efficacious and offer participants protection from COVID-19. The information gained from this study will inform development decisions.

For the safety of participants, the protocol has incorporated various risk mitigation measures including appropriate inclusion and exclusion criteria, close monitoring of participants, and stopping criteria. An independent DSMB will provide study oversight, evaluating cumulative safety and other clinical data at regular intervals. Taking these measures into account, the potential risks identified in association with GRAd-COV2 are justified by the anticipated benefit that may be afforded to participants for the prevention of COVID-19.

#### 6.3 Study Rationale

The aim of the study is to assess the safety, efficacy, and immunogenicity of GRAd-COV2 for prevention of COVID-19. The COVID-19 pandemic has caused major disruption to healthcare systems with significant socioeconomic impacts. Currently, there are no licensed treatments available against COVID-19 and accelerated vaccine development has been pursued at unprecedented speed and (public and private) level of financial investment. Safe and effective vaccines for COVID-19 prevention would have significant global public health impact. While few candidate vaccines have now reached approval or emergency use authorization in different Countries and regions, additional successful vaccines are envisaged to be

This document is confidential.

needed to increase dose production capacity and for rapid and full coverage of the susceptible world population.

This document is confidential.

### 7 STUDY OBJECTIVES AND ENDPOINTS

| **Objective** [a] | **Estimand** [b] **Description/Endpoint** |
|---|---|
| **Primary for Phase III part of the Study** | |
| 1) To estimate the efficacy of a single or repeated (21 days apart) IM dose of GRAd-COV2 compared to placebo for the prevention of COVID-19 in adults ≥ 18 years of age | **Population:** Full analysis set, excluding participants who are seropositive at baseline. |
| | **Endpoint:** A binary response, whereby a participant is defined as a COVID-19 case if their first case of SARS-CoV-2 RT -PCR-positive symptomatic illness occurs ≥ 28 days post first dose or ≥ 7 days after the second dose of study intervention. Otherwise, a participant is not defined as a COVID-19 case. For the phase II participants, since all of them receive two doses, cases will be considered valid if they occur ≥ 7 days post second dose. For the phase III participants, the two described conditions will apply depending on the schedule selected in phase II, ie, if the single dose is selected, cases will be considered valid if they occur ≥28 days post dose; if the double dose is selected, cases will be considered valid if they occur ≥ 7 days post second dose. The endpoint will be treated as time to event. |
| | **Intercurrent events**: The intercurrent events (i.e. withdrawals before having met the primary efficacy endpoint, deaths for reasons that are unrelated to COVID-19 and COVID-19 infections that occur before 28 days post first dose or before 7 days post second dose) will be handled using a “while on treatment” strategy, where the time to COVID-19 is censored at the date of withdrawal or death or early COVID-19 infection. In case the double-dose schedule is selected for phase III, subjects missing the second dose will be excluded from the primary analysis. |
| | **Summary measure:** VE, calculated as 1-HR, where HR (=Hazard Ratio) between GRAd-COV2 and placebo is estimated with a Cox proportional hazard model. |
| 2) To assess the safety and tolerability of a single or repeated (21 days apart) IM dose of GRAd-COV2 compared to placebo in adults ≥ 18 years of age | a) Incidence of AEs for 28 days post each dose of study intervention |
| | b) Incidence of SAEs, MAAEs, and AESIs from Day 1 post treatment through Day 360 |
| **Primary for phase II part of the Study** | |
| 1) To assess the reactogenicity of a single or repeated (21 day apart) IM dose of GRAd-COV2 compared to placebo in adults ≥ 18 years of age | Incidence of local and systemic solicited AEs for 7 days post each dose of study intervention |

This document is confidential.

| Objective [a] | Estimand [b] Description/Endpoint |
|---|---|
| 2) To identify the schedule to be further studied in phase III by assessing antibody responses to S antigen following a single or repeated (21 days apart) IM dose of GRAd-COV2 or placebo (first 450 participants) | a) Post-treatment GMTs and GMFRs from day of dosing baseline value to 35 days post first dose (14 days after second dose) in SARS-CoV-2 S and/or RBD antibodies. |
| | b) Proportion of participants who have a post-treatment seroresponse (≥ 4-fold rise in titers from day of dosing baseline value to 35 days post first dose, i.e. 14 days after second dose) to the S and/or RBD antigens of GRAd-COV2. |
| **Key Secondary (study phase indicated in parenthesis)** | |
| 3) To estimate the efficacy of 1 single or repeated (21 days apart) IM dose of GRAd-COV2 compared to placebo for the prevention of severe or critical symptomatic COVID-19 (Phase III) | Time to first SARS-CoV-2 RT-PCR-positive severe or critical symptomatic illness occurring ≥ 28 days post first dose or ≥ 7 days after second dose of study intervention. |
| **Other Secondary (study phase indicated in parenthesis)** | |
| 1) To estimate the efficacy of a single or repeated (21 days apart) IM dose of GRAd-COV2 compared to placebo for the prevention of SARS-CoV-2 infection - Asymptomatic cases (Phase III) | Proportion of participants who have a post-treatment response (negative at baseline to positive post treatment with study intervention) for SARS-CoV-2 Nucleocapsid antibodies over time. |
| 2) To estimate the efficacy of a single or repeated (21 days apart) IM dose of GRAd-COV2 compared to placebo for the prevention of symptomatic COVID-19 using CDC criteria (Phase III) | Time to first case of SARS-CoV-2 RT-PCR- positive symptomatic illness occurring ≥ 28 days post first dose or ≥ 7 days after second dose of study intervention using CDC criteria. |
| 3) To estimate the efficacy of 1 single or repeated (21 days apart) IM dose of GRAd-COV2 compared to placebo for the prevention of COVID-19-related Emergency Department visits (Phase III) | Time to first COVID-19-related Emergency Department admission occurring ≥ 28 days post first dose or ≥ 7 days post second dose of study intervention. |
| 4) To evaluate VE to prevent death caused by COVID-19 (Phase III). | Time to COVID-19 related deaths occurring ≥ 28 days post first dose or ≥ 7 days post second dose of study intervention. |
| 5) To assess antibody responses to S antigen following a single or repeated (21 days apart) IM dose of GRAd-COV2 or placebo (Phase II and Illness Visits only) | a) Post-treatment GMTs and GMFRs from day of dosing baseline value to 35 days post first dose (14 days post second dose) in SARS-CoV-2 S and/or RBD antibodies. |
| | b) The proportion of participants who have a post-treatment seroresponse (≥ 4-fold rise in titers from day of dosing baseline value to 35 days post first dose, i.e 14 days post second dose) to the S and/or RBD antigens of GRAd-COV2. |

This document is confidential.

| **Objective**[a] | **Estimand**[b] **Description/Endpoint** |
|---|---|
| 6) To determine anti-SARS-CoV-2 neutralizing antibody levels in serum following 1 single or repeated (21 days apart) IM dose of GRAd-COV2 or placebo (Phase II and Illness Visits only) | a) Post-treatment GMTs and GMFRs from day of dosing baseline value to 35 days post first dose (14 days post second dose) in SARS-CoV-2 neutralizing antibodies. |
| | b) Proportion of participants who have a post-treatment seroresponse (≥ 4-fold rise in titers from day of dosing baseline value to 35 days post first dose, i.e. 14 days post second dose) to GRAd-COV2 as measured by SARS-CoV-2 neutralizing antibodies. |

[a] Illness Visits: Participants who present with qualifying symptoms will be tested for SARS-CoV-2 and if positive, will complete illness visits.

[b] Estimand is the target of estimation to address the scientific question of interest posed by the primary objective. Attributes of an estimand include the population of interest, the variable (or endpoint) of interest, the specification of how intercurrent events are reflected in the scientific question of interest, and the population-level summary for the variable.

AE = adverse event; AESI = adverse event of special interest; CDC = Centers for Disease Control and Prevention; COVID-19 = coronavirus disease 2019; ELISpot = enzyme-linked immunospot; GMFR = geometric mean fold rise; GMT = geometric mean titer; IFN-γ = interferon-gamma; IM = intramuscular; MAAE = medically attended adverse event; MSD = Meso Scale Discovery; NP = nasopharyngeal; RT-PCR = reverse transcriptase polymerase chain reaction; qRT-PCR = quantitative reverse transcriptase polymerase chain reaction, RBD = receptor binding domain; S = Spike; SAE = serious adverse event; SARS-CoV-2 = severe acute respiratory syndrome- coronavirus-2; VE = vaccine efficacy.

This document is confidential.

### 8 INVESTIGATIONAL PLAN

#### 8.1 Description of Overall Study Design and Plan

RT-CoV-2_01 is a Phase II/III randomized, observer-blind, placebo-controlled multicenter international study assessing the safety, efficacy, and immunogenicity of GRAd-COV2 compared to saline placebo for the prevention of COVID-19. Participants will be adults ≥ 18 years of age who are healthy or have medically-stable chronic diseases. In the phase II part approximately 900 participants will be randomized in a 1:1:1 ratio to receive one single IM dose of either $2x10^{11}$ vp GRAd-COV2 (n = approximately 300) plus 1 dose of saline placebo after 21 days or a repeated (21 days apart) IM dose of $1x10^{11}$ vp GRAd-COV2 vp (n= approximately 300) or two doses of saline placebo (n = approximately 300) on Day 1 and 22. Randomization will be stratified based on age and, if they are < 65 years of age, based on the presence or absence of risk factors for severe illness from COVID-19 based on CDC recommendation as of May 2020 [19]. There will be 3 strata for randomization: ≥ 65 years, < 65 years and categorized to be at increased risk ("at risk") for the complications of COVID-19, and < 65 years "not at risk". Risk will be defined based on the study participants' relevant past and current medical history. At least 25% of enrolled participants, will be either ≥ 65 years of age or < 65 years of age and "at risk" at Screening.

Participants who are < 65 years old will be categorized as at risk for severe COVID-19 illness if they have at least 1 of the following risk factors at Screening:

- Chronic lung disease (e.g. emphysema and chronic bronchitis, idiopathic pulmonary fibrosis, and cystic fibrosis) or moderate to severe asthma
- Significant cardiac disease (e.g. heart failure, coronary artery disease, congenital heart disease, cardiomyopathies, and pulmonary hypertension)
- Severe obesity (body mass index ≥ 40 kg/m2)
- Diabetes (Type 1, Type 2)
- Liver disease
- Human Immunodeficiency Virus (HIV) infection

An independent DSMB will provide oversight, to ensure safe and ethical conduct of the study and, together with a Steering Committee, based on safety (900 participants 1 week after dosing) and immunogenicity (450 participants at week 5 after the first dosing) data generated in phase II part, will recommend the expansion to phase III and the best regimen to be used.

Once the phase III expansion is granted, according to the epidemic evolution and the availability on the market of alternative vaccine(s) and the characteristics of vaccination campaign, the phase III study design will be adapted following these 3 potential scenarios:

1) a superiority trial vs placebo on the overall population;

2) a superiority trial vs placebo on a population subset (low risk subjects as for the infection outcome and so with a low priority of receiving a new vaccine);

3) a non-inferiority trial vs the available alternative vaccine on a surrogate endpoint, if available.

In case of scenarios 1 and 2, additional 9400 participants will enter the study for the evaluation of safety and efficacy. However, at the time of commencing phase III, the assumptions on which current estimates of sample size are based might have been changed and so sample size might need re-computation.

At present, scenario 3 is not evaluated in terms of sample size, because the assumptions for these computations are not known. If scenario 3 is to be followed due to ethic issues about placebo use, a protocol amendment will be submitted to define the sample size and the other protocol details.

Participants who present with at least one of the qualifying symptoms listed below through Day 360 will be assessed for COVID-19. With the exception of fever, shortness of breath, or difficulty breathing, the symptom must be present for 2 or more days. Participants with a COVID-19 qualifying symptom(s) will be tested for SARS-CoV-2, and if positive will complete illness visit assessments, as presented in Table 4. See Section 12.1 for details on COVID-19 assessments.
The Study Flow Diagram is presented in Figure 1.

This document is confidential.

*Figure 1 Study Flow Diagram*

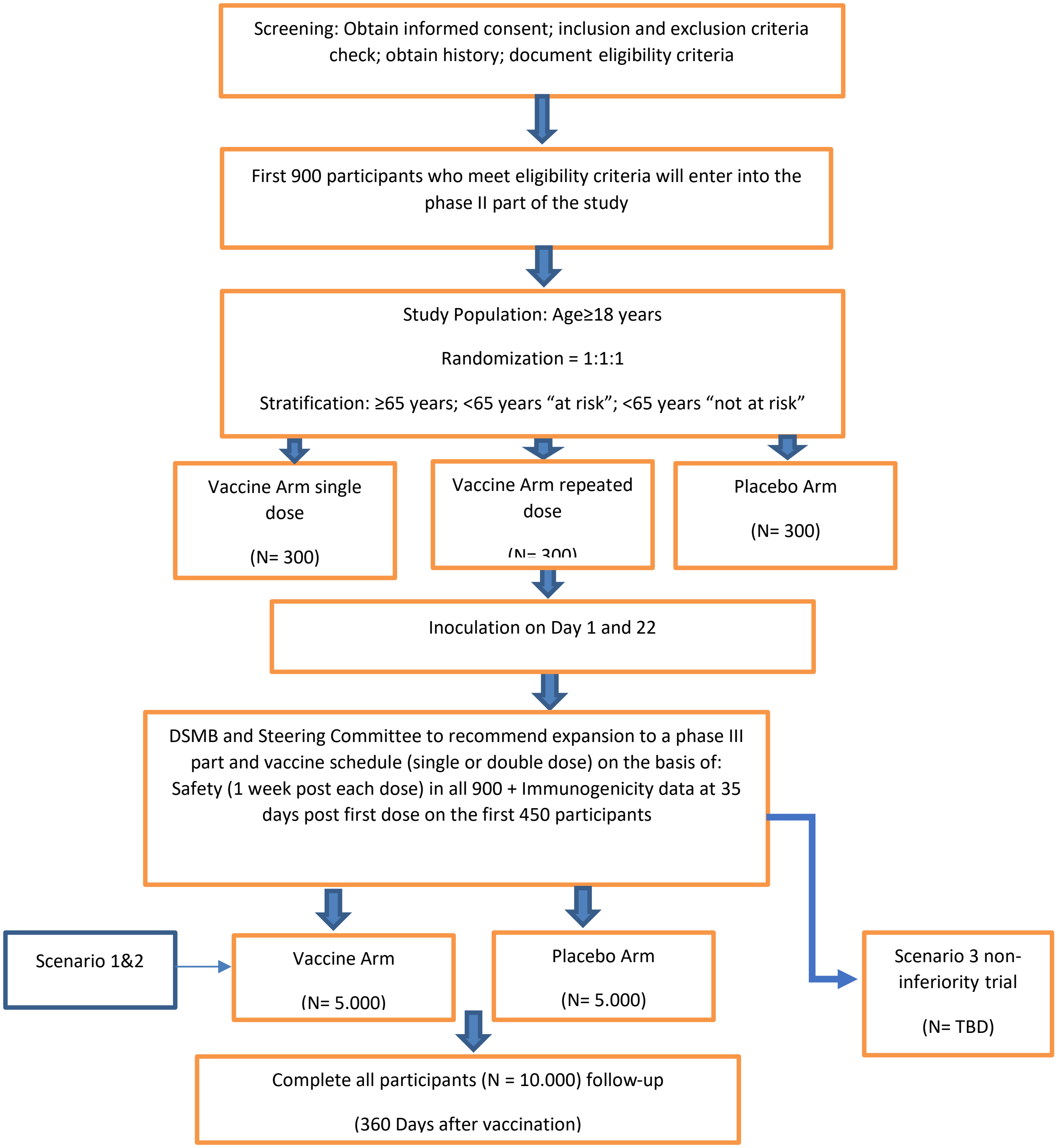

This document is confidential.

#### 8.2 Discussion of Study Design

The participant population is male and female adults ≥ 18 years of age. Inclusion of older adults is based on data that are being gathered from the ongoing phase I study sponsored by ReiThera. Study RT-CoV-2 enrolled 45 adults 18 to 55 years of age and enrolled 45 adults who are 65 to 85 years of age.

Adults with medically-stable chronic diseases may participate if, according to the judgement of the investigator, hospitalization within the study period is not anticipated and the participant appears likely to be able to remain on study through the end of protocol-specified follow-up.

The study will exclude females who are pregnant or breast-feeding and individuals less than 18 years of age. Women who are pregnant or breast-feeding are excluded at this point as nonclinical developmental and reproductive toxicity studies to support vaccinating these individuals have yet to be performed. Additionally, it is planned that children and adolescents will be evaluated for their response to the vaccine once safety and efficacy have been established in adults.

Participants who have been previously diagnosed with laboratory-confirmed SARS-CoV-2 infection are excluded from study participation. Participants who will result positive to SARS-CoV-2 infection before the second dose injection will not receive that dose, but will continue to be followed-up according to the study protocol. Participants with previous undiagnosed infection are not excluded. Participant's baseline serostatus will be determined but baseline serostatus will not be used as a basis for exclusion from the study. Participants who are seropositive at baseline are enrolled in order to gather safety data in this group as it is anticipated that, if proven to be efficacious, the vaccine will rapidly be distributed to millions of individuals and that these individuals will not be tested for serologic evidence of previous infection prior to vaccination. Participant's baseline serostatus will be determined so that subgroup analyses for both safety and efficacy can be performed by baseline serostatus.

The study population represents the initial target population for GRAd-COV2. If GRAd-COV2 demonstrates efficacy for the prevention of COVID-19, the safety and immunogenicity of the vaccine in additional groups such as pregnant women, and children and adolescents may be assessed in future studies.

#### 8.3 End of Study

A participant is considered to have completed the study if he/she has completed all phases of the study including the last scheduled procedure shown in the SoA (Section 3).

The end of the study is defined as the date of the last scheduled procedure shown in the SoA (Section 3) for the last participant in the study globally.

This document is confidential.

### 9 SELECTION OF STUDY POPULATION

Prospective approval of protocol deviations to recruitment and enrollment criteria, also known as protocol waivers or exemptions, is not permitted.

#### 9.1 Inclusion Criteria

Participants are eligible to be included in the study only if all of the following criteria apply:

1. Adult, ≥ 18 years of age at the time of consent
2. Medically stable such that, according to the judgment of the investigator, hospitalization within the study period is not anticipated and the participant appears likely to be able to remain on study through the end of protocol-specified follow-up
   – A stable medical condition is defined as disease not requiring significant change in therapy or hospitalization for worsening disease during the 3 months prior to enrollment
3. Able to understand and comply with study requirements/procedures (if applicable, with assistance by caregiver, surrogate, or legally authorized representative) based on the assessment of the investigator
4. Contraceptive use by women should be consistent with local regulations regarding the methods of contraception for those participating in clinical studies
5. Female participants
   (a) Women of childbearing potential must:
   - Have a negative pregnancy test on the day of screening and on Day 1
   - Use one highly effective form of birth control for at least 28 days prior to Day 1 and agree to continue using one highly effective form of birth control through 60 days following administration of the dose of study intervention.
     A highly effective method of contraception is defined as one that can achieve a failure rate of less than 1% per year when used consistently and correctly (see Table 5). Periodic abstinence, the rhythm method, and withdrawal are NOT acceptable methods of contraception.

   (b) Women are considered of childbearing potential unless they meet either of the following criteria:

   - Surgically sterilized (including bilateral tubal ligation, bilateral oophorectomy, or hysterectomy), or
   - Post-menopausal

  - For women aged < 50 years, post-menopausal is defined as having both:
    - A history of ≥ 12 months amenorrhea prior to randomization, without an alternative cause, following cessation of exogenous sex-hormonal treatment, and
    - A follicle-stimulating hormone level in the post-menopausal range until follicle-stimulating hormone is documented to be within menopausal range, the participant is to be considered of childbearing potential
- For women aged ≥ 50 years, post-menopausal is defined as having a history of ≥ 12 months amenorrhea prior to randomization, without an alternative cause, following cessation of exogenous sex-hormonal treatment

*Table 5 Highly Effective Methods of Contraception*

| Barrier Methods | Hormonal Methods |
|---|---|
| • Intrauterine device<br>• Intrauterine hormone-releasing system (IUS) [a]<br>• Bilateral tubal occlusion<br>• Vasectomized partner [b]<br>• Sexual abstinence [c] | • Combined (estrogen- and progestogen-containing hormonal contraception)<br>◦ Oral (combined pill)<br>◦ Injectable<br>◦ Transdermal (patch)<br>• Progestogen-only hormonal contraception<br>◦ Oral<br>◦ Injectable<br>◦ Implantable |

a This is also considered a hormonal method

b Provided that partner is the sole sexual partner of the woman of childbearing potential study participant and that the vasectomized partner has received medical assessment of the surgical success

c Sexual abstinence is considered a highly effective method only if defined as refraining from heterosexual intercourse during the entire period of the study and if it is the preferred and usual lifestyle of the participant

6. Capable of giving signed informed consent as described in Appendix A, which includes compliance with the requirements and restrictions listed in the ICF and in this protocol.

#### 9.2 Exclusion Criteria

Participants are excluded from the study if any of the following criteria apply:

1. History of allergy to any component of the vaccine
2. History of Guillain-Barré syndrome or any other demyelinating condition

This document is confidential.

3. Significant infection or other acute illness, including fever >37,3 °C (> 99.14 °F) on the day prior to or day of randomization
4. History of laboratory-confirmed SARS-CoV-2 infection
5. Any confirmed or suspected immunosuppressive or immunodeficient state, including asplenia (only for phase II)
6. Recurrent severe infections and use of immunosuppressant medication within the past 6 months (≥ 20 mg per day of prednisone or its equivalent, given daily or on alternate days for ≥ 15 days within 30 days prior to administration of study intervention) (only for phase II) The following exceptions are permitted:
   – Topical/inhaled steroids or short-term oral steroids (course lasting ≤ 14 days)
   – Human immunodeficiency virus-positive stable participants on stable antiretroviral therapy
7. History of primary malignancy (only for phase II) except for:
   (a) Malignancy with low potential risk for recurrence after curative treatment (for example, history of childhood leukaemia) or metastasis (for example, indolent prostate cancer) in the opinion of the site investigator.
   (b) Adequately treated non-melanoma skin cancer or lentigo maligna without evidence of disease
   (c) Adequately treated uterine cervical carcinoma in situ without evidence of disease
   (d) Localized prostate cancer
8. Clinically significant bleeding disorder (eg, factor deficiency, coagulopathy, or platelet disorder), or prior history of significant bleeding or bruising following IM injections or venipuncture
9. Severe and/or uncontrolled cardiovascular disease, respiratory disease, gastrointestinal disease, liver disease, renal disease, endocrine disorder, and neurological illness, as judged by the Investigator (mild/moderate well-controlled comorbidities are allowed) (Only for phase II)
10. Any other significant disease, disorder, or finding that may significantly increase the risk to the participant because of participation in the study, affect the ability of the participant to participate in the study, or impair interpretation of the study data

This document is confidential.

11. Receipt of, or planned receipt of investigational or licensed products indicated for the treatment or prevention of SARS-CoV-2 or COVID-19
12. Receipt of any vaccine (licensed or investigational) other than licensed influenza vaccines within 30 days prior to and after administration of study intervention
13. Receipt of immunoglobulins and/or any blood products within 3 months prior to administration of study intervention or expected receipt during the period of study follow-up
14. Involvement in the planning and/or conduct of this study (applies to both Sponsor staff and/or staff at the study site)
15. For women only - currently pregnant (confirmed with positive pregnancy test) or breast-feeding
16. Has donated ≥ 450 mL of blood products within 30 days prior to randomization or expects to donate blood within 90 days of administration of the dose of study intervention

#### 9.3 Lifestyle Considerations

1 Participants must follow the contraception requirements outlined in Section 9.1

2 Restrictions relating to concomitant medications are described in Section 10.5

3 Agree to use digital medical device if diagnosed with COVID-19 as described in Section 12.1.2.2

#### 9.4 Screen Failures

Screen failures are defined as participants who consent to participate in the clinical study but are not subsequently randomly assigned to study intervention. A minimal set of screen failure information is required to ensure transparent reporting of screen failure participants to meet the Consolidated Standards of Reporting Trials publishing requirements and to respond to queries from regulatory authorities. Minimal information includes demography, screen failure details, eligibility criteria, and any SAEs.

Individuals who do not meet the criteria for participation in this study (screen failure) may be rescreened. Only a single rescreening is allowed in the study. Rescreened participants are required to sign a new ICF (Section 14.1.3), and will be assigned a new participant number.

This document is confidential.

### 10 STUDY INTERVENTION

In general, study intervention is defined as any investigational intervention(s), marketed product(s), or placebo intended to be administered to or medical device(s) utilized by a study participant according to the study protocol. For this study, study intervention is defined as GRAd-COV2 or saline placebo (Table 6). The medical device used for assessment of COVID-19 symptoms (ie, digital medical device [Section 12.1.2.2]) is not considered a study intervention.

#### 10.1 Study Interventions Administered

##### 10.1.1 Investigational Products

*Table 6 Investigational Products*

| Intervention Name | GRAd-COV2 | Placebo |
|---|---|---|
| **Type** | Vaccine | Placebo |
| **Dose Formulation** | 10 mM Tris, 75 mM NaCl, 1 mM MgCl2, 0.02% PS80, 5% sucrose, 0.1 mM EDTA, 10 mM Histidine, 0.5% ethanol, pH 7.4 | 0.9% (w/v) saline |
| **Unit Dose Strength(s)** | $2x10^{11}$ vp/mL | |
| **Dosage Level(s)** | $2x10^{11}$ vp as single administration<br>or $1x10^{11}$ vp as repeated administration 21 days apart | |
| **Route of Administration** | Intramuscular | Intramuscular |
| **Use** | Experimental | Placebo |
| **IMP and NIMP** | IMP | IMP |
| **Sourcing** | Provided centrally by the Sponsor | Provided by the Sponsor |
| **Packaging and Labeling** | Will be provided in vials within a carton. Each carton and vial will be labelled as required per country requirement | Will be provided in vials within a carton. Each carton will be labelled as required per country |
| **Current/Former Name or Alias** | Not applicable | Not applicable |

IMP = investigational medicinal product; NIMP = non-investigational medical product; vp = viral particles; w/v = weight/volume.

This document is confidential.

**GRAd-COV2**

The GRAd-COV-2 investigational vaccine is manufactured in formulation buffer as sterile, clear, colorless solution essentially free of particles. The vaccine is dispensed in a 3 mL borosilicate glass vial with a fill volume of 1.2 mL, closed by rubber stoppers and sealed with an aluminum flip-off cap.

Unopened vials of GRAd-COV2 vials must be stored at ≤-60°C (≤-76°F) for the duration of assigned shelf-life. GRAd-COV2 must be kept in original packaging until use to prevent prolonged light exposure.

**Placebo**

Commercially available 0.9% (w/v) saline for injection will be provided by the Sponsor and will be administered as placebo and used to dilute the investigational vaccine for the $1x10^{11}$ vp dose injection

###### 10.1.2 Dosage Instructions

Participants will receive a single dose of either $2x10^{11}$ vp GRAd-COV2 on day 1 plus saline placebo on day 22 or a repeated dose of $1x10^{11}$ vp on day 1 and 22 or saline placebo on day 1 and 22; the dose will be administered on Day 1 and 22 in phase II part of the study and on day 1 or day 1 and 22 in phase III (depending on the selected regimen) (see Table 2 and Table 3).

It is recommended that the study interventions be administered as an IM injection into the deltoid of the non-dominant arm. The second dose of IMP should be administered in the same arm as the first dose. Other injection sites may be used if necessary.

All study participants will be observed in the clinic for at least 30 minutes after vaccination.

Allergic reactions to vaccines are possible. Therefore, appropriate drugs and medical equipment to treat acute anaphylactic reactions must be immediately available, and study personnel must be trained to recognize and treat anaphylaxis.

#### 10.2 Preparation/Handling/Storage/Accountability

1. The investigator or designee must confirm appropriate temperature conditions have been maintained during transit for all study intervention received and any discrepancies are reported and resolved before use of the study intervention.
2. Only participants enrolled in the study may receive study intervention and only authorized site staff may supply or administer study intervention. All study intervention must be stored in a secure, environmentally controlled, and monitored (manual or automated) area in accordance with the labelled storage conditions with access limited to the investigator and authorized site staff.
3. The investigator, institution, or the head of the medical institution (where applicable) is responsible for study intervention accountability, reconciliation, and record maintenance (ie, receipt, reconciliation, and final disposition records).

This document is confidential.

4. Further guidance and information for the final disposition of unused study interventions are provided in the Pharmacy Manual or specified handling instructions.

##### 10.2.1 Dose Preparation and Administration

###### 10.2.1.1 GRAd-COV2

Dose of GRAd-COV2 must be prepared by the unblinded pharmacist (or designee in accordance with local and institutional regulations) using aseptic technique. Each dose is prepared in the following way:

- For $2x10^{11}$ vp (single dose regimen) withdrawing 1 mL volume from a vial of GRAd-COV2 in a sterile syringe
- For $1x10^{11}$ vp (double dose regimen) withdrawing 1.2 mL volume from a vial of saline solution in a sterile syringe; injecting the 1.2 mL of saline solution into the vial of GRAd-COV2; gently mixing the vial and withdrawing 1 mL volume from the diluted vial of GRAd-COV2 in a sterile syringe. Each vaccine vial may therefore provide up to two $1x10^{11}$ vp doses.

GRAd-COV2 does not contain preservatives. Each vial must be assigned a beyond-use-time of 4 hours from removal from the freezer of the GRAd-COV2 vial, after which any unused portion must be discarded according to a GMO wasting procedure

Once a GRAd-COV2 dose is drawn into a syringe for administration, the dose must be administered within the beyond-use-time of the vial. If GRAd-COV2 dose administration is not completed within the 4-hour vial beyond-use-time, a new dose must be prepared from a new vial.

Each vial of GRAd-COV2 has a label-claim volume of 1.2 mL.

###### 10.2.1.2 Placebo

Doses of placebo must be prepared by the unblinded pharmacist (or designee in accordance with local and institutional regulations) using aseptic technique. Each placebo dose is prepared by withdrawing 1 mL from a 0.9% (w/v) saline vial in a sterile syringe.

Saline (0.9% [w/v]) does not contain preservatives. Each vial must be assigned a beyond use time of 4 hours from first needle puncture, after which any unused portion must be discarded.

Once a placebo dose is drawn into a syringe for administration, the dose must be administered within the beyond-use-time of the vial. If placebo dose administration is not completed within the 4-hour vial beyond-use-date, a new dose must be prepared from a new vial.

This document is confidential.

#### 10.3 Measures to Minimize Bias: Randomization and Blinding

##### 10.3.1 Randomization

All participants will be centrally assigned to randomized study intervention using an IRT. Before the study is initiated, user guides, the log in information, and directions for the IRT will be provided to each study site. Randomization will be stratified by age and by risk (≥ 18 to < 65 years "at no risk", ≥ 18 to < 65 years "at risk", and ≥ 65 years), with at least 25% of participants to be enrolled in the older age stratum and/or in the ≥ 18 to < 65 years "at risk" stratum.

Where a participant does not meet all the eligibility criteria but incorrectly received study intervention, the investigator should inform the Study Physician immediately, and a discussion should occur between the Study Physician and the investigator regarding whether to continue or discontinue the participant.

##### 10.3.2 Blinding

Neither the participant nor any of the investigators or Sponsor staff who are involved in the treatment or clinical evaluation and monitoring of the participants will be aware of the study intervention received. Since GRAd-COV2 and placebo are visually distinct prior to dose preparation (due to differences in container closure), IMP will be handled by an unblinded pharmacist (or designee in accordance with local and institutional regulations) at the study site. Once drawn into syringes for administration, GRAd-COV2 and placebo are not visually distinct from each other.

In the phase II part of the study, all participants will receive two doses (one vaccine and one placebo in the single dose arm; two vaccines in the double dose arm; two placebo injections in the placebo arm). If the single dose arm is selected, all participants of phase III will receive one dose (vaccine or placebo); if the double-dose arm is selected for phase III, all participants will receive two doses (two vaccine doses or two placebo).

The IRT will provide the investigator(s) or pharmacists a randomization number to be associated with the actual dose tracking number (dose kit number) allocated to the participant at the dispensing visit. Routines for this will be described in the IRT user manual that will be provided to each study site.

The randomization code should not be broken except in medical emergencies when the appropriate management of the participant requires knowledge of the treatment randomization. The investigator documents and reports the action to the Sponsor, without revealing the treatment given to participant to the Sponsor staff.

The Sponsor retains the right to break the code for SAEs that are unexpected and are suspected to be causally related to an investigational medicinal product and that potentially require expedited reporting to regulatory authorities. Randomization codes will not be broken for the planned analyses of data until all decisions on the evaluability of the data from each individual participant have been made and documented.

This document is confidential.

Since the COVID-19 vaccination campaign is rapidly progressing and around 50 % of the target population has received at least one dose of the available authorized vaccines, it is considered no longer feasible and even unethical to keep subjects participating in ongoing blinded clinical trials in the placebo arm. Therefore, considering that all randomized subjects have already reached day 57 visit, which is one of the identified key study time point for immune response assessment, upon approval by both the Competent Authority (CA) and Ethics Committee, the randomization code will be automatically broken for all subjects. The participants assigned to placebo arm will be discontinued from the study as soon as they receive alternative available vaccines in the frame of national vaccination campaign. On the contrary participants belonging the two study arms who received GRAd-COV2 vaccine will continue to be monitored for safety and efficacy according to the protocol even if they will decide to access to the vaccination campaign.

##### 10.3.3 Procedures for Unblinding

The IRT will be programmed with blind-breaking instructions. In case of an emergency, in which the knowledge of the specific blinded study intervention will affect the immediate management of the participant's condition (eg, antidote available), the investigator has the sole responsibility for determining if unblinding of a participants' intervention assignment is warranted. Participant safety must always be the first consideration in making such a determination. If a participant's intervention assignment is unblinded, the Sponsor must be notified within 24 hours after breaking the blind.

#### 10.4 Study Intervention Compliance

When participants are dosed at the study site, they will receive study intervention directly from the investigator or designee, under medical supervision. The date, and time if applicable, of dose administered will be recorded in the source documents and recorded in the eCRF. The dose of study intervention and study participant identification will be confirmed at the time of dosing by a member of the study site staff other than the person administering the study intervention.

#### 10.5 Concomitant Therapy

Any medication or vaccine (including over-the-counter or prescription medicines) that the participant is receiving at the time of enrollment or receives during the period specified in the SoA (Section 3), must be recorded in the eCRF along with the information listed below.

Vitamins and/or herbal supplements are not to be recorded.

- Reason for use
- Dates of administration including start and end dates
- Dosage information including dose and frequency

The Study Physician should be contacted if there are any questions regarding concomitant or prior therapy.

This document is confidential.

##### 10.5.1 Permitted Concomitant Medications

Participants may take concomitant medications prescribed by their primary care provider for management of chronic medical conditions and/or for health maintenance. Primary care providers, or where appropriate investigators, should prescribe appropriate concomitant medications or treatments deemed necessary to provide full supportive care and comfort during the study. Participants who develop COVID-19 after receiving study intervention should be treated with licensed medications and interventions according to standard of care. All routine vaccinations other than influenza are permitted beginning > 30 days after the last dose of study intervention. Licensed influenza vaccines are permitted at any time.

##### 10.5.2 Prohibited Concomitant Medications

The following medications are prohibited, and the Sponsor must be notified if a participant receives any of these prohibited medications. The use of the following concomitant medications and/or vaccines, however, will not definitively require withdrawal of the participant from the study, but may determine a participant's evaluability in the per-protocol analysis set.

If a participant receives a prohibited concomitant medication, the investigator in consultation with the Sponsor will evaluate any potential impact on receipt of study intervention based on time the medication was administered, the medication's pharmacology and pharmacokinetics, and whether the medication will compromise the participant's safety or interpretation of the data (see Section 11.1).

- Investigational or licensed products indicated for the prevention of SARS-CoV-2 or COVID-19
- Note: For participants who become hospitalized with COVID-19, receipt of licensed treatment options and/or participation in investigational treatment studies is permitted
- Receipt of any vaccine (licensed or investigational) other than licensed influenza vaccines within 30 days prior to the first dose and after administration of last dose of study intervention
- Glucocorticoids at a dose ≥ 20 mg/day of prednisone or equivalent given daily or on alternate days for ≥ 14 consecutive days between randomization and the participant's scheduled final visit
- Other systemically administered drugs with significant immunosuppressive activity, such as azathioprine, tacrolimus, cyclosporine, methotrexate, or cytotoxic chemotherapy between randomization and the participant's scheduled final visit
- Immunoglobulins and/or any blood product

#### 10.6 Dose Modification

Study intervention will be administered as described in Section 10.1.2. Dose modification is not permitted.

#### 10.7 Intervention After the End of the Study

There is no intervention after the end of the study (see definition in Section 8.3).

This document is confidential.

### 11 DELAYING OR DISCONTINUING STUDY TREATMENT AND PARTICIPANT WITHDRAWAL FROM THE STUDY

#### 11.1 Criteria for Delay of Study Treatment

Body temperature must be measured at the Day 1 and Day 22 visits prior to any study treatment administration. The following events constitute criteria for delay of study treatment, and if either of these events occur at the time scheduled for dosing, the participant may be injected at a later date within the time window specified in Tables 2-3, or the participant may be discontinued from dosing at the discretion of the investigator:

- Acute moderate or severe infection with or without fever at the time of dosing
- Fever, defined as body temperature ≥ 37.3°C (99.14°F) at the time of dosing

Participants with a minor illness without fever, as assessed by the investigator, can be administered IMP. Participants with a fever of 37.3°C 99.14°F) or higher will be contacted within the time window acceptable for participation and reevaluated for eligibility. If the Investigator determines that the participant's health on the day of administration temporarily precludes dosing with IP, the visit should be rescheduled within the allowed interval for that visit.

#### 11.2 Discontinuation of Study Treatment

Every reasonable attempt will be made to follow up with participants for safety throughout the entire study period, even if further dosing is discontinued or the participant misses one or more visits. Unless consent is withdrawn, a participant who withdraws or is withheld from receiving the second dose of study vaccine will remain in the study and complete all scheduled visits and assessments.

The investigator, in consultation with the Sponsor's medical monitor, may withhold a participant from further dosing if the participant experiences any of the following:

- Becomes pregnant Develops, during the course of the study, symptoms or conditions listed in the exclusion criteria
- Experiences an AE (other than reactogenicity) after dosing that is considered by the investigator to be related to IMP and is of Grade 3 (severe) or greater intensity
- Experiences an AE or SAE that, in the judgment of the investigator, requires study IMP withdrawal due to its nature, severity, or required treatment, regardless of the causal relationship to vaccine
- Experiences a clinically abnormal vital sign measurement or finding on physical examination, or general condition that, in the judgment of the investigator, requires IMP withdrawal. The reason(s) for discontinuation from further dosing will be recorded in the eCRF.

This document is confidential.

Prior to receiving a second dose of IMP, participants will be reassessed to ensure that they continue to meet eligibility requirements as outlined below.

The following events in a participant constitute absolute contraindications to any further dosing of the IMP to that participant. If any of these events occur during the study, the participant must not receive additional doses of vaccine but will be encouraged to continue study participation for safety through 24 months following last dose.

- Diagnosed COVID-19 by detection of SARS-CoV-2 in Day 1 NP swab or COVID-19 diagnosed prior to Day 22. If COVID-19 is suspected on or prior to Day 22, further administration of IMP must be withheld until COVID-19 test results are available.
- Anaphylaxis or systemic hypersensitivity reaction following the administration of vaccine.
- Any SAE judged by investigator or Sponsor to be related to study vaccine.
- Any clinically significant medical condition that, in the opinion of the investigator, poses an additional risk to the participant if he/she continues to participate in the study.

The following events constitute contraindications to administration of study vaccine at certain points in time, and if any of these events occur at the time scheduled for dosing, the participant may be injected at a later date, within the time window specified in the SoA, or the participant may be withdrawn from dosing at the discretion of the investigator (Section 11.3):

- Acute moderate or severe infection with or without fever at the time of dosing
- Fever, defined as body temperature ≥ 37.3°C (99.14°F) at the time of dosing

Participants with a minor illness without fever, as assessed by the investigator, can be administered IMP. Participants with a fever of 37.3°C (99.14°F) or higher will be contacted within the time window acceptable for participation and reevaluated for eligibility.

#### 11.3 Participant Withdrawal from the Study

Participants who withdraw from the study will not be replaced. A “withdrawal” from the study refers to a situation wherein a participant does not return for the following and final visits planned in the protocol.

Participants can withdraw consent and withdraw from the study at any time, for any reason, without prejudice to further treatment the participant may need to receive. The investigator will request that the study participant complete all study procedures pending at the time of withdrawal.

If participant desires to withdraw from the study because of an AE, the investigator will try to obtain agreement to follow up with the participant until the event is considered resolved or stable and will then complete the end of study electronic case report form (eCRF).

Information related to the withdrawal will be documented in the eCRF. The Investigator will document whether the decision to withdraw a participant from the study was made by the participant or by the Investigator, as well as which of the following possible reasons was responsible for withdrawal:

This document is confidential.

- AE (specify)
- SAE (specify)
- Death
- Pregnancy
- Study terminated by Sponsor
- Withdrawal of consent by participant (specify)
- Other (specify)

Participants who are withdrawn from the study because of AEs (including SAEs) must be clearly distinguished from participants who are withdrawn for other reasons. Investigators will follow up with participants who are withdrawn from the study as result of an SAE or AE until resolution of the event.

If during the conduct of the study the participant should find himself in a position to access the national COVID vaccination program, the investigator should provide the participant with the information related to the immunological values and discuss with him the best therapeutic option to choose. However, it would be desirable that, even after vaccination with the commercial drug, the subject remains in the study until the end, attending all the scheduled follow-up visits

A participant withdrawing from the study may request destruction of any samples taken and not tested, and the investigator must document this in the site study records.

If the participant withdraws consent for disclosure of future information, the Sponsor may retain and continue to use any data collected before such a withdrawal of consent.

The Sponsor will continue to retain and use all research results that have already been collected for the study evaluation, unless the participant has requested destruction of these samples. All biological samples that have already been collected may be retained and analyzed at a later date (or as permitted by local regulations).

#### 11.4 Lost to Follow-up

Participants will be considered lost to follow-up (LTFU) if they repeatedly fail to return for scheduled visits without stating an intention to withdraw consent and they cannot be contacted by the study site. The following actions must be taken if a participant fails to return to the clinic for a required study visit:

- The site must attempt to contact the participant and reschedule the missed visit as soon as possible, counsel the participant on the importance of maintaining the assigned visit schedule and ascertain whether the participant wishes to and/or should continue in the study.
- Before a participant is deemed lost to follow-up, the investigator or designee must make every effort to regain contact with the participant (where possible, 3 telephone calls and, if necessary, a certified letter to the participant's last known mailing address or local equivalent methods). These contact attempts (eg, dates of telephone calls and registered

This document is confidential.

letters) should be documented in the participant's medical record. A participant should not be considered LTFU until these efforts have been made.

- Should the participant continue to be unreachable, he/she will be considered to have withdrawn from the study.

### 12 STUDY ASSESSMENTS AND PROCEDURES

- Study procedures and their timing are summarized in the SoA (Section 3). Protocol waivers or exemptions are not allowed.
- Immediate safety concerns should be discussed with the Sponsor immediately upon occurrence or awareness.
- Adherence to the study design requirements, including those specified in the SoA (Section 3) is essential and required for study conduct.
- All screening evaluations must be completed and reviewed to confirm that potential participants meet all eligibility criteria. The investigator will maintain a screening log to record details of all participants screened and to confirm eligibility or record reasons for screening failure, as applicable.
- Procedures conducted as part of the participant's routine clinical management (eg, blood count) and obtained before signing of the ICF may be utilized for screening or baseline purposes provided the procedures met the protocol-specified criteria and were performed within the time frame defined in the SoA, that is not earlier than14 days
- Due to the ongoing pandemic, recent regulatory and local Ethics Committee and public health guidance will be applied at the site locations regarding alternations in the ability of study participants to attend an investigational site for protocol-specified visits, with the site's investigator being allowed to conduct safety assessments (eg, telephone contact, alternative location for assessment, including local laboratories or imaging centres) when necessary and feasible, as long as such visits are sufficient to assure the safety of study participants. Serum samples may be drawn using local phlebotomy services, home health, or other modalities if site visits cannot occur. Study vaccination visits must have adequate oversight for issues associated with immediate severe reactions.

#### 12.1 Efficacy Assessments

##### 12.1.1 Monitoring COVID-19 Symptoms

Study sites will contact participants weekly (telephone/email/text message) through Day 360 with reminders to monitor for COVID-19 symptoms reporting. Participants who present with at least one of the COVID-19 qualifying symptoms listed below must contact the study team. With the exception of fever, shortness of breath, or difficulty breathing, the symptom must be present for 2 or more days. During the 7 days following administration of dose of study intervention, investigator judgement should be used to determine which participants should initiate illness visits as symptoms may be due to the reactogenicity of the study intervention as opposed to potentially due to infection with SARS-CoV-2.

If a participant presents with a COVID-19 qualifying symptom(s) (Table 7) will be tested locally for SARS-CoV-2 (see Section 12.6.1.1). If positive, the participant will be instructed to continue illness visits. If negative, the participant will continue with scheduled assessments per his/her assigned study (ie, phase II [Table 2] or phase III [Table 3]).

*Table 7 COVID-19 Qualifying Symptoms*

| **Participant must present with at least one of the following symptoms** | |
|---|---|
| **Duration** | **Symptom** |
| No minimum duration | Fever |
| | Shortness of breath |
| | Difficulty breathing |
| Must be present for ≥ 2 days | Chills |
| | Cough |
| | Fatigue |
| | Muscle aches |
| | Body aches |
| | Headache |
| | New loss of taste |
| | New loss of smell |
| | Sore throat |
| | Congestion |
| | Runny nose |
| | Nausea |
| | Vomiting |
| | Diarrhea |

Adapted from (CDC 2020).

##### 12.1.2 Illness Visits

Symptomatic participants (as defined in Section 12.1.1) will be instructed to visit the study site for initiation of illness assessments (Table 4); where supported, home or remote visits may be substituted for the on-site visits. Symptomatic participants will complete the Day 1 illness visit and will be instructed to continue with the home collection requirements (e.g. digital medical device and Illness e-Diary recordings). SARS-CoV-2 RT-PCR results will be available during the home collection period and participants will be informed of their status. The results of the COVID-19 RT-PCR testing should also be reported to the participants' primary care providers. Only participants who test positive will be instructed to continue with the illness visits, including digital medical device and Illness e-Diary recordings. Participants who test negative for SARS-CoV-2 will be instructed to stop all illness visit assessments and return the digital medical device. Participants will continue with follow-up visits

###### 12.1.2.1 SARS-COV-2 Testing and Other Virology Assessments

At the Day 1 illness visit, nasopharyngeal swabs will be collected and tested for SARS-COV-2 by authorized RT-PCR assays (see Section 12.6.1.1).

Other virology assessments are described in Section 12.6.

###### 12.1.2.2 Digital Medical Device

At the Day 1 illness visit, participants will be provided with individual certified digital medical device for measuring, at least daily, blood pressure, heart rate, body temperature and transdermal SpO2. Patients will be also provided with an electronic patient diary in which to report any symptoms, AEs (if any) and the concomitant medications (if any).

The data self-recorded by the participants through the medical devices will be automatically transmitted via an encrypted transaction to a telemedicine platform (Genius ROSA) fully validated, certified Class II Medical Device and GDPR compliant, for a real time availability to the medical staff of the clinical sites who can then provide appropriate follow-up and manage the patient according to the official national or local guidance for the home care of Covid-19 positive patients.

The data from the medical devices and remote monitoring are not intended to substitute for protocol-mandated standard safety monitoring, participant self-reporting, or participant oversight, which remains the responsibility of the investigator.

Along with the device, participants will be provided with a paper-based Quick Start Guide containing general instructions for the proper use of the medical devices. A reference copy of the document will be retained in the Site Master File.

Participants who test positive for SARS-CoV-2 will be instructed to continue the self-measurements with the medical devices until the COVID-19-associated symptoms resolve or until the Day 28 illness visit. Participants who test negative and stop the illness visits will be instructed to return the digital medical device kit set.

###### 12.1.2.3 Illness e-Diary

An Illness e-Diary will be used to collect self-reported information about COVID-19- associated symptoms (listed below per CDC (CDC 2020). At the Day 1 illness visit, participants (or, if applicable, their caregiver, surrogate, or legally authorized representative) will be given access to the Illness e-Diary and trained by study staff on how to record the information and assess the severity of the symptoms.

This document is confidential.

Participants who test positive for SARS-CoV-2 will be instructed to continue recording in the Illness e-Diary until symptoms resolve or until the Day 28 illness visit. Participants who test negative will be instructed to stop Illness e-Diary recording.

Study sites will monitor the health status of participants via Illness e-Diary responses after the Day 1 illness visit, and will call participants as needed based on these responses.

**COVID-19 Symptoms**

- Fever
- Shortness of breath
- Difficulty breathing
- Chills
- Cough
- Fatigue
- Muscle aches
- Body aches
- Headache
- New loss of taste
- New loss of smell
- Sore throat
- Congestion
- Runny nose
- Nausea
- Vomiting
- Diarrhea

##### 12.1.3 Severe Covid-19

The severity of COVID-19 will be evaluated in participants who test positive for SARS-CoV-2 by RT-PCR. A diagnosis of severe or critical COVID-19 will include laboratory-confirmed COVID-19 (SARS-CoV-2 RT-PCR-positive symptomatic illness) plus any of the following:

- Clinical signs at rest indicative of severe systemic illness (respiratory rate ≥ 30 breaths per minute, heart rate ≥ 125 beats per minute, oxygen saturation ≤ 93% on room air at sea level, or partial pressure of oxygen to fraction of inspired oxygen ratio < 300 mm Hg)
- Respiratory failure (defined as needing high-flow oxygen, noninvasive ventilation, mechanical ventilation or extracorporeal membrane oxygenation)
- Evidence of shock (systolic blood pressure < 90 mm Hg, diastolic blood pressure
- < 60 mm Hg, or requiring vasopressors)
- Significant acute renal, hepatic, or neurologic dysfunction
- Admission to an intensive care unit
- Death

This document is confidential.

###### 12.1.4 Medical Notes Review

With the participant's consent, the study team will request access to medical notes or submit a data collection form for completion by attending clinical staff on any medically-attended COVID-19 episodes. Any data relevant for assessment of the efficacy endpoints or vaccine-associated enhanced respiratory disease (see Section 12.3.9) will be collected. These are likely to include, but not limited to, information on intensive care unit admissions, clinical parameters such as oxygen saturation, respiratory rates and vital signs, need for oxygen therapy, need for ventilatory support, imaging, blood tests results, and overall outcome (survival or death).

###### 12.1.5 Monitoring of Asymptomatic Infection

Blood samples will be collected according to the SoA (Section 3) for SARS-CoV-2 serology testing to monitor participants for asymptomatic infection. See description of assessment in Section 12.5.2.1.

##### 12.2 Safety Assessments

Planned time points for all safety assessments are provided in the SoA (Section 3).

###### 12.2.1 Physical Examinations

A complete physical examination will be performed at screening followed by targeted physical examinations as specified in the SoA (Section 3).

- A complete physical examination will include, but not be limited to, assessment of height, weight, general appearance, head, ears, eyes, nose, throat, neck, skin, as well as cardiovascular, respiratory, abdominal, and nervous systems. Each clinically significant abnormal finding at screening will be recorded in the medical history.
- A targeted physical examination will include areas suggested by the medical history. Each clinically significant abnormal finding following vaccination will be recorded as an AE.
- All physical examinations will be performed by a licensed healthcare provider (eg, physician, physician assistant, or licensed nurse practitioner).

###### 12.2.2 Vital Signs

Vital signs, including heart rate, pulse oximetry, blood pressure, and body temperature, will be performed as specified in the SoA (Section3). The participant should be resting prior to the collection of vital signs.

Data collected through the digital medical device on blood pressure, heart rate, body temperature, and oxygen saturation level will be recorded as exploratory efficacy measurements and should not be reported as AEs unless resulting in an MAAE or SAE.

Situations in which vital sign results should be reported as AEs are described in Section 12.3.5.

This document is confidential.

##### 12.2.3 Clinical Laboratory Assessments

A Laboratory Manual will be provided to the sites that specifies the procedures for collection, processing, storage, and shipment of samples, as well as laboratory contact information, specific to this clinical study.

For women participants of childbearing potential, a urine sample for pregnancy testing will be collected according to the SoA (Section.3). Urine pregnancy tests for β-hCG may be performed at the site using a licensed test (dipstick). If urine tests positive or indeterminate, a quantitative serum β-hCG will be performed for confirmation.

Only for phase II a safety blood sample will be collected at baseline and day 36 to test the following parameters:

Hematocrit, Hemoglobin, Red Blood Cell, White Blood Cell with formula, MCV, Platelets, Creatinine, BUN, AST, ALT, Total Bilirubin, Direct Bilirubin, Glucose.

Additional safety samples may be collected if clinically indicated at the discretion of the investigator.

#### 12.3 Adverse Events and Serious Adverse Events

The principal investigator is responsible for ensuring that all staff involved in the study are familiar with the content of this section.

The definitions of an AE or SAE can be found in Appendix 1.

All AEs are considered to be unsolicited AEs (collected by 'open question' at study visits) unless categorized as solicited AEs recorded in the phase II only.

Solicited AEs are local or systemic predefined events for assessment of reactogenicity. Solicited AEs will be collected in a Solicited AE e-Diary only for participants in the phase II part of the study (see Section 12.3.7), and will be assessed separately from the (unsolicited) AEs collected during the study.

General information for AEs in this protocol excludes solicited AEs.

AEs will be reported by the participant (or, when appropriate, by a caregiver, surrogate, or the participant's legally authorized representative).

The investigator and any designees are responsible for detecting, documenting, and recording events that meet the definition of an AE.

##### 12.3.1 Time Period and Frequency for Collecting AE and SAE Information

AEs will be recorded for 28 days post each dose of study intervention.

Solicited AEs will be recorded only for participants in the phase II part of the Study for 7 days post each dose of study intervention.

SAEs will be recorded from the time of signature of the informed consent form through the last participant contact.

MAAEs and AESIs will be recorded from Day 1, post treatment, through the last participant contact.

See the SoA (Section 3) for the scheduled timepoints.

If the investigator becomes aware of an SAE with a suspected causal relationship to the study intervention that occurs after the end of the clinical study in a participant treated by him or her, the investigator shall, without undue delay, report the SAE to the Sponsor.

##### 12.3.2 Follow-up of AEs and SAEs

Any AEs that are unresolved at the participant's last AE assessment in the study are followed up by the investigator for as long as medically indicated, but without further recording in the eCRF. The Sponsor retains the right to request additional information for any participant with ongoing AE(s)/SAE(s) at the end of the study, if judged necessary.

**AE Variables**

The following variables will be collected for each AE:

- AE (verbatim)
- Date when the AE started and stopped
- Severity grade/maximum severity grade/changes in severity grade
- Whether the AE is serious or not
- Investigator causality rating against the study intervention (yes or no)
- Action taken with regard to study intervention
- AE caused participant's withdrawal from study (yes or no)
- Outcome

In addition, the following variables will be collected for SAEs:

- Date AE met criteria for SAE
- Date investigator became aware of SAE
- AE is serious due to
- Concomitant medication
- Therapy administered, if any
- Date of hospitalization
- Date of discharge
- Probable cause of death
- Date of death
- Autopsy performed
- Causality assessment in relation to study procedure(s)

- Causality assessment to other medication

The grading scales from US FDA guidance for healthy volunteers enrolled in a preventive vaccine clinical study [20] will be utilized for all unsolicited events with an assigned severity grading.

###### 12.3.3 Causality Collection

The investigator should assess causal relationship between study intervention and each AE, and answer ‘yes’ or ‘no’ to the question ‘Do you consider that there is a reasonable possibility that the event may have been caused by the investigational product?’

For SAEs, causal relationship should also be assessed for other medication and study procedures. Note that for SAEs that could be associated with any study procedure the causal relationship is implied as ‘yes.’

A guide to the interpretation of the causality question is found in Appendix 1.

###### 12.3.4 Adverse Events Based on Signs and Symptoms

All AEs spontaneously reported by the participant or reported in response to the open question from the study site staff: ‘Have you had any health problems since the previous visit/you were last asked?’, or revealed by observation will be collected and recorded in the eCRF. When collecting AEs, the recording of diagnoses is preferred (when possible) to recording a list of signs and symptoms. However, if a diagnosis is known and there are other signs or symptoms that are not generally part of the diagnosis, the diagnosis and each sign or symptom will be recorded separately.

###### 12.3.5 Adverse Events Based on Examinations and Tests

The results from the Clinical Study Protocol-mandated vital signs will be summarized in the CSR.

Deterioration as compared to baseline in protocol-mandated vital signs should therefore only be reported as AEs if they fulfil any of the SAE or MAAE criteria or are considered to be clinically relevant as judged by the investigator (which may include but not limited to consideration as to whether treatment or non-planned visits were required).

If deterioration in a vital sign is associated with clinical signs and symptoms, the sign or symptom will be reported as an SAE or MAAE, and the associated vital sign will be considered as additional information.

###### 12.3.6 Hy’s Law

Cases where a participant shows elevations in liver biochemistry may require further evaluation. Any occurrences of AST or ALT ≥ 3 × ULN together with total bilirubin ≥ 2 × ULN and confirmed as a Hy's Law case should be reported as an SAE.

This document is confidential.

**Hy's Law**

AST or ALT ≥ 3 × ULN together with TBL ≥ 2 × ULN, where no other reason, other than the study intervention, can be found to explain the combination of increases, eg, elevated ALP indicating cholestasis, viral hepatitis, another drug. The elevation in transaminases must precede or be coincident with (ie, on the same day) the elevation in TBL, but there is no specified timeframe within which the elevations in transaminases and TBL must occur.

##### 12.3.7 Solicited Adverse Events (Only for Phase II Part of the Study)

Local and systemic predefined solicited AEs for reactogenicity assessment (Table 8) will be collected in a Solicited AE e-Diary for 7 days following administration of each dose of study intervention only from participants in the phase II part of the Study.

Solicited AEs should not be reported as unsolicited AEs (see Section 12.3). However, solicited AEs should be reported as SAEs or MAAEs if they fulfil the criteria (see Sections 12.3 and 12.3.8, respectively).

*Table 8 List of Predefined Solicited Adverse Events for Reactogenicity Assessment*

| Local | Systemic |
|---|---|
| Pain at the site of injection | Fever (> 100 °F [> 37.8 °C]) [a] |
| Erythema/redness at the site of injection [b] | Chills |
| Tenderness | Muscle pains |
| Induration/swelling at the site of the injection [b] | Fatigue |
| | Headache |
| | Malaise |
| | Nausea |
| | Vomiting |

[a] Fever measured by any route. Investigators who consider a temperature lower than this cutoff as a fever or a 'fever' reported by participants without documentation by a thermometer should record the event as 'elevated body temperature.'

[b] Swelling and redness must be ≥ 0.25 inches (≥ 0.6 centimeters) in diameter.

**Solicited AE e-Diary**

On Day 1, participants in the phase II part of the study (or, if applicable, their caregiver, surrogate, or legally authorized representative) will be given an oral thermometer, tape measure, and access to the Solicited AE e-Diary, with instructions on use, along with the emergency 24-hour telephone number to contact the on-call study physician if needed.

This document is confidential.

Participants will be instructed to record for 7 days following administration of each dose of study intervention, the timing and severity of local and systemic solicited AEs, if applicable, and whether medication was taken to relieve the symptoms.

**Severity Assessment of Solicited AEs**

Severity will be assessed for solicited AEs by the participant (or, if applicable, their caregiver, surrogate, or legally authorized representative) according to toxicity grading scales modified and abridged from the US FDA guidance [20] as defined in Appendix 4. Because solicited AEs are expected to occur after vaccination, they will not be assessed for relationship to study intervention.

##### 12.3.8 Medically Attended Adverse Events

MAAEs will be collected according to the timepoints specified in the SoA (Section 3).

MAAEs are defined as AEs leading to medically-attended visits that were not routine visits for physical examination or vaccination, such as an emergency room visit, or an otherwise unscheduled visit to or from medical personnel (medical doctor) for any reason. AEs, including abnormal vital signs, identified on a routine study visit or during the scheduled illness visits will not be considered MAAEs.

##### 12.3.9 Adverse Events of Special Interest

AESIs will be collected according to the timepoints specified in the SoA (Section 3).

AESIs are events of scientific and medical interest specific to the further understanding of study intervention safety profile and require close monitoring and rapid communication by the investigators to the Sponsor. An AESI can be serious or non-serious. All AESIs will be recorded in the eCRF. Serious AESIs will be recorded and reported as per Section 12.3.10.

AESIs for GRAd-COV2 are listed below. They are based on Brighton Collaboration case definitions [21].

**Brighton Collaboration AESIs**

<u>AESIs relevant to vaccination in general include:</u>

- Neurologic
  - Generalized convulsion
  - Guillain-Barre syndrome
  - Acute disseminated encephalomyelitis
- Immunologic
  - Vasculitis
  - Anaphylaxis
  - Vaccine-associated enhanced respiratory disease
- Hematologic
  - Thrombocytopenia

This document is confidential.

##### 12.3.10 Reporting of Serious Adverse Events

All SAEs have to be reported, whether or not considered causally related to the study intervention, or to the study procedure(s). All SAEs will be recorded in the eCRF.

If any SAE occurs in the course of the study, investigators or other site personnel will inform the appropriate Sponsor representatives and the Contract Research Organization, recording the SAE data on the AEs section of the eCRF within 24 hours of learning of the event. The paper SAE forms are only intended as a back-up option when the eCRF system is not accessible. In this case, the original paper forms are to remain on site and copies are to be transmitted via email or confirmed facsimile (fax) transmission to:

- Fax number: +39 02 36026913

The investigator should also report information on SAEs that continue after patient has completed his/her participation in the study (whether study completion or withdrawal), unless patient has withdrawn his/her consent.

Information on SAEs will be recorded on the AEs section form in the eCRF. Follow-up reports (as many as required) should be completed following the same procedure; the system will mark the SAE form as "Follow up no. XX"

The designated Sponsor representative will work with the investigator to ensure that all the necessary information is provided to ReiThera within one calendar day of initial receipt for fatal and life-threatening events and within 5 calendar days of initial receipt for all other SAEs.

For fatal or life-threatening AEs where important or relevant information is missing, active follow-up will be undertaken immediately. Investigators or other site personnel will inform Sponsor representatives of any follow-up information on a previously reported SAE within one calendar day, ie, immediately but no later than 24 hours of when he or she becomes aware of it.

Once the investigators or other site personnel indicate an AE is serious in the EDC system, an automated email alert is sent to the designated Sponsor representative.

The Sponsor representative will advise the investigator/study site staff how to proceed. For further guidance on the definition of an SAE, see Appendix 1.1.

The reference document for definition of expectedness is the GRAd-COV2 IB, Section 6.2.2.

##### 12.3.11 Pregnancy

All pregnancies and outcomes of pregnancy with conception dates following administration of study intervention should be reported to the Sponsor, except if the pregnancy is discovered before the participant has received any study intervention.

###### 12.3.11.1 Maternal Exposure

Female participants who are pregnant or have a confirmed positive pregnancy test at screening or Day 1 will be excluded from the study (see Section 9.2). Female participants who are pregnant or have a confirmed positive pregnancy test before the second dose injection, will not receive that dose. Pregnancy itself is not regarded as an AE unless there is a suspicion that the study intervention may have interfered with the effectiveness of a contraceptive medication. Congenital abnormalities/birth defects and spontaneous miscarriages should be reported and handled as SAEs. Elective abortions without complications should not be handled as AEs. The outcome of all pregnancies (spontaneous miscarriage, elective termination, ectopic pregnancy, normal birth or congenital abnormality) should be followed up and documented even if the participant was discontinued from the study.

If any pregnancy occurs in the course of the study, then the investigator or other site personnel informs the appropriate Sponsor representatives within 1 day, ie, immediately but no later than 24 hours of when he or she becomes aware of it.

The designated Sponsor representative works with the investigator to ensure that all relevant information is provided to ReiThera within 1 or 5 calendar days for SAEs (see Section 12.3.10) and within 30 days for all other pregnancies that are not associated with an SAEs.

The same timelines apply when outcome information is available.

If the investigator becomes aware of a pregnancy occurring in the partner of a subject participating in the study, the pregnancy should be reported to the Sponsor (or designee) within 24 hours of knowledge of the event. Information regarding the pregnancy must only be submitted after obtaining written consent from the pregnant partner. The investigator will arrange counseling for the pregnant partner by a specialist to discuss the risks of continuing with the pregnancy and the possible effects on the fetus.

##### 12.3.12 Medication Error

If a medication error occurs, then the investigator or other site personnel informs the appropriate Sponsor representatives within 1 day, ie, immediately but no later than 24 hours of when he or she becomes aware of it.

The designated Sponsor representative works with the investigator to ensure that all relevant information is completed within 1 (Initial Fatal/Life-Threatening or follow up

Fatal/Life-Threatening) or 5 (other serious initial and follow up) calendar days if there is an SAE associated with the medication error (see Section 12.3.10) and within 30 days for all other medication errors.

This document is confidential.

The definition of a Medication Error can be found in Appendix 1.4 4.

###### 12.3.13 Medical Device Deficiencies

Any deficiency observed with the digital medical device will be collected and reported to the Contract Research Organization by the investigators or other site personnel within one day ie, immediately but no later than 24 hours of when he or she becomes aware of it.

A medical device deficiency is an inadequacy of a medical device with respect to its identity, quality, durability, reliability, safety, or performance. Medical device deficiencies include malfunctions, use errors, and information supplied by the manufacturer.

The Contract Research Organization will then replace the defective medical device.

##### 12.4 Overdose

For this study, any dose of study intervention exceeding that specified in the protocol will be considered an overdose.

There is no specific treatment for an overdose with GRAd-COV2. If overdose occurs, the participant should be treated supportively with appropriate monitoring as necessary.

- An overdose with associated AEs is recorded as the AE diagnosis/symptoms on the relevant AE modules in the eCRF and on the Overdose eCRF module
- An overdose without associated symptoms is only reported on the Overdose eCRF module

If an overdose occurs in the course of the study, the investigator or other site personnel inform appropriate Sponsor representatives immediately, but no later than 24 hours after when he or she becomes aware of it.

The designated Sponsor representative works with the investigator to ensure that all relevant information is provided to the ReiThera within one or 5 calendar days for overdoses associated with an SAE (see Section 12.3.10) and within 30 days for all other overdoses.

##### 12.5 Human Biological Samples

Instructions for the collection and handling of biological samples will be provided in the study-specific laboratory manual. Samples should be stored in a secure storage space with adequate measures to protect confidentiality. Further details on Handling of Human Biological Samples are provided in Appendix 2.
Samples will be stored for a maximum of 15 years from the date of the issue of the CSR in line with consent and local requirements, after which they will be destroyed/repatriated.

Remaining biological sample aliquots will be retained at the Sponsor or its designee for a maximum of 15 years following issue of the CSR. Additional use excludes genetic analysis and includes but is not limited to, analysis of COVID-19 and other coronavirus-related diseases or vaccine-related responses, e.g.

exploratory immunology, such as serology, systems serology and profiling of B- and T-cell repertoire. The results from further analysis will not be reported in the CSR.

##### 12.5.1 Pharmacokinetics

Pharmacokinetic parameters are not evaluated in this study.

##### 12.5.2 Immunogenicity Assessments

###### 12.5.2.1 SARS-CoV-2 Serology Assessments

Serum samples will be collected to assess SARS-CoV-2 antigen-specific antibody levels from participants according to the SoA (Section3). Authorized laboratories will assess serologic responses to GRAd-COV2 by the rate of participants seroconverting from negative to positive as defined by a validated immunoassay directed at the SARS-CoV-2 S antigen. The rate of asymptomatic SARS-CoV-2 infection in participants receiving GRAd-COV2 vs placebo will be determined by seroconversion in a SARS-CoV-2 Nucleocapsid assay operated by an authorized laboratory.

###### 12.5.2.2 SARS-CoV-2 Neutralizing Antibody Assessments

Serum samples to measure SARS-CoV-2 neutralizing antibody levels will be collected from participants in the phase II part of the study according to the timepoints specified in the SoA (Section3). Authorized laboratories will measure neutralizing antibodies to SARS-CoV-2 using validated wild-type neutralization assay or pseudo-neutralization assays.

###### 12.5.2.3 Assessment of Cell-mediated Immune Responses (Exploratory)

Cell-mediated immune responses (i.e. B-cell and T-cell responses) will be assessed by characterizing PBMCs using methods that may include T-cell ELISpot assays to

SARS-CoV-2 antigens, flow cytometry after intracellular cytokine staining, and other methodology as determined by the Sponsor and/or authorized laboratories. Data on Th1/Th2 polarization after GRAd-COV2 vaccination will be provided and may be reported separately from the CSR. Samples will be collected from participants recruited in one site in Italy (Centro Ricerche Cliniche Verona).

###### 12.5.2.4 Additional Serum Immunogenicity

Additional serum samples for exploratory immunogenicity evaluation will be obtained according to the SoA (Section 3). Exploratory sera samples may be utilized to investigate additional humoral and cellular immune response biomarkers as well as potential correlates of protection as determined by the Sponsor and/or authorized laboratories based upon emerging safety, efficacy, and immunogenicity data.

##### 12.5.3 Pharmacodynamics

Pharmacodynamics are not evaluated in this study.

This document is confidential.

#### 12.6 Human Biological Sample Biomarkers

##### 12.6.1 Collection of Mandatory Samples for Biomarker Analysis

By consenting to participate in the study, the participant consents to participate in the mandatory research components of the study. Samples for biomarker research are required and will be collected from participants during illness visits as specified in the SoA (Section3). Nasopharyngeal swabs will be collected at site illness visits for virologic assessments. These biomarker measurements will support our understanding of potential correlates of protection, duration of immune responses, and correlations between immunogenicity and reactogenicity. Details for sample collection, processing, and testing will be provided in the Laboratory Manual.
Any results from such analyses may be reported separately from the CSR.

###### 12.6.1.1 Virologic Assessments

Instructions for obtaining and processing nasopharyngeal swab samples are provided in the Laboratory Manual. Nasopharyngeal swabs will be assessed by authorized RT-PCR assays for the detection of SARS-CoV-2 by local and central laboratories.

###### 12.6.1.2 Others Study-related Biomarker Research

Already collected samples may be analyzed for different biomarkers thought to play a role in COVID-19 severity or outcomes including, but not limited to serum or plasma cytokines, quantification of RNA, micro-RNA, and/or non-coding RNA using quantitative RT-PCR, microarray, sequencing, or other technology in blood and PBMCs to evaluate their association with observed clinical responses to GRAd-COV2. Other study-related biomarker research excludes genetic analysis.

For storage, re-use, and destruction of biomarker samples see Section 12.5.

#### 12.7 Medical Resource Utilization and Health Economics

Medical resource utilization and health economics are not applicable in this study.

This document is confidential.

### 13 STATISTICAL CONSIDERATIONS

#### 13.1 General Considerations

This Section summarizes the planned statistical analysis strategy and the procedures for the study. All the details will be provided in the statistical analysis plan (SAP), which will be finalized before the first clinical database lock for the study (ie, before the first analysis in Phase II part of the study). If, after the study has begun, but prior to any unblinding, changes are made to the primary and/or the key secondary hypotheses (or to the statistical methods related to these hypotheses), then the protocol will be amended, while changes to other secondary or exploratory analyses will not require a protocol amendment. The latter changes will be described in the SAP and the Clinical Study Report (CSR) for the study, along with an explanation as to when and why they were made.

Depending on the scenario which will be considered feasible for the phase III part of the study, a protocol amendment may be needed to re-define study design, sample size and related statistical analysis.

The data of the phase II part of the study will be analyzed at different milestones during the study: at the phase II interim analysis, which will be conducted when five-week post first dose immunogenicity and safety data are available for the first 450 participants and one-week safety data are available for all 900 participants of this study phase. Phase II data will also be analyzed on later timepoints at any phase III milestones for the statistical analysis (see below). The data of the phase II interim analysis will be assessed by the DSMB and the Steering Committee to decide whether the study can be continued or not and the best regimen to be used.

The data of the phase III part of the study will also be analyzed at different milestones: at the phase III interim analysis for efficacy, at the primary analysis for efficacy and at the final analysis. The data of the phase III efficacy interim analysis will be assessed by the DSMB and the Steering Committee to provide recommendations on actions to be taken for regulatory submission. However, even in case of successful results at this interim analysis, the study will continue until its natural end.

The type I error of the study has been adjusted only for the multiple arm comparisons in the first analysis of phase II and for the phase III efficacy interim and final analyses. The sample size of phase III will include the sample of phase II.

The case definition of the primary efficacy endpoint might change a bit as the scientific community learns more about the disease and outcomes from the different COVID-19 vaccine development programs. In this case an amendment to the study protocol will be done.

More details are provided in Sections 13.3 (on sample size), 13.6 (on multiplicity adjustment), and 13.9 (on interim analyses).

#### 13.2 Statistical Hypotheses for the Primary Efficacy Analysis

The primary efficacy endpoint for phase III is a binary response (case yes / case no), whereby a participant is defined as a COVID-19 case if their first case of SARS-CoV-2 RT-PCR-positive symptomatic illness occurs

This document is confidential.

≥ 28 days post single dose or ≥ 7 days post second dose of study intervention. Otherwise, a participant is not defined as a COVID-19 case.

For the primary efficacy objective, the null hypothesis of this study is that the vaccine efficacy (VE) of GRAd-COV2 to prevent first occurrence of COVID-19 is equal to 30% (ie, H0 efficacy: VE = 0.3 vs Ha efficacy: VE≠30%). The study will have met the primary efficacy objective if the observed VE point estimate is of at least 50% and the corresponding overall (ie, considering both interim and final analysis) 95% two-sided confidence interval (CI) of VE rules out 30% at either the interim analysis or the primary analysis (see below actual level of CIs at interim and primary analyses). Actually, this is equivalent to consider H0 efficacy: VE ≤ 0.3 vs Ha efficacy: VE>30% and to use the overall 97.5% one-sided CI of VE.

Since VE, i.e. the percent reduction in the hazard of the primary endpoint (GRAd-COV2 vs. placebo), is equal to 1-Hazard Ratio (HR), where HR is the ratio between the hazard of the infection in the vaccine group relative to the hazard of the infection in the control group, the null hypothesis may be equivalently expressed as: H0 efficacy: HR=0.7 vs Ha efficacy: HR≠0.7 using two-sided overall 95% CIs or H0 efficacy: HR ≥ 0.7 vs Ha efficacy: HR<0.7 using overall 97.5% one-sided CIs.

A Cox proportional hazard model will be used to assess the magnitude of the treatment group difference (ie, HR) between GRAd-COV2 and placebo at a two-sided overall 0.05 significance level.

#### 13.3 Sample Size Determination

##### 13.3.1 Phase II Part of the Study

The 450 participants (150 in each arm), who are planned for the interim analysis of the phase II part of the study, will allow to determine a 1.5-fold increase in the geometric mean ratio (GMR) of the specific antibody titers (see below) between each of the vaccine groups and the placebo group under the following assumptions:

- Antibody titers follow a log-normal distribution (natural log transformation)
- Alpha=0.025 (two-sided)
- Beta a bit lower than 0.1 (ie, power a bit higher than 90%)
- Coefficient of variation of measurements in the original scale=1.25
- Student t-test is conducted at two-sided level
- Drop-out rate is negligible.

The nominal type I error level of 0.025 (two-sided) has been based on the Bonferroni procedure for the two comparisons vs. placebo. For the comparison between the two vaccine groups, in case both separate from placebo, with a significance level of 0.05, the study will have a power of 95% to detect the same fold increase (ie, 1.5). This procedure ensures control of the interim-wise alpha level at the 0.05 level for the close testing principle.

It is to be noticed that dose selection will not be based only on immunogenicity outcome. The testing strategy described above will constitute a tool for decision making but safety evaluations as well as

This document is confidential.

logistical considerations (impact of using one vs two doses on public health) will inform the final decisions on dose selection.

The analysis of immunogenicity data collected on the whole 900 subjects enrolled in the phase II part of the study will be primarily descriptive and, therefore, no alpha adjustment for the interim and final analyses of the phase II part of the study is needed.

The assumption for the coefficient of variation estimate (ie, 1.2) seems conservative based on preliminary results of the phase I study (only young subjects).

##### 13.3.2 Phase III Part of the Study

###### 13.3.2.1 Primary Analysis (Scenarios 1 and 2)

Same assumptions can be made for sample size computation in the first two scenarios described in Section 8.1, being the evaluation of low-risk subjects of scenario 2 referred to the infection outcome and not the likelihood of getting an infection.

The primary statistical analysis will be conducted in the Efficacy analysis set (see Section 13.4).

The sample size is driven by the total number of cases to demonstrate VE to prevent COVID-19 infection of any severity. Assuming a true VE of 70%, with a 1:1 (GRAd-COV2: placebo) allocation ratio in the randomization, a total of 74 confirmed COVID-19 cases will provide 90% power to conclude that true VE>30% (i.e. to reject the null hypothesis H0: VE ≤ 30%), with one interim analysis performed at 50% of the target total number of cases, using an O'Brien-Fleming boundary for efficacy and an overall two-sided false positive error rate of 0.05. The computation method is based on a superiority by a margin test (see Section 13.2) for the ratio of two proportions. The total number of cases pertains to the Efficacy analysis set (see Section 13.4).

Approximately 10000 participants will be needed to obtained this number of cases, considering a 1:1 probability ratio to receive a single or a double IM dose of GRAd-COV2 (the active group, n = approximately 5000) or saline placebo (the control group, n = approximately 5000) with the following assumptions (power 90% and overall alpha 0.05 two-sided):

- Accrual of 74 confirmed endpoint cases is possible within 6 months
- Six-month COVID-19 incidence rate in the placebo arm is 1.25
- Six-month dropout rate is 1%
- One interim analysis at 50% of total target cases is performed with O'Brien-Fleming boundaries for efficacy monitoring
- Approximately 10% of participants are excluded from the Efficacy analysis set.

The number indicated above includes 600 participants enrolled in the phase II part of the study (considering the selected vaccine group and the placebo groups), so 9400 new participants (4700 in the active and 4700 in the control group) will have to be enrolled in the phase III part of the study.

This document is confidential.

The interim analysis will be performed when approximately 50% of the total target COVID-19 cases (ie, approximately 37 cases) meeting the primary efficacy endpoint definition are reported across the active and control groups within the population of participants who are not seropositive at baseline (Efficacy analysis set). The boundaries for interim and final analyses are described in Section 13.9, Table 14. With this number of cases, the study will have adequate power to detect VE, only if the latter is higher than expected (ie, at least 90%).

Considering an accrual period of 2 months and that the cases of the first month after vaccination are not valid cases, it is estimated that it will take approximately 5 and 8 months from study start (first subject first dose), to accrue 50% and 100% of the target number of cases (ie, approximately 37 and 74 cases, respectively) in the Efficacy analysis set.

A statistically significant finding at the interim analysis will not be considered a reason to stop the study. It will be interpreted as early assessment of efficacy and may be used for an Emergency Use approval (see Section 13.9 on interim analysis).

The assumptions for sample size computation (particularly, the incidence rate of the placebo group, the drop-out rate and the rate of exclusions from the Efficacy analysis set) will be re-evaluated at the time of starting the phase III part of the study. In addition, since this is an event driven study, the Sponsor may adjust the size of the study or the duration of the follow-up based on the blinded review of the total number of cases of COVID-19 accrued during the study and to the estimated percentages of study participants with serologic evidence of SARS-CoV-2 infection at baseline.

###### 13.3.2.2 Primary Analysis (Scenario 3)

At present, scenario 3 described under Section 8.1 is not evaluated in terms of sample size, because the assumptions for these computations are not known. If scenario 3 is to be followed due to ethic issues about placebo use, a protocol amendment will be submitted to define the sample size and the other protocol changes.

###### 13.3.2.3 Safety Endpoints

For safety outcomes, Table 9 shows the probability of observing at least 1 AE for a given true event rate of a particular AE, for various sample sizes, that approximately mimic the sample sizes that will be considered in the different milestones. For example, if the true AE rate is 1%, with 100 participants in a group, there is 77.9% probability of observing at least 1 AE.

*Table 9 Probability of Observing at Least 1 AE According to Different True Event Rates and Different Sample Sizes*

| True event rate of an AE | N=150 (Sample size per arm at phase II interim analysis for subjects observed up to 1 months) | N=300 (Sample size per arm at phase II interim analysis for subjects observed up to 7 days) | N=5000 (Sample size per arm at phase III) |
|---|---|---|---|
| **0.01%** | 0.015 | 0.030 | 0.393 |
| **0.02%** | 0.030 | 0.058 | 0.632 |
| **0.05%** | 0.072 | 0.139 | 0.918 |
| **0.10%** | 0.139 | 0.259 | 0.993 |
| **0.20%** | 0.259 | 0.452 | >0.999 |
| **0.35%** | 0.409 | 0.651 | >0.999 |
| **0.50%** | 0.529 | 0.778 | >0.999 |
| **1.00%** | 0.779 | 0.951 | >0.999 |
| **2.00%** | 0.952 | 0.997 | >0.999 |
| **5.00%** | >0.999 | >0.999 | >0.999 |
| **10.00%** | >0.999 | >0.999 | >0.999 |

AE=Adverse event; N=number of participants in the considered sample.

The table provides just a crude estimate of observing at least 1 AE for given true event rates, without considering the patient-observation time.

###### 13.3.2.4 Key Secondary Endpoint

The key secondary objective of the study is to demonstrate VE of a single IM dose of GRAd-COV2 compared to placebo for the prevention of severe or critically severe infections.

With approximately 15 cases of severe or critically severe COVID-19 infections (ie, 20% of 74 cases), the study will have approximately 80% power to show a lower limit of the 95.1% two-sided CI of VE above 0% when assuming that VE is 85%.

The sample size of this secondary endpoint may be adjusted based on a lower than expected incidence rate of COVID-19 cases during the study and on a higher than expected proportion of seropositive participants at baseline.

#### 13.4 Population for Analysis

The analysis populations considered in the present study are defined in Table 10.

*Table 10 Populations for Analysis*

| Population | Description |
|---|---|
| **All participants analysis set** | All participants screened for the study, to be used for reporting participants disposition and screening failures. |
| **Full analysis set (FAS)** | All randomized participants who received the dose of study intervention, irrespective of their protocol adherence and continued participation in the study. Participants will be analyzed according to their randomized treatment irrespective of whether or not they have prematurely discontinued, according to the intent-to-treat principle. Participants who withdraw consent to participate in the study will be included up to the date of their study withdrawal. |
| **Efficacy analysis set (EAS)** | All subjects of the FAS who had no immunologic or virologic evidence of prior COVID-19 (ie, negative nasopharingeal swab test at Day 1, and/or bAb against SARS-CoV-2 nucleocapsid below LOD or LLOQ) at Day 1 before the first dose of GRAd-COV2. Participants will be analyzed according to the group to which they were randomized. |
| **Per-protocol analysis set (PPAS)** | All subjects of the EAS who received the correct dose of randomized treatment and who do not have any major protocol deviations. Detailed criteria defining this analysis set will be documented in the SAP. |
| **Safety analysis set (SA set)** | All participants who received the dose of study intervention.<br>Erroneously treated participants (ie, those randomized to GRAd-COV2 but actually receiving placebo or vice versa) are accounted for in this analysis set by assigning them to the treatment they actually received. |
| **All available immunogenicity analysis set (AAIAS)** | All participants in the SA set who have immune response assessments (ie, 900 participants). Participants will be analyzed according to the treatment group to which they were randomized. |
| **Immunogenicity analysis set (IAS)** | All participants in the SA set who have immune response assessments and no protocol deviations judged to have the potential to interfere with the generation or interpretation of an immune response. Examples of protocol violations will be documented in the SAP. Participants will be analyzed according to the treatment group to which they were randomized. |

#### 13.5 Statistical Analyses

This Section provides a summary of the planned statistical analyses of the primary and secondary endpoints.

This document is confidential.

There is only one key secondary endpoint in the study to be tested in a confirmatory way, ie, the Time to first case of SARS-CoV-2 RT-PCR-positive severe or critical symptomatic illness. This endpoint will be tested using a hierarchical testing approach, ie, it will be tested only if the primary endpoint reaches statistical significance and using the same alpha level used for the primary endpoint. The testing procedure does not include adjustments to control the type I error rate over the other secondary efficacy endpoints.

No formal multiple comparison adjustments will be employed for multiple safety endpoints.

The interim phase II data base lock (DBL) will occur when data are available (entered and cleaned) for the first 450 participants having reached five weeks of follow-up after the first dose and for the total 900 participants having reached seven days of follow-up after the second dose. The interim efficacy DBL of the study will occur after approximately 37 cases have been confirmed for the primary endpoint within the population of participants who are not seropositive at baseline (EAS), while the primary DBL will occur after approximately 74 events have been confirmed for the primary endpoint in the same population.

All participants in the study will be assessed for efficacy and safety for 1 year following the first dose of study intervention (ie, will be observed up to Day 360). The final DBL will occur when all participants have completed the study.

All personnel involved in the conduction and the statistical analysis of the study will remain blinded until the final DBL and protocol deviations are identified (see Section 13.10 for the unblinding procedures).

All statistical analyses will be performed by the selected CRO, using SAS version 9.4. The categorical variables will be summarized using frequency and percentages, where the denominator is the underlying analysis set population unless otherwise stated. The continuous variables will be summarized with descriptive statistics of number of available observations, mean, standard deviation, median, minimum and maximum, and quartiles where appropriate.

All point estimates will be presented together with 95% two-sided CIs, unless otherwise stated. P-values corresponding to two-sided tests will be presented for comparisons between treatment groups.

##### 13.5.1 Patient Disposition

The summary of screening failures, the disposition of subjects and the exposure to vaccinations will be tabulated by treatment group and overall. This summary will include all study populations starting with the All participants set.

##### 13.5.2 Baseline Characteristics

Demographic and baseline characteristics, including medical history and physical examination, will be summarized by treatment group for the FAS, the EAS, and the IAS. If there are major differences between the FAS and the PPAS or between the AAIAS and the IAS (i.e. more than 20% difference is observed in their size), the summaries will be repeated also for the PPAS and for the AAIAS, respectively.

##### 13.5.3 Efficacy

Efficacy analyses will be performed using the FAS, EAS and PPAS. The primary analysis population will be the EAS. All statistical tests and corresponding CIs will be two-sided, unless otherwise specified.

An overview of the primary and secondary efficacy objectives, endpoints, and the associated case definitions is presented in Table 11.

*Table 11 Primary and Secondary Efficacy and Immune Response Objectives, Endpoints, and Associated Case Definitions*

| Objective | Endpoint | Case Definition |
|---|---|---|
| **Primary Efficacy Endpoint** | | |
| To estimate the efficacy of a single or repeated (21 days apart) IM dose of GRAd-COV2 compared to placebo for the prevention of COVID-19 in adults ≥ 18 years of age. | Time to first SARS-CoV-2 RT-PCR-positive symptomatic illness. | Participants are defined as COVID-19 cases if their first case of SARS-CoV-2 RT-PCR-positive symptomatic illness occurs ≥ 28 days post first dose or ≥7 days post second dose of study intervention. Participants will be included in the primary endpoint if they have RT-PCR-confirmed SARS-CoV-2 and meet the following criteria at any point from their initial illness visit at the site (Day 1) through their last illness visit (Day 28):<br>One or more Category A findings<br>**OR**<br>Two or more Category B findings.<br>***Category A findings:***<br>• Pneumonia diagnosed by chest x-ray, or computed tomography scan.<br>• Oxygen saturation of ≤ 94% on room air or requiring either new initiation or escalation in supplemental O2.<br>• New or worsening dyspnea/ shortness of breath.<br>***Category B findings:***<br>• Fever > 100 °F (> 37.8 °C).<br>• New or worsening cough.<br>• Myalgia/muscle pain.<br>• Fatigue that interferes with activities of daily living.<br>• Vomiting and/or diarrhea (only one finding to be counted toward endpoint definition).<br>• Anosmia and / or ageusia (only one finding to be counted toward endpoint definition). |

This document is confidential.

| Objective | Endpoint | Case Definition |
|---|---|---|
| **Key Secondary Efficacy Endpoint** | | |
| To estimate the efficacy of 1 single or repeated (21 days apart) IM dose of GRAd-COV2 compared to placebo for the prevention of severe or critical symptomatic COVID-19. | Time to first case of SARS-CoV-2 RT-PCR-positive severe or critical symptomatic illness. | Participant must have laboratory-confirmed COVID-19 (SARS-CoV-2 RT-PCR-positive) symptomatic illness occurring ≥ 28 days post dose or ≥7 days post second dose of study intervention plus any of the following:<br>• Clinical signs at rest indicative of severe systemic illness (respiratory rate ≥ 30 breaths per minute, heart rate ≥ 125 beats per minute, oxygen saturation ≤ 93% on room air at sea level, or partial pressure of oxygen to fraction of inspired oxygen ratio < 300 mm Hg).<br>• Respiratory failure (defined as needing high-flow oxygen, noninvasive ventilation, mechanical ventilation or extracorporeal membrane oxygenation).<br>• Evidence of shock (systolic blood pressure < 90 mm Hg, diastolic blood pressure < 60 mm Hg, or requiring vasopressors).<br>• Significant acute renal, hepatic, or neurologic dysfunction.<br>• Admission to an intensive care unit.<br>• Death. |
| **Other Secondary Efficacy Endpoints** | | |
| To estimate the efficacy of a single or repeated (21 days apart) IM dose of GRAd-COV2 compared to placebo for the prevention of SARS-CoV-2 asymptomatic infection. | Proportion of participants who have a post-treatment response (negative at baseline to positive post treatment with study intervention) to SARS-CoV-2 Nucleocapsid | Participants who have a post-treatment response (negative at baseline to positive post treatment with study intervention) for SARS-CoV-2 Nucleocapsid antibodies over time. |

This document is confidential.

| Objective | Endpoint | Case Definition |
|---|---|---|
| | antibodies. | |
| To estimate the efficacy of a single or repeated (21 days apart) IM dose of GRAd-COV2 compared to placebo for the prevention of symptomatic COVID-19 using CDC criteria. | Time to first case of SARS-CoV-2 RT-PCR-positive symptomatic illness. | First case of SARS-CoV-2 RT-PCR-positive symptomatic illness for a participant occurring ≥ 28 days post single dose or ≥7 days post second dose of study intervention using the CDC criteria (CDC 2020): No minimum duration: Fever; Shortness of breath; Difficulty breathing. Present for ≥ 2 days: Chills; Cough; Fatigue; Muscle aches; Body aches; Headache; New loss of taste; New loss of smell; Sore throat; Congestion; Runny nose; Nausea; Vomiting; Diarrhea. |
| To estimate the efficacy of 1 single or repeated (21 days apart) IM dose of GRAd-COV2 compared to placebo for the prevention of COVID-19-related Emergency Department visits. | Time to first COVID-19-related Emergency Department visits. | COVID-19-related Emergency Department visits occurring ≥ 28 days post dose or ≥7 days post second dose of study intervention. |
| To evaluate VE to prevent death caused by COVID-19. | Time to COVID-19 related deaths. | COVID-19 related deaths occurring ≥ 28 days post dose or ≥7 days post second dose of study intervention. |
| **Secondary Immunogenicity Endpoints** | | |
| To assess antibody responses to S antigen following a single or repeated (21 days apart) IM dose of GRAd-COV2 or placebo and persistence of immune response.<br><br>(Phase II and Illness Visits of phase III only). | First endpoint:<br><br>S-binding IgG levels and RBD-binding IgG levels. | Not applicable. |
| | Second endpoint:<br><br>Proportion of participants who have a post-treatment sero-response to the S, RBD antigens of GRAd-COV2. | Post-treatment sero-response (≥ 4-fold rise in titers from day of dosing baseline value to 35 days post single dose and 14 days post second dose) to the S, RBD antigens of GRAd-COV2. |
| To determine anti-SARS-CoV-2 neutralizing antibody levels in serum following 1 single or | First endpoint:<br><br>SARS-CoV-2 neutralizing | Not applicable. |

This document is confidential.

| Objective | Endpoint | Case Definition |
|---|---|---|
| repeated (21 days apart) IM dose of GRAd-COV2 or placebo.<br>(Phase II and Illness Visits of phase III only) | titers. | |
| | Second endpoint:<br>Proportion of participants who have a post-treatment sero-response to GRAd-COV2 as measured by SARS-CoV-2 neutralizing antibodies. | Post-treatment sero-response (≥ 4-fold rise in titers from day of dosing baseline value to 35 days post single dose and 14 days post second dose) to GRAd-COV2 as measured by SARS-CoV-2 neutralizing antibodies. |

CDC = Centers for Disease Control and Prevention; COVID-19 = coronavirus disease 2019; GMFR = geometric mean fold rise; GMT = geometric mean titer; IM = intramuscular; S =Spike; RBD = receptor binding domain; RT-PCR = reverse transcriptase polymerase chain reaction; SARS-CoV-2 = severe acute respiratory syndrome-coronavirus-2.

###### 13.5.3.1 Analysis for Deciding Phase III Expansion and Dose Selection in Phase II Part of the Study

Decision about phase III start and dose selection will be based on safety analyses and immunogenicity data, which will be conducted when five-week post first dose immunogenicity and safety data (all types of AEs) are available for the first 450 participants and one-week safety data (local and systemic solicited AEs) post second dose are available for all 900 participants of this study phase. Immunogenicity data will be available 35 days post single dose and 14 days post second dose. For the between group comparisons, the Day 36 data (ie, 14 days after second dose) will be primary. Details on the analysis of immunogenicity data are provided in Section 13.5.4, while analysis of safety data is described under Section 13.5.5.

###### 13.5.3.2 Primary Efficacy Analysis

The primary efficacy endpoint will be formally assessed at two milestones during the study, ie, at the efficacy interim analysis (see Section 13.9) and at the primary analysis. The timing of these analyses will be driven by the number of cases observed in the study within the subset of participants who are not seropositive at baseline. The interim analysis will be carried out when approximately 37 events meeting the primary efficacy endpoint definition have been observed, and the primary analysis will be carried out when approximately 74 events meeting the primary efficacy endpoint definition have been observed.

A final analysis will also be carried out when all participants have completed the 2-year study to assess durability of VE. However, while the first two analyses are adjusted for multiplicity (through the O'Brien Fleming spending function), the final analysis will not be controlled for multiplicity and statistical hypotheses will be tested at a nominal 5% significance level based on a two-sided test.

*Primary efficacy estimand*

The primary estimand will be used for the analysis of the primary efficacy endpoint. It will be based on participants in the EAS (see Section 13.4). For participants with multiple events, only the first occurrence will be used for the primary efficacy endpoint analysis.

The estimand will the VE point estimate (=1-estimated HR) with the corresponding CI (at final analysis: a 95.1% two-sided CI or equivalently a 97.55% one-sided CI; at interim analysis: a 99.7% two-sided CI or equivalently a 99.85% one-sided CI). A Cox proportional hazard model will be used to assess the magnitude of the HR between GRAd-COV2 and placebo.

The set of intercurrent events for this estimand may include:

1) Participants who withdraw from the study prior to having met the primary efficacy endpoint or who die for reasons that are unrelated to COVID-19.
2) Participants who experience an early COVID-19 infection up to 28 days post single or 7 days post repeated study vaccination.
3) In case the two-dose schedule is selected for phase III, missing second dose of vaccine is an additional intercurrent event.

The intercurrent events listed at points 1) and 2) will be handled using a “while on treatment” strategy, where the time to COVID-19 is censored at the date of withdrawal from the study or death in the former case and to the time of early COVID-19 infection in the latter.

The subjects missing the second dose (see point 3) above) will be excluded from the primary analysis.Participants who are sero-positive at baseline (pre-dose) will be excluded from primary endpoint analysis.

Missing data will not be imputed in the primary analysis.

The same estimand described above but assessed in the FAS will be used to evaluate the efficacy of GRAd-COV2 compared to placebo against confirmed COVID-19 in all participants independently on whether they have or not evidence of infection before vaccination.

Additional estimands will be specified in the SAP for the primary efficacy endpoint to carry out sensitivity analyses for assessing the robustness of results of the primary analysis. These sensitivity analyses will explore different methods for handling intercurrent events and different assumptions for missing data (see Section 13.7).

Primary efficacy analysis

A Cox proportional hazard model with treatment group and protocol stratification factor as independent fixed effects will be used to assess the magnitude of the treatment effect, ie, the HR between GRAd-COV2 and placebo at a two-sided 0.05 study-wise significance level. VE will be estimated as 1–HR.

This document is confidential.

At the interim analysis, the VE will be presented with a two-sided 99.7% CI, and statistical significance will be achieved if the lower limit of this CI is > 30%. The success criterion for the interim analysis will be statistical significance with an observed VE point estimate of at least 50%.

At the primary analysis, VE will be presented with a two-sided 95.1% CI, and statistical significance will be achieved if the lower limit of this CI is > 30%.

The CIs will be based on the O'Brien Fleming alpha-spending function for two-group sequential tests.

###### 13.5.3.3 Key Secondary Efficacy Endpoint

The key secondary endpoint, ie, Time to first case of SARS-CoV-2 RT-PCR-positive severe or critical symptomatic illness will be analyzed following the same methodology described for the primary endpoint. In brief, the estimand will be the VE point estimate (=1-estimated HR) and will be based on participants in the EAS (see Section 13.4). As for intercurrent events, in addition to the ones defined for the primary endpoint, there are the non-severe COVID-19 cases. These will be censored at the time of COVID-19 infection.

This endpoint will be tested in a confirmatory way only if the primary endpoint reaches statistical significance at the required alpha level (depending on interim or final analysis).

###### 13.5.3.4 Other Secondary Efficacy Endpoints

The set of secondary endpoints concerning efficacy are summarized in Table 11, while the population level summaries for these endpoints are described in Table 12.

The proportion of participants who have a post-treatment response (negative at baseline to positive post treatment with study intervention) to SARS-CoV-2 Nucleocapsid antibodies will be derived for the vaccine and control groups by visit, with corresponding 95% Clopper-Pearson exact two-sided CIs. VE will be estimated as 1-incidence ratio (active/placebo). This analysis will be performed on the AAS.

All endpoints based on time to event will be analyzed following the same methodology outlined for the primary endpoint. HRs will be estimated from each model, with corresponding 95% CIs (two-sided). P-values, corresponding to two-sided tests, will be presented to compare the vaccine against the control.

All p-values will be nominal as secondary endpoints are not controlled for multiplicity. To support these analyses, descriptive statistics will be produced for the vaccine and control groups. Full details will be documented in the SAP.

*Table 12 Estimands of Secondary Efficacy and Immune Response Objectives*

This document is confidential.

| Objective | Population Level Summary |
|---|---|
| **Efficacy** | |
| To estimate the efficacy of a single or repeated (21 days apart) IM dose of GRAd-COV2 compared to placebo for the prevention of asymptomatic SARS-CoV-2 infection. | VE point estimate (=1-estimated HR) with the corresponding 95% two-sided CI. |
| To estimate the efficacy of a single or repeated (21 days apart) IM dose of GRAd-COV2 compared to placebo for the prevention of symptomatic COVID-19 using CDC criteria. | As the primary estimand but using the CDC criteria (see Table 11). |
| To estimate the efficacy of one single or repeated (21 days apart) IM dose of GRAd-COV2 compared to placebo for the prevention of severe or critical symptomatic COVID-19. | As the primary estimand but considering only the severe and critical cases (see Table 11). |
| To estimate the efficacy of one single or repeated (21 days apart) IM dose of GRAd-COV2 compared to placebo for the prevention of COVID-19-related Emergency Department visits. | As the primary estimand but considering the time to first COVID-19 related Emergency Department admission. |
| To evaluate VE to prevent death caused by COVID-19. | As for the primary estimand but considering the COVID-19 cases that had a death outcome. |
| **Immunogenicity** | |
| To assess antibody responses to S antigen following a single or repeated (21 days apart) IM dose of GRAd-COV2 or placebo.<br>(900 participants - Phase II and Illness Visits of phase III only). | Post treatment GMTs and GMFRs from day of dosing to 35 days post single dose and 14 days post second dose in SARS-CoV-2 S and RBD antibodies, and percentage of participants with titers greater than defined threshold(s), at baseline and each follow-up visit after vaccination. |
| To determine anti-SARS-CoV-2 neutralizing antibody levels in serum following one single or repeated (21 days apart) IM dose of GRAd-COV2 or placebo.<br>(900 participants Phase II and Illness Visits of phase III only) | Post treatment GMTs and GMFRs from day of dosing to 35 days post single dose and 14 days post second dose in SARS-CoV-2 neutralizing antibodies, and percentage of participants with titers greater than defined threshold(s), at baseline and each follow-up visit after vaccination. |

AE = adverse event; AESI = adverse event of special interest; CDC = Centers for Disease Control; COVID-19 = coronavirus disease 2019; GMFR = geometric mean fold rise; GMT = geometric mean titer; HR=Hazard Ratio; IM = intramuscular; MAAE = medically attended adverse event; RT-PCR = reverse transcriptase polymerase chain reaction; RBD = receptor binding domain; S = Spike; SAE = serious adverse event; SARS-CoV-2 = severe acute respiratory syndrome-coronavirus-2; VE = vaccine efficacy.

This document is confidential.

###### 13.5.3.5 Exploratory Endpoints

The endpoint of duration of symptoms will be assessed by determining the total number of days that a study participant with COVID-19 remains symptomatic through daily assessments after diagnosis.

Vaccine efficacy to prevent all-cause mortality will be assessed by similar analysis methods as used for the primary endpoint analysis, using FAS, EAS, and PPAS. All deaths, regardless of cause from the time of randomization, will be included. If the number of deaths becomes large enough to warrant analysis, the same Cox proportional hazard model described for the primary objective will be applied.

The Burden of Disease (BOD) is defined as illustrated in Table 13, based on the post SARS-CoV-2 infection follow-up.

*Table 13 Burden of Disease Score*

| Patient State | BOD Score |
|---|---|
| Uninfected/Asymptomatic infection | 0 |
| Symptomatic without hospitalization | 1 |
| Hospitalization | 2 |
| Death | 3 |

Abbreviations: BOD=burden of disease score.

A BOD score will be computed for each participant to compare the severity of symptoms between treatment groups.
Other exploratory endpoints may be added with justification provided in the SAP and CSR.

###### 13.5.3.6 Secondary Immune Response Endpoints

The analyses of immunogenicity data will be primarily performed using the IAS. An additional analysis will be performed based on all available participants with immunogenicity data (AAIAS) if there is a large enough difference in sample size (≥20%) between the AAIAS and the IAS populations. All statistical tests and corresponding CIs will be two-sided. The alpha level will be 2.5% for the comparisons of each active dose vs. placebo and 5% for the comparison between the two active doses (if both are statistically significant vs. placebo). Analogously, the CIs will be at the 97.5% and 95% confidence levels, in the two cases respectively.

The set of secondary endpoints concerning immune response are summarized in Table 11 and their population level summaries are described in Table 12.

The proportion of participants who have a post-treatment sero-response to the S and RBD antigens of GRAd-COV2 will be computed for the vaccine and control groups, with corresponding two-sided Clopper-Pearson exact CIs. Similarly, the proportion of participants who have a post-treatment sero-response to GRAd-COV2 as measured by SARS-CoV-2 neutralizing antibodies will be computed for the vaccine and control groups, with corresponding two-sided Clopper-Pearson exact CIs.

This document is confidential.

The GMT and GMFR endpoints will be analyzed on the natural log scale by separate analysis of variance (ANOVA) models, which will include the treatment and the protocol defined strata as categorical covariates. GMFRs will be limited to participants with non-missing values prior to vaccination and at the post-vaccination time point. On the log scale, the models will be used to estimate a mean response for the two vaccine and control groups and their difference (vaccine - control). Then, these values will be back transformed to give geometric means for the two vaccine and control groups and ratio of geometric means (vaccine/control), with corresponding two-sided CIs. A p-value, corresponding to a two-sided test, will be presented to compare each vaccine group against the placebo group for each of the two analyses.

To support these analyses, descriptive statistics will be produced for each of the two vaccine groups and the control group.

At the final analysis on all 900 subjects, efficacy will be assessed on these subjects and, if there is a sufficient number of COVID-19 cases in this group, immunogenicity data will be correlated with the efficacy data.

Full details will be documented in the SAP.

##### 13.5.4 Safety

###### 13.5.4.1 Primary Safety Endpoints Overview

The safety analyses will be performed in the SA set.

The safety of GRAd-COV2 will primarily be assessed by:

- Incidence of local and systemic solicited AEs for 7 days post each dose of study intervention (phase II only).
- Incidence of AEs for 28 days post each dose of study intervention.
- Incidence of SAEs, MAAE (defined in Section 8.3.8) and AESIs (defined in Section 12.3.9) from Day 1 post treatment through Day 360.

At the interim analysis concerning the phase II part of the study the percentage of participants reporting the following events will be computed, separately for each of the three treatment groups:

- Solicited local reactions for up to 7 days following intervention for 900 participants.
- Solicited systemic events for up to 7 days following intervention for 900 participants.
- AEs/SAEs/MAAE/AESIs from day 1 up to 7 days after the second dose for 900 participants.
- AEs/SAEs/MAAE/AESIs from day 1 up to 35 days after the first dose for 450 participants.

This document is confidential.

Two-sided 95% exact CIs using Clopper-Pearson method will be provided for the percentage of participants with any solicited (local and systemic, separately) adverse reactions for each treatment group.

All other safety analyses will be performed on the two study phases combined (except for the dose group that will not be selected for phase III, which will be analyzed separately). The safety analyses will be performed at every planned analysis on the data that will be available by the time of performing that analysis.

AEs, SAEs, MAAEs, and AESIs will be coded using the most recent version of the Medical Dictionary for Regulatory Activities (MedDRA). These events will be presented for each treatment group by system organ class and preferred term. These tables will include the number and percentage of participants reporting at least one event, the number of events and the observation time adjusted rates, where appropriate. These tables will also be stratified by severity grade and relationship to the investigational product as assessed by the investigators. AEs leading to discontinuation from second dose will be summarized.

A listing will provide details for each individual AE / SAE / MAAE / AESI. Full details of all analyses will be provided in the SAP.

###### 13.5.4.2 Other Safety Endpoints

For SARS-CoV-2-positive participants, vital sign measurements and targeted physical examinations will be performed as specified in the SoA (see Section 3).

Standard descriptive analyses will be presented for observed values and changes from baseline. Details of these analyses will be provided in the SAP.

The concomitant medications will be tabulated by treatment group and overall.

##### 13.6 Methods for Multiplicity Control

In the analysis of immunogenicity endpoints for dose comparison, multiplicity has been addressed using the Bonferroni approach for the comparisons of each vaccine group vs placebo. In case both active groups separate from placebo, the comparison between the two active doses will be carried out at the 5% significance level. This procedure ensures that the overall alpha level for these multiple comparisons is 5% for the closure principle.

The primary efficacy endpoint will be assessed at two time points during the study, ie, at the interim analysis and at the primary analysis.

The O'Brien Fleming alpha-spending function has been used to account for multiplicity, where the Type I error is 0.3% at the interim analysis and 0.49% at the primary analysis such that the overall Type I error is controlled at 5% two-sided. Thus, the interim and primary analyses will present estimates with two-sided 99.7% and 95.1% CIs, respectively, and statistical significance will be achieved if the lower bounds of these CIs are > 30%. At the interim or primary analysis, the success criterion for the study will be statistical significance with an observed VE point estimate of at least 50%.

This document is confidential.

For testing the key secondary endpoint, a hierarchical testing approach will be used, ie, it will be tested only if the primary endpoint is statistically significant. The other secondary endpoints will not be controlled for multiplicity. Thus, nominal two-sided p-values will be presented to compare the vaccine against the control, alongside two-sided 95% CIs.

#### 13.7 Sensitivity Analyses

The model assumptions will be checked, and the robustness of the primary analysis will be assessed through various sensitivity analyses. To support the primary analysis, Poisson regression and Beta-binomial models will be fitted to the data. Another sensitivity analysis will be to compute the crude VE without any adjustment, together with the appropriate Clopper-Pearson CI. In the primary analysis, missing data are not imputed. Multiple imputation strategies according to different missing data mechanisms will be applied within these models. Full details of these sensitivity analyses will be specified in the SAP.

#### 13.8 Subgroup Analyses

Subgroup analyses will be carried out to assess the consistency of the treatment effect across key, pre-defined, subgroups. These analyses will focus on the primary efficacy endpoint, but they may be performed also on secondary endpoints, if deemed appropriate. The list of subgroups includes but is not limited to: protocol defined strata, gender, race, and serostatus at baseline. Full details of all subgroup analyses will be described in the SAP, including hypotheses that will be tested and the covariates and interaction terms to be included in the statistical models.

#### 13.9 Interim Analysis

There are two planned interim analyses, one for immunogenicity and safety data in the phase II part of the study and one for efficacy in the phase III part of the study. The study has been adjusted for multiplicity only with respect of the efficacy interim analysis conducted when 50% of the target cases are observed.

The first interim analysis (phase II part of the study) will occur as soon as five-week immunogenicity and safety data are available (and cleaned) for the first 450 subjects and 7-day safety data have been collected (and cleaned) on all 900 subjects of the phase II part of the study. The objective of this interim analysis will be to decide whether to start or not the phase III part of the study and with which vaccine dose (see Section 13.5.3.1).

The interim analysis for efficacy will occur when approximately 50% of total target cases across the two treatment groups have been observed. The primary objective of this interim analysis is early detection of reliable evidence that VE is above 30%. The O'Brien Fleming spending function is used to calculate the efficacy bounds and to preserve the overall (two-sided) 0.05 false positive error rate.

There is no intention to stop the study early if the efficacy is demonstrated at this interim analysis. The DSMB and the Steering Committee will review the interim results and make recommendations to the Sponsor in terms of study results reporting and unblinding, based on the boundaries of early efficacy as described in this Section, safety data, and data external to this study. In addition to possible early efficacy, the DSMB and the Steering Committee will monitor for non-efficacy and vaccine harm; the guiding principles (non-binding) will be provided in the DSMB and Steering Committee Charters, and the details will be provided in the SAP.

Table 14 summarizes the timing, number of cases and decision guidance at the interim and primary analysis. The study has been powered to include this interim efficacy analysis, based on the primary efficacy endpoint.

*Table 14 Interim and Final Analysis Plan for Efficacy*

| Analysis | Number of Cases | Nominal Alpha (two-sided) | Efficacy Bound | Expected Time* |
|---|---|---|---|---|
| Efficacy Interim Analysis | 37 | 0.003 | 2,96 | Approx. 5 months after FPFD |
| Efficacy Primary Analysis | 74 | 0.49 | 1,97 | Approx. 8 months after FPFD |

*Time at which the target # of cases can be reached depends on actual accrual rate and infection incidence rate.
FPFI=First participant first intervention; VE=Vaccine Efficacy.

Indeed, the primary efficacy analysis is also an interim analysis in the sense that it will be conducted when the total number of confirmed target cases have been collected and the study will continue with the objective to collect follow-up data for a total of 24 months for each subject. The study does not treat this analysis as a formal interim analysis, ie, no adjustment for multiplicity has been made. This implies that the analysis with two-year follow-up data will be descriptive.
The SAP will describe the planned interim analyses in greater detail.

#### 13.10 Unblinding for Interim Analyses and Responsibility for Analyses

The Biostatistics department of the CRO in charge for performing all data management and statistics related activities of the study will generate the randomized allocation schedule(s) for study treatment assignment. The randomization will be implemented via an IWR system.

Planned interim analyses and primary analysis are described in Section 13.9.

Unblinding at the level of the participants will be restricted to an independent unblinded statistician and one or more statistical programmers, as needed, performing the interim analyses, who will have no other responsibilities associated with the study.

An external DSMB and a Steering Committee will review the interim data to monitor safety, integrity of the study and to provide recommendation on primary efficacy analyses. These two Committees will review treatment level results of the interim analyses, provided by the independent unblinded statistician(s). Limited additional Sponsor personnel may be unblinded to the treatment level results of the interim analyses, if required, in order to act on the recommendations of the DSMB and the Steering Committee.

Depending on the recommendations from these Committees after the formal interim analysis for efficacy, the Sponsor may decide to prepare a dossier for regulatory submission. In this case, pre-identified Sponsor members, including regulatory staff and the analysis and reporting team, will be unblinded to treatment assignment and remain unblinded for the remaining part of the study. The roles that will be needed for trial continuation will have to be replaced.

The study will continue after the primary analysis up to completing two-years of follow-up. Participants and investigators will remain always blinded till the end of the study.

Details on personnel who will be unblinded and the extent of their unblinding will be described in a specific Data Access Plan, which may be an appendix of the SAP or a separate document.

#### 13.11 Data Monitoring Committee

An independent DSMB will provide oversight, to ensure safe and ethical conduct of the study. During the study, the benefit/risk assessment will be continuously monitored by this Committee to ensure that the balance remains favorable (see also Section 14.1.5).

Further details, composition, and operation of the DSMB will be described in the DSMB Charter.

#### 13.12 Steering Committee

A Steering Committee will provide oversight, to ensure ethical conduct of the study and that tall decisions to be made have a solid scientific background.

Further details, composition, and operation of this Committee will be described in the Steering Committee Charter.

This document is confidential.

### 14 SUPPORTING DOCUMENTATION AND OPERATIONAL CONSIDERATIONS

#### 14.1 Regulatory, Ethical, and Study Oversight Considerations

##### 14.1.1 Regulatory and Ethical Considerations

- This study will be conducted in accordance with the protocol and with the following:
  - Consensus ethical principles derived from international guidelines including the Declaration of Helsinki and Council for International Organizations of Medical Sciences International Ethical Guidelines
  - Applicable ICH GCP Guidelines
  - Applicable laws and regulations
- The protocol, protocol amendments, ICF, IB, and other relevant documents (eg, advertisements) must be submitted to an IRB/IEC by the investigator and reviewed and approved by the IRB/IEC before the study is initiated.
- Any amendments to the protocol will require IRB/IEC and applicable Regulatory Authority approval before implementation of changes made to the study design, except for changes necessary to eliminate an immediate hazard to study participants.
- The Sponsor will be responsible for obtaining the required authorizations to conduct the study from the concerned Regulatory Authority. This responsibility may be delegated to a CRO but the accountability remains with the Sponsor.
- The investigator will be responsible for providing oversight of the conduct of the study at the site and adherence to requirements of 21 CFR, ICH guidelines, the IRB/IEC, European Regulation 536/2014 for clinical studies (if applicable), European Medical Device Regulation 2017/745 for clinical device research (if applicable), and all Food and Drug Administration (FDA) Regulations, as applicable and all other applicable local regulations

**Regulatory Reporting Requirements for SAEs**

- Prompt notification by the investigator to the Sponsor of an SAE is essential so that legal obligations and ethical responsibilities towards the safety of participants and the safety of a study intervention under clinical investigation are met.
- The Sponsor has a legal responsibility to notify both the local regulatory authority and other regulatory agencies about the safety of a study intervention under clinical investigation. The Sponsor will comply with country-specific regulatory requirements relating to safety reporting to the regulatory authority, IRBs/IECs, and investigators.

This document is confidential.

- For all studies except those utilizing medical devices, investigator safety reports must be prepared for suspected unexpected serious adverse reactions (SUSAR) according to local regulatory requirements and Sponsor policy and forwarded to investigators as necessary.
  - European Medical Device Regulation 2017/745 for clinical device research (if applicable), and all other applicable local regulations

An investigator who receives an investigator safety report describing an SAE or other specific safety information (eg, summary or listing of SAEs) from the Sponsor will review and then file it along with the Investigator's Brochure and will notify the IRB/IEC, if appropriate according to local requirements.

##### 14.1.2 Financial Disclosure

Investigators and sub-investigators will provide the Sponsor with sufficient, accurate financial information as requested to allow the Sponsor to submit complete and accurate financial certification or disclosure statements to the appropriate regulatory authorities. Investigators are responsible for providing information on financial interests during the course of the study and for 1 year after completion of the study.

##### 14.1.3 Informed Consent Process

- The investigator or his/her representative will explain the nature of the study to the participant or his/her legally authorized representative and answer all questions regarding the study. Participants must be informed that their participation is voluntary and they are free to refuse to participate and may withdraw their consent at any time and for any reason during the study. Participants or their legally authorized representative will be required to sign a statement of informed consent that meets the local regulations, ICH guidelines, and, where applicable, the IRB/IEC or study center requirements
- The study medical record must include a statement that written informed consent was obtained before the participant was enrolled in the study and the date the written consent was obtained. The authorized person obtaining the informed consent must also sign the ICF.
- Participants must be re-consented to the most current version of the ICF(s) during their participation in the study if required by the ECs.
- A copy of the ICF(s) must be provided to the participant or the participant's legally authorized representative.

Participants who are rescreened are required to sign a new ICF.

The ICF will contain a separate section that addresses and documents the collection and use of any mandatory and/or optional human biological samples. The investigator or authorized designee will explain to each participant the objectives of the analysis to be done on the samples and any potential future use.

This document is confidential.

Participants will be told that they are free to refuse to participate in any optional samples or the future use and may withdraw their consent at any time and for any reason during the retention period.

In addition to the traditional paper based ICF, in this study investigators and patients will have the option to discuss and sign the consent form in an electronic format (Genius ENGAGE), with the support of multimedia materials that will facilitate the understanding of the most relevant information of the ICF.

##### 14.1.4 Data Monitoring and Protection

- Participants will be assigned a unique randomization (identifier) code. Any participant records or datasets that are transferred to the Sponsor will contain the identifier only;
- Participant names or any information which would make the participant identifiable will not be transferred.
- The participant must be informed that his/her personal study-related data will be used by the Sponsor in accordance with the GDPR as well as with any other local data protection regulation. The level of disclosure and use of their data will be also explained to the participant in the informed consent.
- The participant must be informed that his/her medical records may be examined by Clinical Quality Assurance auditors or other authorized personnel appointed by the Sponsor, by appropriate IRB/IEC members, and by inspectors from regulatory authorities.
- If for healthy or logistics reasons related to the pandemic outbreak, on-site monitoring visits will not be possible at the planned time intervals, such monitoring visits will be conducted in a virtual mode. A purpose-built source document workspace (Genius SITE VAULT) under the full and unique control of the site staff and completely separated from the study database will be implemented. Site staff will upload certified electronic copies of the source documents and will allow the study site monitor to conduct the verification of the source documents against the eCRF data. Encryption protocols as well audit trails assure full security and regulatory and GDPR compliance.

##### 14.1.5 Committees Structure

A COVID-19 Vaccine DSMB and Steering Committee (SC) organized by the Sponsor, comprised of independent experts will be convened to provide oversight, to ensure safe and ethical conduct of the study. The COVID-19 Vaccine DSMB and SC will facilitate the interim analysis for safety and efficacy and have the responsibility of evaluating cumulative safety and other clinical study data at regular intervals and making appropriate recommendations based on the available data. During the study, the benefit/risk assessment will be continuously monitored by the COVID-19 Vaccine DSMB and SC to ensure that the balance remains favorable. For example, events of potential vaccine-associated enhanced respiratory disease will be evaluated by periodic reviews of COVID-19 cases by the DSMB and SC. Specifically, the study will be paused for DSMB review if a statistically significantly higher risk ratio (> 1), at the 1-sided 5% significance level, is seen for cases of severe COVID-19 in the vaccine arm compared to the placebo arm.

This assessment for a potentially increased risk ratio will begin after 8 cases of severe COVID-19 have accrued in the study. Based on the output of the review, the study could be paused for further evaluation of the potential signal. Full details of the COVID-19 Vaccine DSMB and SC composition and operations can be found in the COVID-19 Vaccine DSMB and SC Charter.

###### 14.1.6 Dissemination of Clinical Study Data

A description of this clinical study will be available on http://www.clinicaltrials.gov as will the summary of the study results when they are available. The clinical study and/or summary of study results may also be available on other websites according to the regulations of the countries in which the study is conducted.

###### 14.1.7 Data Quality Assurance

- All participant data relating to the study will be recorded on eCRF unless transmitted to the Sponsor or designee electronically (e.g. laboratory data). The investigator is responsible for verifying that data entries are accurate and correct by electronically signing the eCRF.
- The investigator must maintain accurate documentation (source data) that supports the information entered in the eCRF.
- The investigator must permit study-related monitoring, audits, IRB/IEC review, and regulatory agency inspections and provide direct access to source data documents.
- Monitoring details describing strategy (e.g. risk-based initiatives in operations and quality such as Risk Management and Mitigation Strategies and Analytical Risk-Based Monitoring), methods, responsibilities and requirements, including handling of noncompliance issues and monitoring techniques (central, remote, or on-site monitoring) are provided in the relevant study plans.
- The Sponsor or designee is responsible for the data management of this study including quality checking of the data.
- The Sponsor assumes accountability for actions delegated to other individuals (e.g. Contract Research Organizations).
- Study monitors will perform ongoing source data verification to confirm that data entered into the eCRF by authorized site personnel are accurate, complete, and verifiable from source documents; that the safety and rights of participants are being protected; and that the study is being conducted in accordance with the currently approved protocol and any other study agreements, ICH GCP, and all applicable regulatory requirements.
- Records and documents, including signed ICFs, pertaining to the conduct of this study must be retained by the investigator for 15 years after study completion unless local regulations or institutional policies require a longer retention period. No records may be destroyed during the retention period without the written approval of the Sponsor. No records may be transferred to another location or party without written notification to the sponsor.

This document is confidential.

##### 14.1.8 Source Documents

- Source documents provide evidence for the existence of the participant and substantiate the integrity of the data collected. Source documents are filed at the investigator's site.
- Data entered in the eCRF that are transcribed from source documents must be consistent with the source documents or the discrepancies must be explained. The investigator may need to request previous medical records or transfer records, depending on the study. Also, current medical records must be available.

##### 14.1.9 Study and Site Start and Closure

The first act of recruitment is the first participant screened and will be the study start date.

The Sponsor designee reserves the right to close the study site or terminate the study at any time for any reason at the sole discretion of the Sponsor. Study sites will be closed upon study completion. A study site is considered closed when all required documents and study supplies have been collected and a study-site closure visit has been performed.

The investigator may initiate study-site closure at any time, provided there is reasonable cause and sufficient notice is given in advance of the intended termination.

Reasons for the early closure of a study site by the Sponsor or investigator may include but are not limited to:

- Failure of the investigator to comply with the protocol, the requirements of the IRB/IEC or local health authorities, the Sponsor's procedures, or GCP guidelines
- Inadequate recruitment of participants by the investigator
- Discontinuation of further study intervention development

If the study is prematurely terminated or suspended, the Sponsor shall promptly inform the investigators, the IECs/IRBs, the regulatory authorities, and any contract research organization(s) used in the study of the reason for termination or suspension, as specified by the applicable regulatory requirements. The investigator shall promptly inform the participant and should assure appropriate participant therapy and/or follow-up.

Participants from terminated sites may have the opportunity to be transferred to another site to continue the study.

##### 14.1.10 Publication Policy

- The results of this study may be published or presented at scientific meetings. If this is foreseen, the investigator agrees to submit all manuscripts or abstracts to the Sponsor before submission. This allows the Sponsor to protect proprietary information and to provide comments.

This document is confidential.

- The Sponsor will comply with the requirements for publication of study results. In accordance with standard editorial and ethical practice, the Sponsor will generally support publication of multicenter studies only in their entirety and not as individual site data. In this case, a coordinating investigator will be designated by mutual agreement.
- Authorship will be determined by mutual agreement and in line with International Committee of Medical Journal Editors authorship requirements.

This document is confidential.

### 16 APPENDICES

This document is confidential.

### APPENDIX 1. Adverse Events: Definitions and Procedures for Recording, Evaluating, Follow-up, and Reporting

#### Appendix 1.1 Definition of Adverse Events

An AE is the development of any untoward medical occurrence in a patient or clinical study participant administered a medicinal product and which does not necessarily have a causal relationship with this treatment. An AE can therefore be any unfavorable and unintended sign (eg, an abnormal laboratory finding), symptom (for example nausea, chest pain), or disease temporally associated with the use of a medicinal product, whether or not considered related to the medicinal product.

The term AE is used to include both SAEs and non-SAEs and can include a deterioration of a pre-existing medical occurrence. An AE may occur at any time, including run-in or washout periods, even if no study intervention has been administered.

#### Appendix 1.2 Definition of Serious Adverse Events

An SAE is an AE occurring during any study phase (ie, run-in, treatment, washout, follow-up), that fulfils one or more of the following criteria:

- Results in death
- Is immediately life-threatening
- Requires in-participant hospitalization or prolongation of existing hospitalization
- Results in persistent or significant disability or incapacity
- Is a congenital abnormality or birth defect
- Is an important medical event that may jeopardize the participant or may require medical treatment to prevent one of the outcomes listed above.

AEs for **malignant tumors** reported during a study should generally be assessed as **SAEs**. If no other seriousness criteria apply, the 'Important Medical Event' criterion should be used. In certain situations, however, medical judgement on an individual event basis should be applied to clarify that the malignant tumor event should be assessed and reported as a **non-SAE**. For example, if the tumor is included as medical history and progression occurs during the study, but the progression does not change treatment and/or prognosis of the malignant tumor, the AE may not fulfil the attributes for being assessed as serious, although reporting of the progression of the malignant tumor as an AE is valid and should occur. Also, some types of malignant tumors, which do not spread remotely after a routine treatment that does not require hospitalization, may be assessed as non-serious; examples in adults include Stage 1 basal cell carcinoma and Stage 1A1 cervical cancer removed via cone biopsy.

**Life Threatening**

'Life-threatening' means that the participant was at immediate risk of death from the AE as it occurred or it is suspected that use or continued use of the study intervention would result in the participant's

This document is confidential.

death. 'Life-threatening' does not mean that had an AE occurred in a more severe form it might have caused death (e.g. hepatitis that resolved without hepatic failure).

**Hospitalization**

Outpatient treatment in an emergency room is not in itself an SAE, although the reasons for it may be (e.g. bronchospasm, laryngeal oedema). Hospital admissions and/or surgical operations planned before or during a study are not considered AEs if the illness or disease existed before the participant was enrolled in the study, provided that it did not deteriorate in an unexpected way during the study.

**Important Medical Event or Medical Treatment**

Medical and scientific judgement should be exercised in deciding whether a case is serious in situations where important medical events may not be immediately life threatening or result in death, hospitalization, disability, or incapacity but may jeopardize the participant or may require medical treatment to prevent one or more outcomes listed in the definition of serious. These should usually be considered as serious.

Simply stopping the suspect drug does not mean that it is an important medical event; medical judgement must be used.

Examples of important medical events include such events as listed below:

- Angioedema not severe enough to require intubation but requiring iv hydrocortisone treatment
- Hepatotoxicity caused by acetaminophen overdose requiring treatment with N-acetylcysteine
- Intensive treatment in an emergency room or at home for allergic bronchospasm
- Blood dyscrasias (e.g. neutropenia or anaemia requiring blood transfusion, etc.) or convulsions that do not result in hospitalization
- Development of drug dependency or drug abuse

**Intensity Rating Scale**

The grading scales found in the US FDA guidance for healthy volunteers enrolled in a preventive vaccine clinical study [20] will be utilized for all events with an assigned severity grading.

It is important to distinguish between serious and severe AEs. Severity is a measure of intensity whereas seriousness is defined by the criteria in this section. An AE of severe intensity need not necessarily be considered serious. For example, nausea that persists for several hours may be considered severe nausea, but not an SAE unless it meets the criteria shown in this section. On the other hand, a stroke that results in only a limited degree of disability may be considered a mild stroke but would be an SAE when it satisfies the criteria shown in this section.

#### Appendix 1.3 A Guide to Interpret the Causality Question

When making an assessment of causality consider the following factors when deciding if there is a 'reasonable possibility' that an AE may have been caused by the IMP.

- Time Course. Exposure to suspect drug. Has the participant actually received the suspect drug? Did the AE occur in a reasonable temporal relationship to the administration of the suspect IMP?
- Consistency with known IMP profile. Was the AE consistent with the previous knowledge of the suspect IMP (pharmacology and toxicology) or drugs of the same pharmacological class? Or could the AE be anticipated from its pharmacological properties?
- De-challenge experience. Did the AE resolve or improve on stopping or reducing the dose of the suspect IMP?
- No alternative cause. The AE cannot be reasonably explained by another etiology such as the underlying disease, other drugs, other host or environmental factors.
- Re-challenge experience. Did the AE reoccur if the suspected IMP was reintroduced after having been stopped?
- Laboratory tests. A specific laboratory investigation (if performed) has confirmed the relationship.

In difficult cases, other factors could be considered such as:

- Is this a recognized feature of overdose of the IMP?
- Is there a known mechanism?

Causality of 'related' is made if following a review of the relevant data, there is evidence for a 'reasonable possibility' of a causal relationship for the individual case. The expression 'reasonable possibility' of a causal relationship is meant to convey, in general, that there are facts (evidence) or arguments to suggest a causal relationship.

The causality assessment is performed based on the available data including enough information to make an informed judgment. With no available facts or arguments to suggest a causal relationship, the event(s) will be assessed as 'not related'.

Causal relationship in cases where the disease under study has deteriorated due to lack of effect should be classified as no reasonable possibility.

#### Appendix 1.4 Medication Error

For the purposes of this clinical study a medication error is an unintended failure or mistake in the treatment process for a ReiThera study intervention that either causes harm to the participant or has the potential to cause harm to the participant.

This document is confidential.

A medication error is not lack of efficacy of the IMP, but rather a human or process related failure while the IMP is in control of the study site staff or participant.

Medication error includes situations where an error.

- Occurred
- Was identified and intercepted before the participant received the IMP
- Did not occur, but circumstances were recognized that could have led to an error Examples of events to be reported in clinical studies as medication errors:
- IMP name confusion
- Dispensing error, e.g. medication prepared incorrectly, even if it was not actually given to the participant
- IMP not administered as indicated, for example, wrong route or wrong site of administration
- IMP not taken as indicated, e.g. tablet dissolved in water when it should be taken as a solid tablet
- IMP not stored as instructed, e.g. kept in the fridge when it should be at room temperature
- Wrong participant received the medication (excluding IRT errors)
- Wrong IMP administered to participant (excluding IRT errors)

Examples of events that **do not** require reporting as medication errors in clinical studies:

- Errors related to or resulting from IRT - including those which lead to one of the above listed events that would otherwise have been a medication error
- Participant accidentally missed IMP dose(s), e.g. forgot to take medication
- Accidental overdose (will be captured as an overdose)
- Errors related to background and rescue medication, or standard of care medication in open label studies

Medication errors are not regarded as AEs but AEs may occur as a consequence of the medication error.

This document is confidential.

### APPENDIX 2. Handling of Human Biological Samples

#### Appendix 2.1 Chain of Custody

A full chain of custody is maintained for all samples throughout their lifecycle.

The investigator at each study site keeps full traceability of collected biological samples from the participants while in storage at the study site until shipment or disposal (where appropriate) and records relevant processing information related to the samples whilst at site.

The sample receiver keeps full traceability of the samples while in storage and during use until used or disposed of or until further shipment and keeps record of receipt of arrival and onward shipment or disposal.

The Sponsor or delegated representatives will keep oversight of the entire life cycle through internal procedures, monitoring of study sites, auditing or process checks, and contractual requirements of external laboratory providers

Samples retained for further use will be stored in the ReiThera-assigned biobanks or other sample archive facilities and will be tracked by the appropriate ReiThera Team during for the remainder of the sample life cycle.

#### Appendix 2.2 Withdrawal of Informed Consent for Donated Biological Samples

The Sponsor ensures that biological samples are destroyed at the end of a specified period as described in the informed consent.

If a participant withdraws consent to the use of donated biological samples, the samples will be disposed of/destroyed/repatriated, and the action documented. If samples are already analyzed, the Sponsor is not obliged to destroy the results of this research.

Following withdrawal of consent for biological samples, further study participation should be considered in relation to the withdrawal processes.

The investigator:

- Ensures participant's withdrawal of informed consent to the use of donated samples is highlighted immediately to the Sponsor or delegate.
- Ensures that relevant human biological samples from that participant, if stored at the study site, are immediately identified, disposed of as appropriate, and the action documented.
- Ensures that the participant and the Sponsor are informed about the sample disposal.

The Sponsor ensures the organization(s) holding the samples is/are informed about the withdrawn consent immediately and that samples are disposed of or repatriated as appropriate, and the action is documented and study site is notified.

This document is confidential.

#### Appendix 2.3 International Airline Transportation Associate 6.2 Guidance Document

LABELLING AND SHIPMENT OF BIOHAZARD SAMPLES

International Airline Transportation Association (IATA) (https://www.iata.org/whatwedo/cargo/dgr/Pages/download.aspx) classifies infectious substances into 3 categories: Category A, Category B or Exempt

Category A Infectious Substances are infectious substances in a form that, when exposure to it occurs, is capable of causing permanent disability, life-threatening or fatal disease in otherwise healthy humans or animals.

Category A Pathogens are, e.g., Ebola, Lassa fever virus. Infectious substances meeting these criteria which cause disease in humans or both in humans and animals must be assigned to UN 2814. Infectious substances which cause disease only in animals must be assigned to UN 2900.

Category B Infectious Substances are infectious Substances that do not meet the criteria for inclusion in Category A. Category B pathogens are, e.g., Hepatitis A, C, D, and E viruses. They are assigned the following UN number and proper shipping name:

- UN 3373 – Biological Substance, Category B
- are to be packed in accordance with UN 3373 and IATA 650

Exempt - Substances which do not contain infectious substances or substances which are unlikely to cause disease in humans or animals are not subject to these Regulations unless they meet the criteria for inclusion in another class.

- Clinical study samples will fall into Category B or exempt under IATA regulations
- Clinical study samples will routinely be packed and transported at ambient temperature in IATA 650 compliant packaging (https://www.iata.org/whatwedo/cargo/dgr/Documents/DGR-60-EN-PI650.pdf).
- Biological samples transported in dry ice require additional dangerous goods specification for the dry-ice content

This document is confidential.

#### APPENDIX 3. Toxicity Grading Scales for Solicited Adverse Events

The toxicity grading scales for the solicited AEs were modified and abridged from the US FDA Guidance on Toxicity Grading Scale for Healthy Adult and Adolescent Volunteers Enrolled in Preventive Vaccine Clinical Trials [20].

- Table 15: Clinical Abnormalities, Local Reactions to Injectable Product
- Table 16: Clinical Abnormalities, Vital Signs
- Table 17: Clinical Abnormalities, Systemic (General or Illness)

*Table 15 Appendix 3 - Table for Clinical Abnormalities: Local Reactions to Injectable Product*

| **Local Reaction to Injectable Product** | **Reaction Grade** | | | |
|---|---|---|---|---|
| | **Mild (Grade 1)** | **Moderate (Grade 2)** | **Severe (Grade 3)** | **Life Threatening(Grade** |
| Pain | Does not interfere with activity | Repeated use of non-narcotic pain reliever > 24 hours or interferes with activity | Any use of narcotic pain reliever or prevents daily activity | ER visit or hospitalization |
| Tenderness | Mild discomfort to touch | Discomfort with movement | Significant discomfort at rest | ER visit or hospitalization |
| Erythema/redness [a, b] | 1-2 inches (2.5–5 cm) | > 2-4 inches (5.1–10 cm) | > 4 inches (> 10 cm) | Necrosis or exfoliative dermatitis |
| Induration/swelling [a, b] | 1-2 inches (2.5–5 cm) | >2-4 inches (5.1–10 cm) | > 4 inches (> 10 cm) | Necrosis |

[a] In addition to grading the measured local reaction at the greatest single diameter, the measurement should
be recorded as a continuous variable. Reactions < 0.25 inches (< 0.6 centimeters) in diameter will not be recorded.

[b] Grade 4 erythema or induration is determined by study site with participant input rather than being recorded directly in Solicited AE e-Diary.

ER = emergency room.

*Table 16 Appendix 3 - Table for Clinical Abnormalities: Vital Signs*

This document is confidential.

| Vital Sign [a] | Vital Signs Grade | | | |
|---|---|---|---|---|
| | **Mild (Grade 1)** | **Moderate (Grade 2)** | **Severe (Grade 3)** | **Potentially Life Threatening (Grade** |
| Fever (°C) [b]<br>(°F) [b] | 37.9-38.4<br>100.1-101.1 | 38.5-38.9<br>101.2-102.0 | 39.0-40<br>102.1-104 | > 40<br>> 104 |
| Tachycardia (beats/minute) | 101-115 | 116- 130 | > 130 | ER visit or hospitalization for arrhythmia |
| Bradycardia (beats/minute) [c] | 50-54 | 45-49 | < 45 | ER visit or hospitalization for arrhythmia |
| Hypertension; systolic (mm Hg) | 141-150 | 151-155 | > 155 | ER visit or hospitalization for malignant |
| Hypertension; diastolic (mm Hg) | 91-95 | 96-100 | > 100 | ER visit or hospitalization for malignant |
| Hypotension; systolic (mm Hg) | 85-89 | 80-84 | < 80 | ER visit or hospitalization for hypotensive shock |
| Respiratory rate (breaths/minute) | 17-20 | 21-25 | > 25 | Intubation |

Note: Record vital signs as adverse events only if clinically relevant and changed from baseline.

[a] Participant should be at rest for vital signs measurements

[b] No recent hot or cold beverages or smoking

[c] Use clinical judgment when characterizing bradycardia among some healthy participant populations, for example, conditioned athletes

ER = emergency room; Hg = mercury.

*Table 17 Appendix 3 – Table for Clinical Abnormalities: Systemic (General or Ilness)*

This document is confidential.

| **Systemic (General)** | **Systemic Grade** | | | |
|---|---|---|---|---|
| | **Mild (Grade 1)** | **Moderate (Grade 2)** | **Severe (Grade 3)** | **Potentially Life Threatening (Grade 4)** |
| Nausea/vomiting | No interference with activity or 1-2 episodes/24 hrs | Some interference with activity or > 2 episodes/24 hrs | Prevents daily activity, required outpatient IV hydration | ER visit or hospitalization for hypotensive shock |
| Chills | No interference with activity | Some interference with activity | Significant; prevents daily activity | ER visit or hospitalization |
| Headache | No interference with activity | Repeated use of non-narcotic pain reliever > 24 hrs or some interference with activity | Significant; any use of narcotic pain reliever or prevents daily activity | ER visit or hospitalization |
| Fatigue | No interference with activity | Some interference with activity | Significant; prevents daily activity | ER visit or hospitalization |
| Myalgia | No interference with activity | Some interference with activity | Significant; prevents daily activity | ER visit or hospitalization |
| **Systemic Illness** | | | | |
| Illness or clinical adverse event (as defined according to applicable regulations) | No interference with activity | Some interference with activity not requiring intervention | Prevents daily activity and required medical intervention | ER visit or hospitalization |

ER = emergency room; hrs = hours; IV = intravenous.

This document is confidential.

### APPENDIX 4. Adverse Events of Special Interest

AESIs Based on Brighton Collaboration Case Definitions [21]

**Generalized Convulsion**

Generalized convulsion is considered an AESI due to the association with immunization encompassing several different vaccines.

**Guillain-Barré Syndrome and Other Immune-mediated Reactions**

As with many vaccines, temporary ascending paralysis (Guillain-Barré syndrome) or immune-mediated reactions that can lead to organ damage may occur, but this should be extremely rare. Guillain-Barré syndrome is often preceded by an infection. This could be a bacterial or viral infection (gastrointestinal, upper respiratory infections including influenza).

The exact mechanism that triggers Guillain-Barré syndrome is unknown, but it is thought to be triggered by antigenic stimulation resulting in demyelination and damage to the peripheral nerves. This is a condition in which people can develop severe weakness and can be fatal.

Weakness or tingling in the legs or arms and upper body are characteristic symptoms of Guillain-Barré syndrome. Severe cases of Guillain-Barré syndrome cause paralysis and are life-threatening.

Other immune-mediated reactions considered AESIs include acute disseminated encephalomyelitis and vasculitis, which are a theoretical concern based on immunopathogenesis.

**Hypersensitivity Including Anaphylaxis/Anaphylactic Reactions**

Serious allergic reactions including anaphylaxis may occur, as with any vaccine. The incidence of this is unknown, but in general is estimated at one per 105 to 106 vaccinations. These acute reactions may be severe and result in death. Acute allergic reactions may include cardio-respiratory, skin-subcutaneous, and gastrointestinal signs and symptoms, such as chest pain, hypotension, dyspnea, bronchospasm, respiratory failure, urticaria, pruritus, angioedema, nausea, vomiting, diarrhea, and collapse.

**Vaccine-associated Enhanced Respiratory Disease**

Disease enhancement following vaccination as judged by the investigator will be monitored. Detailed clinical parameters will be collected from medical records and aligned with agreed definitions as they emerge. These are likely to include, but are not limited to, oxygen saturation, need for oxygen therapy, respiratory rate, need for ventilatory support, imaging, blood test results, and other clinically relevant assessments. Events of potential vaccine-associated enhanced respiratory disease will be evaluated by regular reviews of COVID-19 cases.

This document is confidential.

**Thrombocytopenia**

Thrombocytopenia is considered an AESI due to the association with immunization encompassing several different vaccines.

Thrombocytopenia is a disorder in which there is an abnormally low platelet count; a normal platelet count ranges from 150 000 to 450 000 platelets per µL. General symptoms of thrombocytopenia include bleeding in the mouth and gums, bruising, nosebleeds, and petechiae (pinpoint red spots/rash). Severe bleeding is the major complication, which may occur in the brain or gastrointestinal tract.

This document is confidential.

### APPENDIX 5. PROTOCOL AMENDMENT History

Amendment Date:

Overall Rationale for the Amendment:

| Section # and Name | Description of Change | Brief Rationale | Substantial/ Non-substantial |
|---|---|---|---|

This document is confidential.
